# Statistical analysis plan for the ‘Pharyngeal electrical stimulation for acute stroke dysphagia trial’ (PhEAST) (ISRCTN98886991)

**DOI:** 10.64898/2026.08.11.26360004

**Authors:** Lisa J Woodhouse, Iris I Mhlanga, Cristina Roadevin, Jacqui K Benfield, Lisa F Everton, Gwenllian Wilkinson, Sarah Greatrex, Cameron JC Skinner, Gemma Squires, Amanda Buck, Corinne Latulipe, Kennedy M Cadman, Nikola Sprigg, Kailash Krishnan, Jason P Appleton, Karl Matz, Helle K Iversen, Satish Mistry, Marilyn James, Timothy J England, Shaheen Hamdy, Alan A Montgomery, Philip M Bath, PhEAST Investigators

**Author notes:** Correspondence: Professor Philip M Bath, Stroke Trials Unit, Mental Health & Clinical Neuroscience, University of Nottingham, Nottingham NG7 2UH.

## Abstract

**Introduction:** Post stroke dysphagia is common, associated with poor functional outcome and lacks treatment strategies beyond behaviour therapies delivered by speech & language therapists. Here, we present the statistical analysis plan for the ongoing pharyngeal electrical stimulation for acute stroke dysphagia trial (PhEAST). PES is a candidate treatment for dysphagia present in non-ventilated stroke patients.

**Methods:** PhEAST is an investigator-initiated international prospective randomised open-label blinded-endpoint phase-4 superiority trial involving 650 participants with tube-dependent post-stroke dysphagia. Consenting patients are randomised to PES versus no PES given on top of standard care with PES given daily for 6 days. The primary outcome is the dysphagia severity rating scale (DSR.S), a measure of swallowing impairment, made at days 14 and 90 and analysed using repeated measures regression.

**Conclusion:** We present the statistical analysis plan for the main analyses based on data up to day 90 along with planned secondary analyses including presentation of baseline data, health economics, cognition and extended follow-up to 12 months.

## INTRODUCTION

Post-stroke dysphagia (PSD, swallowing problems after stroke) is common affecting 60% or more of people with acute stroke and is associated with a poor outcome.^1^ Whilst many patients regain safe swallowing, others continue to suffer from dysphagia for weeks, months or even years. Although food, fluids and drugs will initially need to be administered enterally via a nasogastric tube (NGT), a permanent feeding tube (percutaneous endoscopic gastrostomy [PEG], radiologically-inserted gastrostomy [RIG] or jejunostomy tube [J-tube]) will be needed to facilitate discharge from hospital and long-term feeding if dysphagia persists. Poor outcomes result from aspiration, e.g. causing pneumonia, and reduced nutritional intake, e.g. causing malnutrition. Either can lead to poor functional outcome and death. The management of PSD and staff time in hospital result in high healthcare costs so any effective intervention that reduces swallowing impairment would likely have significant health economic benefits.

The mainstay of management of PSD are screening (usually by nurses),^2^ diagnosis and then treatment. Behavioural interventions (e.g. Shaker, chin tuck against resistance and base of tongue exercises), postural changes and compensatory strategies (e.g. thickening drinks, modifying food and clearing swallows) are the most widely used therapeutic approaches.^3^ In most countries, these are administered by speech and language therapists (SLTs, also called speech and language pathologists) although occupational therapists (OTs) have the same role in other countries, e.g. Denmark. SLTs may also administer physical interventions involving thermal or tactile stimulation.^4^ In addition to pharmacotherapy, a variety of devices have also been tested including acupuncture, neuromuscular electrical stimulation (NMES), repetitive transcranial magnetic stimulation (rTMS) and pharyngeal electrical stimulation (PES).^3,4^ None of these are used widely, largely because there is insufficient large high quality trial data to inform guidelines and practice. In a Cochrane meta-analysis, the potential for efficacy was seen for all the above interventions as judged by reductions (significant or non-significant tendencies) in swallowing impairment and dysphagia.^4^ However, most trials were small (N<100) and the meta-analyses showed considerably heterogeneity (l2>50%). Further, there was evidence of publication bias for most interventions to the extent that any evidence of efficacy might disappear if missing trials had been published and included in the analyses.

Although devices generally have a *conformite europeenne* (European conformity) mark and/or FDA approval, gatekeepers to their use, such as insurers, healthcare commissioners or the UK’s National Institute for Health and Care Excellence (NICE) may not approve them for general use. For example, NICE noted for NMES that *“For dysphagia after a stroke, the evidence suggests potential benefit, but is limited in quality and quantity. Therefore, this procedure should only be used with special arrangements for clinical governance, consent, and audit or research”.*^5^ The solution to these concerns is to perform large definitive phase-3/4 randomised controlled trials that assess efficacy or effectiveness, safety and health economics and so provide level 1a/b evidence that influences healthcare gatekeepers, guidelines and clinical practice.

### Pharyngeal electrical stimulation (PES)

PES has a robust theoretical base and a well-defined treatment paradigm, including only requiring 3-6 days of treatment. ^6^ Human swallowing has bilateral representation in the brain with a ‘dominant’ cortex (unrelated to handedness).^6^ Dysphagia often follows a stroke affecting the dominant swallowing cortex and may be exacerbated in recurrent stroke. Swallowing is dependent on afferent feedback via bulbar cranial nerves innervating the pharynx. Increased sensory input from the pharynx, as delivered as PES, has been shown to drive long-term beneficial changes in the cortical control of swallowing ^7^ with reorganisation of the swallowing cortex.^7–9^

A phase-1 study ^10^ suggested that PES should be delivered at 5 Hz for 10 minutes, a paradigm that maximises the effect on brain excitability.^10,11^ In a randomised dose-comparison phase-2a trial in patients with subacute non-ventilated stroke, PES reduced radiological aspiration as measured using the penetration aspiration scale score (PAS).^12^ PES also reduced swallowing impairment (assessed as the dysphagia severity rating scale, DSRS) and length of stay in hospital in patients with PSD in a sham-controlled parallel group phase II trial.^12^ In a NIHR RfPB-funded multicentre phase-2b randomised sham-controlled trial, the study was feasible in respect of recruitment, compliance and retention; PES non-significantly reduced clinical dysphagia and length of stay in hospital.^13^ An individual patient data meta-analysis of these three small phase II trials (n = 73) found that PES significantly reduced aspiration (PAS) and swallowing impairment (DSRS) and was safe and well tolerated.^14^

The multicentre international randomised single-blind phase-3 STEPS trial assessed PES versus sham in 162 patients with a recent non-ventilated ischaemic or haemorrhagic stroke and radiological penetration or aspiration (PAS> = 3). The study was feasible (recruitment, compliance and retention) and PES was safe but did not reduce aspiration (PAS) or swallowing impairment (DSRS) relative to sham.^15^ Likely explanations for the neutral results included: i) undertreatment in the PES group (average stimulation 14.8mA); ii) partial stimulation in the sham group (through assessment of threshold and tolerability levels); and iii) recruitment of patients whose swallow impairment was too mild (and so who were likely to improve anyway).

PHADER was an international single-arm study in 245 patients with unventilated stroke, ventilated stroke, ventilated-related, traumatic brain injury or other causes of neurogenic dysphagia.^16^ PES was associated with improved DSRS and PAS over 90 days, both overall and in each diagnostic group including in both non-ventilated and ventilated stroke. Analyses from the recent PhEED trial and MAPS audit registry also suggest that PES is associated with improved swallowing over a period of at least 90 days.^17^*ref Importantly, there was no control group in these three studies

In spite of this accumulation of clinical trial data and having a CE mark and FDA designation, PES lacks an adequate evidence base on efficacy, safety and information on health economics for non-ventilated stroke patients. Further, NICE say that“… *more research is needed on pharyngeal electrical stimulation. This procedure should only be done as part of a formal research study and a research ethics committee needs to have approved its use. More research is needed on details of patient selection (including the cause of dysphagia and the timing of the intervention) and effects on length of hospital stay compared with usual care”.*^18^ The PhEAST trial is designed to add significantly to the evidence base and address NICE questions.

PES is effective for accelerating decannulation of intubated stroke patients post-ventilation ^19,20^ and there is common agreement for using PES in this group in European, UK and US guidelines.^21^’^23^ Importantly, PhEAST does not address this population but rather focuses on non-ventilated stroke patients with dysphagia.

## METHODS

### Design

The pharyngeal electrical stimulation for acute stroke dysphagia trial (PhEAST) is assessing the safety and effectiveness of PES in 650 patients with PSD from the UK, Austria and *Denmark. Full information is given in the latest protocol (version 12, 19 May 2026: https://stroke.nottingham.ac.uk/pheast/docs/) and a published version of this.^24^ Adults with acute or subacute stroke (recruitment between 2 and 31 days) and who are tube-dependent for fluids and food (functional oral intake scale,^25^ FOIS score 1-3) are randomised 1:1 to six days of PES versus no PES; practically, this amounts to PES vs NGT and all participants receive best medical-SLT/usual care. The primary outcome is swallow impairment assessed using the dysphagia severity rating scale (DSRS) ^12,26^ at 14 and 90 days and blinded to treatment assignment to avoid observer bias. Analysis will compare the two treatment groups using adjusted repeated measures regression. Although PhEAST extends follow-up to 365 days as a result of secondary funding, this was not the original design and we will focus on reporting data up to 90 days in the primary results publication.

The present paper details the statistical analysis plan (SAP) and health economics analysis plan (HEAP) as well as intended secondary publications, as presented in the accompanying supplements. This information is presented during recruitment, blinded to treatment assignment and prior to locking of the trial database so that analyses are not data-driven or selectively reported.^27^ Following on from our practice for the ENOS, TARDIS and RIGHT-2 trials,^28^’^30^ we include not just information on the primary publication (PES vs. no sham) but also detail additional planned secondary publications, including baseline characteristics, health economics, swallowing up to one year and cognition up to one year. The SAP structure follows that published in 2017 by Gamble eta/.^31,32^

### Supporting information

Additional supporting information may be found in the online version of this article with the accompanying Supplements.

#### Supplement

1. Statistical analysis plan.
2. Health economics analysis plan.
3. Description of main results publication with follow-up to 90 days (day 90 was the original final outcome assessment).
4. Description of baseline data publication including demographics and stroke and dysphagia presentation.
5. Description of health economics to 90 days publication. (A similar format will be used out to day 365 if performed.)
6. Description of planned swallowing/dysphagia publication with follow-up to one year.
7. Description of planned cognition/dementia publication with follow-up to one year.
8. Description of planned responder analysis publication.

#### Other planned analyses/publications include

1. Qualitative assessment of PES delivery.
2. Assessment of trial logistics, delivery and challenges.
3. Calculating swallowing and eating scales from clinical information.
4. Prevalence and use of oral trials of fluids/food.

## DISCUSSION

PhEAST is the largest randomised trial of PES to date for any indication and is designed to provide definitive evidence on effectiveness and safety in patients with tube-dependent post-stroke dysphagia. Indeed, it may be the largest randomised trial ever in post-stroke dysphagia. Recruitment will complete at the end of October 2026 with a target of 650 participants. We anticipate completing day 90 follow-ups in quarter 1 2027 and presenting the main results at the European Stroke Organisation Conference (Vienna) in May 2027. Follow-ups at days 180 and 365 will complete later in 2027 with intended presentation at the International Stroke Conference (Los Angeles) in February 2028.

## ACKNOWLEDGEMENTS

We thank the patients who joined the trial and their relatives for supporting enrolment; NIHR research delivery network staff who screened, recruited and treated patients; and speech & language therapists who screened and followed-up patient outcomes at day 14. PMB is Stroke Association Professor of Stroke Medicine and is an Emeritus NIHR Senior Investigator.

## STATEMENT OF ETHICS

Study approvals were obtained in the UK from the national research ethics committee (REC): East of England - Essex (approval 21/EE/0252, date 6 Dec 2021), Health Research Agency (7 Jan 2022), Health Research Agency Wales (7 Jan 2022), and Scotland (21/SS/0075, 9 Feb 2022). In Austria, approval was given by the Lower Austrian Ethics Commission (GS1-EK-3/201-2021). In Denmark, approval was given by the Medical Research Ethics Committee (MREC) (16-0302-7). All participants or a relative gave written informed consent/consultee assent.

## AUTHOR CONTRIBUTIONS

- PMB, SH, TJE, AAM, MJ, NS, LFE, HKI, KM are co-investigators contributing to conceptualisation, funding acquisition, methodology, supervision, formal analysis and writing.
- LJW, IIM, CR, JKB, GW, SG, CJCS, GS, AB, CL, KMC, were/are trial management committee members contributing to trial administration, investigation, data curation, software, validation and statistical analysis.
- CR, MJ are health economists and wrote the HEAP.
- JPA, KK are medical monitors.
- SM contributed resources and writing.
- PMB wrote the original draft of this manuscript; all other authors contributed to review and editing.

## DATA AVAILABILITY

Once the trial is completed and published, data from PhEAST will be added to trial-level and individual patient data (IPD) meta-analyses of PES in acute/subacute stroke.^4,14^ IPD will be made available to the ‘virtual international stroke trials archive’ (VISTA) ^33^ and subsequently over the web, as with the International Stroke Trial.^34^

## ABBREVIATIONS

ADE: Adverse device effect
AE: Adverse event
BI: Barthel index
BLR: Binary logistic regression
CF: Informed consent form
CI: Chief Investigator overall
CPHR: Cox proportional hazards regression
CRF: Case Report Form
DAP: Data Analysis Plan
D/D: Death or discharge
DMC: Data Monitoring Committee
DSRS: Dysphagia severity rating scale
EAT-10: Eating assessment tool
eCRF: Electronic case record form
EDAR: Eating and drinking with acknowledged risk
EOT: End of Trial
EQ-VAS: EuroQuality of Life-visual analogue scale
EQ-5D-5L: EuroQoL-five dimensions-5 levels
FEES: Fibreoptic endoscopic evaluation of swallowing
FOIS: Functional oral intake scale
FSS: Feeding status scale
FWP: Free water protocol
GCP: Good Clinical Practice
GCS: Glasgow coma scale
HTA: Health Technology Assessment
ICH: Intracerebral haemorrhage
IDDSI: International dysphagia diet standardisation initiative
IDDSI-FDS: IDDSI-functional diet scale
IQCODE: Informant questionnaire on cognitive decline in the elderly
IS: Ischaemic stroke
ITT: Intention-to-treat
J-tube: Jejunostomy tube
LPFV: Last participant first visit
LPLV: Last participant last visit
mA: Milliampere
MHRA: Medicines and Healthcare products Regulatory Agency
mITT: Modified intention-to-treat
MLR: Multiple linear regression
MMSE: Mini-mental state examination
MoCA: Montreal cognitive assessment
mRS: Modified Rankin scale
NGT: Nasogastric tube
NJT: Nasojejunal tube
NHS: National Health Service
NIHR: National Institute for Health and care Research
NIHSS: National Institutes of Health stroke scale
NMES: Neuro-muscular electrical stimulation
OLR: Ordinal logistic regression
OT: Occupational therapist
PAS: Penetration aspiration scale
PEG: Percutaneous endoscopic gastrostomy
PEJ: Percutaneous endoscopic jejunostomy
PES: Pharyngeal electrical stimulation
PI: Principal Investigator at a local centre
PIS: Participant Information Sheet
PLR: Personal legal representative (health)
PP: Per protocol
PSD: Post stroke dysphagia
REC: Research Ethics Committee
R&D: Research and Development department
RIG: Radiologically-inserted gastrostomy
RMR: Repeated measures regression
rTMS: Repetitive transcranial magnetic stimulation
SADE: Serious adverse device effect
SAE: Serious adverse event
SAR: Serious adverse reaction
SLT: Speech & language therapist
SPC: Summary of Product Characteristics
SWAT: Study within a trial
TMG: Trial Management Group
TSC: Trial Steering Committee
USADE: Unexpected serious adverse device effect
VFS: Videofluoroscopy
ZDS: Zung depression scale

## SUPPLEMENT 1

**STATISTICAL ANALYSIS PLAN - PHARYNGEAL ELECTRICAL STIMULATION FOR ACUTE STROKE DYSPHAGIA TRIAL (PhEAST)**

Authors: Philip M Bath, Lisa J Woodhouse, Alan M Montgomery.

### SECTION 1. ADMINISTRATIVE INFORMATION

**1 Title and trial registration**

***1a Title:*** Pharyngeal electrical stimulation for acute stroke dysphagia trial

***Acronym:*** PhEAST

***1b Registration:*** ISRCTN98886991. IRAS project number: 304658. Research ethics: England 21/EE/0252 (date 06/12/2021), Scotland 21/SS/0075 (date 09/02/2022). WHO Ullll-1273-9942. University of Nottingham sponsor protocol 21068

**2 SAP version:** 1.0 (15 July 2026)

**3 Protocol version:** 11.0 (15 January 2026)

**4 SAP revisions**

***4a Revision history:*** None, this is the first version.

***4b Justification for each revision:*** Not relevant.

***4c Timing of revisions:*** Not relevant.

**5 Roles and responsibilities**

***Authors:*** Philip M Bath, Lisa J Woodhouse, Alan A Montgomery.

***Responsible statisticians:*** Lisa J Woodhouse (senior statistician, blinded), Iris Mhlanga (statistician, unblinded), Alan A Montgomery (consultant statistician, blinded)

***Chief Investigator:*** Philip M Bath (blinded)

***Contributors and roles:*** Philip M Bath, Lisa J Woodhouse, Iris Mhlanga, Alan Montgomery, for the PhEAST Investigators

**6 Signatures**

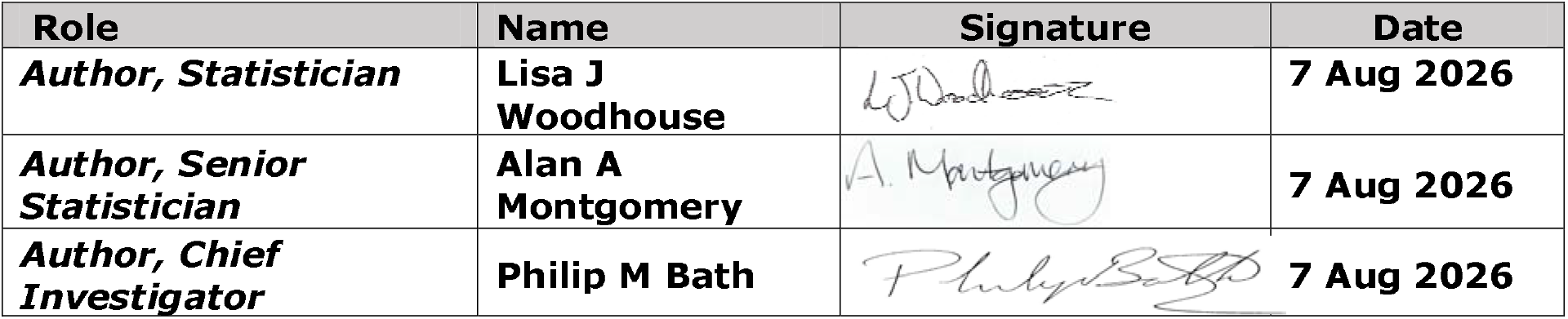

| Role | Name | Signature | Date |
| --- | --- | --- | --- |
| **Author, Statistician** | **Lisa J Woodhouse** |  | **7 Aug 2026** |
| **Author, Senior Statistician** | **Alan A Montgomery** |  | **7 Aug 2026** |
| **Author, Chief Investigator** | **Philip M Bath** |  | **7 Aug 2026** |

### SECTION 2. INTRODUCTION

**7 Background and rationale:** See main text.

**8 Objectives:** To assess whether pharyngeal electrical stimulation (PES) is safe and efficacious at improving post-stroke dysphagia.

***8a Primary Objective:***

To assess whether 6 days of PES versus no PES (on top of best medical/usual care) accelerates return to oral intake of food and drink as assessed using the dysphagia severity rating scale (DSRS) with blinding to treatment.

***8b Secondary Objectives:***

To assess whether PES with usual care versus usual care:

- Improves swallowing and feeding.
- Reduces pneumonia, antibiotic usage, dependence on PEG/RIG feeding, and hospital length of stay. †
- Reduces dependence and disability.
- Is effective in predefined subgroups including by age, sex, cause (stroke type, severity), timing of intervention - see full list in section 27f. †
- Participant subgroups predict response to PES.
- Improves quality-of-life.
- Is cost effective.
- Reduces cognitive impairment.

† Some of objectives are driven, in part, by NICE questions.^18^ NICE refer to PhEAST as part of their review and guidance.

### SECTION 3. STUDY METHODS

**9 Trial design**

PhEAST is an international prospective randomised open-label blinded-endpoint (PROBE) parallel-group superiority phase-4 effectiveness trial of PES versus no PES in 800 hospitalised patients with acute/subacute PSD. All participants receive best medical/usual care. Data collection is performed via a secure internet site with real-time data validation.

**10 Randomisation**

Allocate to PES : no PES 1:1 using minimisation, with 95/5 random element to reduce predictability, on country (non-UK vs UK), age (<75/75+), sex, DSRS (< 12/12), impairment (NIHSS <15/15+), stroke type (ischaemic/haemorrhagic), circulation (anterior/posterior) and time to randomisation (<15/15+ days). (The minimisation-randomisation system is that used in our prior trials: ENOS, TARDIS, PODCAST, TICH-2, RIGHT-2.^35–39^) Including these minimisation variables as covariates in multivariable regression models improves precision of estimated between-group effects and so increases statistical power.^40^

Investigators (doctors, research nurses-coordinators/healthcare professionals, speech and language therapists (SLT) (or occupational therapists [OT] in Denmark may enrol participants. Randomised treatment assignment will occur when essential baseline data are entered into the trial computer system by investigators. As such, allocation is concealed from investigators up to the time that they have screened, consented and collected and completed entry of key baseline data into the trial database.

**11 Sample size/power considerations**

***11a Sample size calculation:*** The primary outcome, dysphagia severity rating scale (DSRS, *S Table 1.1*),^12,26^ will be compared between PES and no PES using multiple linear regression with adjustment for minimisation variables. The null hypothesis (H_0_) is that PES does not alter DSRS at day 14 in participants with PSD. The alternative hypothesis (Hi) is that PES alters DSRS at days 14 and 90 in participants with PSD.

**S Table 1.1:**
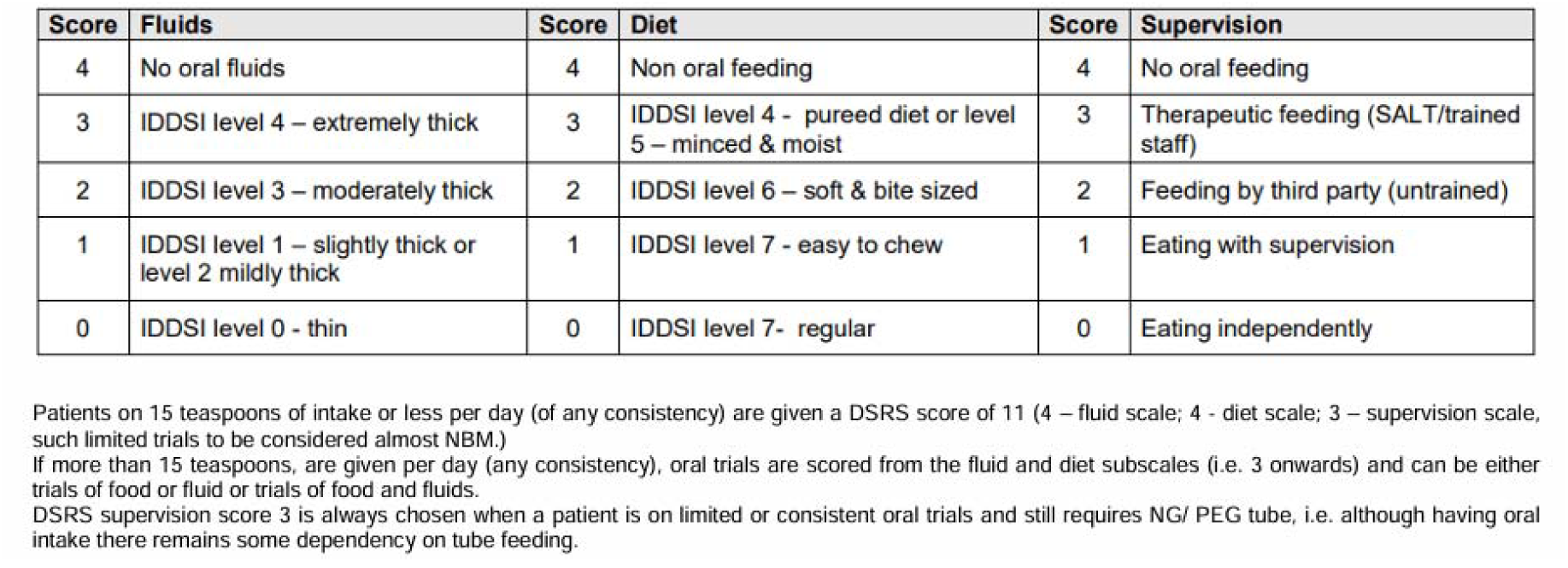
DSRS and subscales including old ‘food’ consistencies and new IDDSI levels.^12,26^.

| Score | Fluids | Score | Diet | Score | Supervision |
| --- | --- | --- | --- | --- | --- |
| 4 | No oral fluids | 4 | Non oral feeding | 4 | No oral feeding |
| 3 | IDDSI level 4 – extremely thick | 3 | IDDSI level 4 - pureed diet or level 5 – minced & moist | 3 | Therapeutic feeding (SALT/trained staff) |
| 2 | IDDSI level 3 – moderately thick | 2 | IDDSI level 6 – soft & bite sized | 2 | Feeding by third party (untrained) |
| 1 | IDDSI level 1 – slightly thick or level 2 mildly thick | 1 | IDDSI level 7 - easy to chew | 1 | Eating with supervision |
| 0 | IDDSI level 0 - thin | 0 | IDDSI level 7- regular | 0 | Eating independently |

Patients on 15 teaspoons of intake or less per day (of any consistency) are given a DSRS score of 11 (4 – fluid scale; 4 - diet scale; 3 – supervision scale, such limited trials to be considered almost NBM.)
If more than 15 teaspoons, are given per day (any consistency), oral trials are scored from the fluid and diet subscales (i.e. 3 onwards) and can be either trials of food or fluid or trials of food and fluids.
DSRS supervision score 3 is always chosen when a patient is on limited or consistent oral trials and still requires NG/ PEG tube, i.e. although having oral intake there remains some dependency on tube feeding.

Assuming 1:1 randomisation; alpha 5% (two-tailed); power 90%; DSRS difference delta=1.2 (this target difference is above the lower level of 0.3 of the estimated minimal clinically important difference ^26^); standard deviation 5.0 *(Table 3); losses* 3% (greater than seen in previous PES trials); crossovers 3%; sample rounded up; a sample size of N=800 is needed (PES n=400, control n=400). The sample size estimation used a standard t test sample size formula. The assumptions are based on pilot trials and STEPS.^12^’^15^ We and others have shown that adjustment for covariates improves statistical power ^41^’^43^ or reduces sample size; however, we have not taken account of this in the above sample size calculation since the relevance of these findings to analysis of DSRS remains unclear. Nevertheless, it is likely that covariate adjustment will improve statistical power so that the final power will probably be greater than assumed here.

This calculation does not take account of the subsequent change in analysis plan which will use DSRS assessments at both days 14 and 90 with analysis using repeated measures regression; see section 26a below for further information.

***11b Sensitivity calculations:*** The sample size varies by the difference in DSRS between PES and no PES (*S Table 1.2*).

**S Table 1.2:**
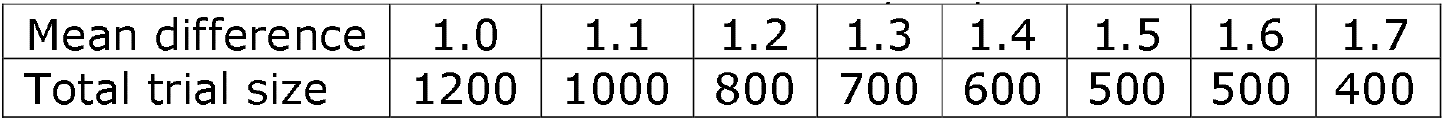
Sample size for a variety of differences in mean DSRS between PES and no PES. Data are shown for the original plan to compare DSRS between Pes and no PES using multiple linear regression. The illustrated mean differences all exceed the lower level of 0.3 of the estimated minimal clinically important difference.^26^

| Mean difference | 1.0 | 1.1 | 1.2 | 1.3 | 1.4 | 1.5 | 1.6 | 1.7 |
| --- | --- | --- | --- | --- | --- | --- | --- | --- |
| Total trial size | 1200 | 1000 | 800 | 700 | 600 | 500 | 500 | 400 |

**S Table 1.3:** Power of trial by achieved final sample size. Data are shown for the revised plan (devised in 2025, approved by TSC in March 2026 during recruitment and prior to data lock and unblinding). 90% power is achieved with 636 participants.

|  |  |  |  |  |  |  |  |  |  |  |  |  |
| --- | --- | --- | --- | --- | --- | --- | --- | --- | --- | --- | --- | --- |
| Size (N) | 822 | 795 | 769 | 742 | 716 | 689 | 663 | 636 | 610 | 583 | 557 | 530 |
| Power (%) | 96 | 95 | 94 | 94 | 93 | 92 | 91 | 90 | 89 | 87 | 86 | 85 |

**12 Framework:** PES will be tested versus no PES for superiority.

**13 Statistical interim analyses and stopping guidance**

***13a Interim analyses:*** One formal interim analysis was planned to be performed to guide the independent Data Monitoring Committee (DMC) at the funder stop-go time-point occurring at 12 months after first enrolment. However, this was not performed because the recruitment of 65 participants at that stage was judged insufficient to justify formal analysis. Since performing the interim analysis later would not influence trial progression, we did not run the analysis.

***13b Adjustments of significance level:*** There is no planned adjustment to the significance level; it remains at P≤.0.05.

***13c Stopping rules:*** The stopping rules for the Data Monitoring Committee regarding effectiveness were based on the combination of presence of’proof beyond a reasonable doubt’ and the likelihood that the results would inform clinical practice.

The possible DMC recommendations at any assessment were:

1. Stop enrolment if the study is negative: statistical evidence that DSRS or fatal SAE rates are significantly higher in the PES than sham group (p<0.01);
2. Stop enrolment if the study is positive: the combination of statistical evidence that DSRS is significantly lower in the PES than sham group “beyond reasonable doubt” and the overall trial results will lead to a change in clinical practice, e.g. by taking account of delta DSRS and evidence that at least some secondary dysphagia-swallowing or other related outcomes are also being benefitted, e.g. one or more of FOIS, IDDSI-FDS, EAT-10, FSS, death, length of hospital stay, pneumonia, antibiotic use.
3. Continue enrolment if the study is neutral: or if conditions 1 and 2 are not present.
4. Modify study design - if it appears that:

a. Sample size calculation assumptions were incorrect, e.g., if standard deviation exceeds 6.0;
b. Apparent study design aspects will lead to incorrect study conclusions;
c. Specific clinical procedures jeopardise the safe execution of the study.

The DMC performed their final assessment of unblinded data in October 2025 and recommended continuation of the trial (as per #3 above).

**14 Timing of final analyses**

Prior to each database lock, the trial managers/trial co-ordinators will chase outstanding data queries and the lock will take place in accordance with the documented data lock procedure once notification has been given to the trial programmer by the chief investigator. Both interim and final locks will be documented and primarily consist of the creation of a read-only copy of the live database, with each copy available to the trial statistician via the online data extract process. The main publication will focus on data up to day 90 as per the original funded plan.

A soft lock will be performed once the final participant has been recruited (LPFV). The trial statistician will share the results of analyses based on this SAP of the interim data with the chief investigator, co-chief investigator and deputy chief investigator; together they will draft the first version of the main publication and will not participate further in participant- or analysis-related decision-making until data cleaning has completed following last participant last visit (LPLV). All other members of the trial team will remain blinded to the results.

The hard lock will be performed once the final participant has had their day 90 visit data collected (LPLV) and data cleaning has completed. The Main publication will be updated with the final results out to day 90 (*Supplement 3*).

Participants will remain in the trial until day 365; analyses for day 180 and 365 data will be performed once all participants have reached day 365 and data cleaning has completed. A secondary publication will focus on these longer-term follow-ups. *(Supplement 6)*.

**15 Timing of outcome assessments:** Outcomes will be assessed blinded-to-allocation at days 14 (protocolised range 13-17), 90 (76-104), 180 (159-201), 365 (337-393) unless otherwise specified (*S Table 1.4*). Day 14 assessments are performed in hospital. Day 90,^36’38’39^’^44–46^ 180 and 365 are performed by central telephone (postal) follow-up. If the participant is an inpatient at day 90, 180 or 365, follow-up may be completed by the hospital site staff. If data are collected in hospital prior to day 90, this will be counted as day 14 if there is no existing day 14 data. If data are collected outside the protocolised windows, data will be assigned as follows:

- Day 14 missing and data collected prior to day 76 will be counted as day 14 data.
- Day 90 missing and data collected prior to day 159 will be counted as day 90 data.
- Day 180 missing and data collected prior to day 337 will be counted as day 365 data.

**S Table 1.4:**
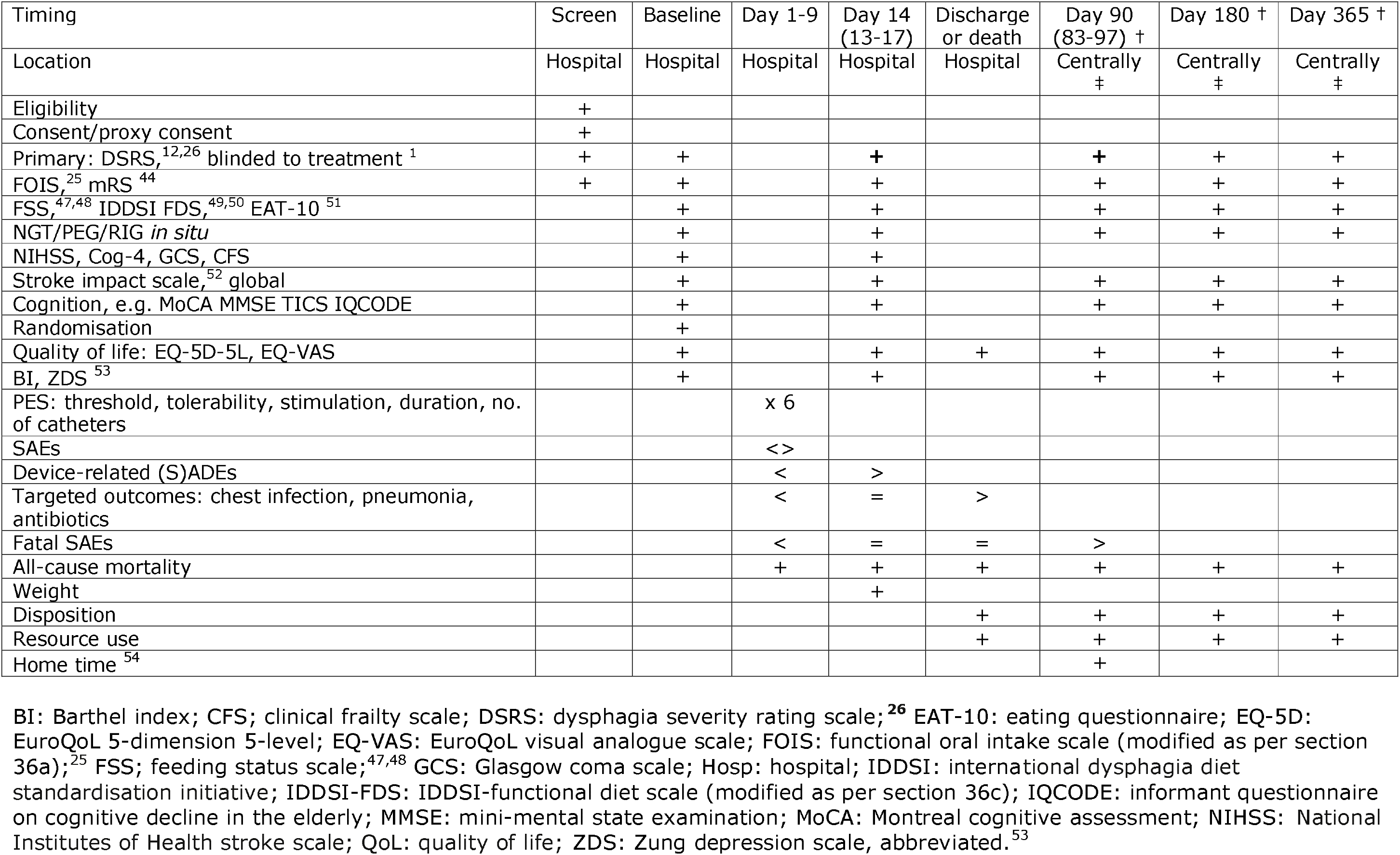
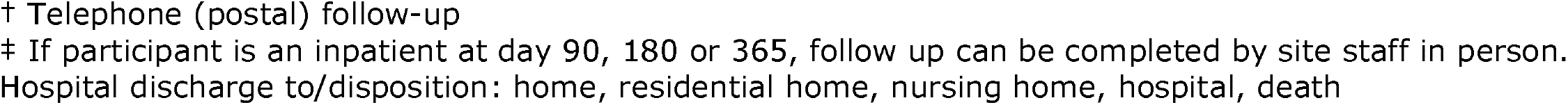
Timing of assessments.

| Timing | Screen | Baseline | Day 1-9 | Day 14 (13-17) | Discharge or death | Day 90 (83-97) † | Day 180 † | Day 365 † |
| --- | --- | --- | --- | --- | --- | --- | --- | --- |
| Location | Hospital | Hospital | Hospital | Hospital | Hospital | Centrally ‡ | Centrally ‡ | Centrally ‡ |
| Eligibility | + |  |  |  |  |  |  |  |
| Consent/proxy consent | + |  |  |  |  |  |  |  |
| Primary: DSRS,<sup>12,26</sup> blinded to treatment<sup>1</sup> | + | + |  | + |  | + | + | + |
| FOIS,<sup>25</sup> mRS<sup>44</sup> | + | + |  | + |  | + | + | + |
| FSS,<sup>47,48</sup> IDDSI FDS,<sup>49,50</sup> EAT-10<sup>51</sup> |  | + |  | + |  | + | + | + |
| NGT/PEG/RIG *in situ* |  | + |  | + |  | + | + | + |
| NIHSS, Cog-4, GCS, CFS |  | + |  | + |  |  |  |  |
| Stroke impact scale,<sup>52</sup> global |  | + |  | + |  | + | + | + |
| Cognition, e.g. MoCA MMSE TICS IQCODE |  | + |  | + |  | + | + | + |
| Randomisation |  | + |  |  |  |  |  |  |
| Quality of life: EQ-5D-5L, EQ-VAS |  | + |  | + | + | + | + | + |
| BI, ZDS<sup>53</sup> |  | + |  | + |  | + | + | + |
| PES: threshold, tolerability, stimulation, duration, no. of catheters |  |  | x 6 |  |  |  |  |  |
| SAEs |  |  | <> |  |  |  |  |  |
| Device-related (S)ADEs |  |  | < | > |  |  |  |  |
| Targeted outcomes: chest infection, pneumonia, antibiotics |  |  | < | = | > |  |  |  |
| Fatal SAEs |  |  | < | = | = | > |  |  |
| All-cause mortality |  |  | + | + | + | + | + | + |
| Weight |  |  |  | + |  |  |  |  |
| Disposition |  |  |  |  | + | + | + | + |
| Resource use |  |  |  |  | + | + | + | + |
| Home time<sup>54</sup> |  |  |  |  |  | + |  |  |

BI: Barthel index; CFS; clinical frailty scale; DSRS: dysphagia severity rating scale;<sup>26</sup> EAT-10: eating questionnaire; EQ-5D: EuroQoL 5-dimension 5-level; EQ-VAS: EuroQoL visual analogue scale; FOIS: functional oral intake scale (modified as per section 36a);<sup>25</sup> FSS; feeding status scale;<sup>47,48</sup> GCS: Glasgow coma scale; Hosp: hospital; IDDSI: international dysphagia diet standardisation initiative; IDDSI-FDS: IDDSI-functional diet scale (modified as per section 36c); IQCODE: informant questionnaire on cognitive decline in the elderly; MMSE: mini-mental state examination; MoCA: Montreal cognitive assessment; NIHSS: National Institutes of Health stroke scale; QoL: quality of life; ZDS: Zung depression scale, abbreviated.<sup>53</sup>
† Telephone (postal) follow-up
‡ If participant is an inpatient at day 90, 180 or 365, follow up can be completed by site staff in person.
Hospital discharge to/disposition: home, residential home, nursing home, hospital, death

### SECTION 4. STATISTICAL PRINCIPLES

**16 Levels of statistical significance**

The result of the analysis of the primary outcome will be shown with a p-value, with P≤.0.05 judged as significant. Otherwise, p-values will not be shown unless essential.

**17 Adjustments for multiplicity**

There will be no adjustment made for multiplicity; rather secondary analyses will be used for hypothesis generating.^55,56^

**18 Confidence intervals**

The results of analyses and comparisons will be reported with 95% confidence intervals (95% CI).

**19 Adherence and protocol deviations**

***19a Definition of adherence:*** Adherence will be assessed by examining the participant’s 6 daily treatment forms as recorded on the REDCap database and based on evidence of treatment administration: threshold, tolerability, calculated stimulation and actual stimulation levels (maximum for each is 50 mA); and exposure assessed as number of treatments received (maximum is 6) and length of each treatment (maximum is 10 minutes).

Adherence will be considered sufficient if actual stimulation level is >20 mA, actual stimulation is the same as calculated stimulation level, six treatments are delivered and each treatment is given for >9 minutes 50 seconds.

***19b Presentation of adherence:*** Adherence will be presented as:

- Mean (standard deviation) for each of threshold, tolerability, calculated stimulation and actual stimulation levels.
- Median [interquartile range] for each of number of treatments and their length.

***19c Protocol violations:*** These are a major deviation from the trial protocol, for example where a participant is enrolled in spite of not fulfilling all the inclusion and exclusion criteria, or where deviations from the protocol could negatively affect participant safety, trial delivery or interpretation. Listed protocol violations are:

1. Enrolment without consent.
2. Enrolment but ineligible.
3. Non-reporting of primary outcome measure.
4. Non-reporting of serious adverse event (SAE), serious adverse device event (SADE), serious unanticipated adverse device event (SUADE).
5. PES treatment beginning after day 1, i.e. not started on day of or after randomisation.
6. Primary outcome (DSRS) at day 14 not performed between days 13 and 17.
7. Primary outcome (DSRS) at day 14 assessor not blinded to treatment assignment.
8. Other dysphagia therapy device(s) used during the trial period of days 0-90.
9. Any other major violation of the trial protocol.

All protocol violations must be reported immediately to the Chief Investigator, via the online electronic case report form. The CI will notify the Sponsor if a violation has an impact on participant safety or integrity of the trial data. The Sponsor will advise on appropriate measures to address the occurrence, which may include reporting of a serious GCP breach, internal audit of the trial and seeking counsel of the trial committees. Violations, deviations and device deficiencies will be presented in listings in the main publication *(Supplement 3)*.

**19d Protocol deviations:** These are minor deviations from the protocol that affect the conduct of the trial in a minor way. This includes any deviation from the trial protocol that is not listed as a Protocol Violation. Deviations will be collected, via the online electronic case report form. The list of deviations is not exhaustive and so there will be a drop-down menu as well as a free text box in the eCRF.

All protocol deviations must be reported immediately to the Chief Investigator via the online electronic case report form (eCRF). The CI will notify the Sponsor if a deviation or violation has an impact on participant safety or integrity of the trial data. The Sponsor will advise on appropriate measures to address the occurrence, which may include reporting of a serious GCP breach, internal audit of the trial and seeking counsel of the trial committees. Deviations will be presented in listings in the main publication *{Supplement* 3).

**19e Device deficiencies:** Apparent device deficiencies (whether of base station or catheters) must be reported on the relevant eCRF; these will be forwarded to the manufacturer for further assessment. Deficiencies will be presented in listings in the main publication *{Supplement 3)*.

**20 Analysis populations**

All available data will be used and levels of missing data will be reported.

1. ***Safety:*** All randomised participants - primary safety analysis.
2. ***Modified intention to treat (mITT):*** All randomised participants with recorded DSRS data at days 14 and/or 90 analysed using repeated measures regression and without imputation - primary efficacy analysis.
3. ***Intention to treat (ITT):*** All randomised participants with recorded or imputed DSRS data at days 14 and 90 analysed using repeated measures regression and imputations made using regression imputation - sensitivity efficacy analysis.
4. ***Per protocol set (PP):*** All participants in the mITT set who are deemed to have no protocol violations that could interfere with the objectives of the study - sensitivity efficacy analysis.

Participants will be included or excluded from analyses of these populations as follows:

***Excluded from all analyses***

- Enrolled without valid consent.
- Enrolled with consent but no randomisation.

***Excluded from per protocol analyses***

- Enrolled but ineligible:

- Already in another trial where a co-enrolment agreement is not in place.
- FOIS >3.
- Randomised <2 or >31 days.
- Randomised to PES but did not receive it.
- Randomised to no PES but did receive it.
- Dysphagia therapy device(s) used between randomisation and day 90.
- Received PES treatment late, i.e. day >1 after randomisation.
- Primary outcome not performed between days 13 to 17 and/or days 76 to 104.
- Non-reporting of primary outcome measure at both days 14 and 90.
- Received active treatment with another device prior to the 90 day follow-up.

### SECTION 5. TRIAL POPULATION

**21 Screening data:** Sites will attempt to keep screening data in logs; where kept, these will be aggregated and presented.

**22 Eligibility**

***22a Inclusion***

1. Hospitalised adults.
2. Age > = 18 years.
3. Recent (2-31 days) ischaemic or haemorrhagic, anterior or posterior circulation, stroke (as diagnosed clinico-radiologically) at a stroke centre.
4. Clinical dysphagia with dependency on tube feeding defined as a functional oral intake scale (FOIS ^25^) score of:

i. Nothing by mouth, feeding by NGT/PEG,
ii. Tube dependent feeding with minimal attempts of food or liquids, or
iii. Tube dependent feeding with consistent oral intake of food or liquids.
5. Baseline DSRS supervision score of either 3 (requiring therapeutic feeding by SALT team; on oral trials) or 4 (No oral feeding) (Supplement 1, Table 2).
6. Conscious or semi-conscious defined as a NIHSS item 1a score of 0, 1 or 2 (where the patient requires repeated stimulation to arouse).

Women of childbearing age may be included since the treatment time is short (6 daily 10 minute sessions) with no residual effects.

***22b Exclusion***

1. Non-stroke dysphagia, e.g. due to traumatic brain haemorrhage, subarachnoid haemorrhage, brain tumour, Parkinson’s disease, multiple sclerosis, severe dementia, head or neck cancer.
2. Pre-stroke dysphagia for whatever reason and whether resolved or not.
3. Pre-stroke dependency, i.e. modified Rankin scale (mRS ^44^) is 4 or 5.
4. NIHSS item 1a score of 2 (where the patient only responds to pain) or 3.
5. Ongoing or anticipated ventilation/intubation/tracheostomy. Patients who have been ventilated (and been decannulated if they required a tracheotomy) are eligible if they remain dysphagic.
6. Ongoing or planned treatment of dysphagia with other:

a. Forms of electrical / magnetic stimulation, e.g. neuromuscular electrical stimulation (NMES), transcranial direct-current stimulation (TDCS), repetitive transcranial magnetic stimulation (rTMS), Ampcare; or
b. Devices, e.g. expiratory muscle strength training (EMST), IQoro, IOPI, biofeedback using EMG electrodes, or chin tuck against resistance using a ball/chin depressor.
c. These exclusions apply for the duration of trial follow-up.
7. Malignant middle cerebral artery syndrome (although this typically presents before 4 days). Patients are eligible once stable after hemicraniectomy.
8. Pacemaker, cochlear implant or implantable cardioverter-defibrillator.
9. Need for >35% of oxygen.
10. Participants not likely to be in the treating hospital for at least 14 days (including the 6-day treatment period), i.e. expected to be repatriated to, or rehabilitated at, a non-participating organisation.
11. Two or more NGT tubes pulled out within the last week unless nasal bridle in place.
12. Investigator feels patient will not tolerate PES catheter.
13. Pregnancy if known at time of enrolment. A pregnancy test is not required.
14. Participating in another randomised controlled treatment trial for post-stroke dysphagia.
15. Palliative care.
16. Presence of a pharyngeal pouch.
17. Investigator believes dysphagia will be short-term, e.g. signs of impending recovery in swallowing (enrolment is reserved for patients whom the investigator considers are unlikely to resolve spontaneously over the next few days).
18. Participant is risk-feeding at time of screening.

**23 Recruitment**

Recruitment will be summarised in a CONSORT flow diagram showing recruitment, randomisation and follow-ups at days 14 and 90.

**24 Withdrawal/follow-ups**

***24a Withdrawals:*** Withdrawals are considered as:

- Withdrawal from treatment (applies only to PES group).
- Withdrawal from individual follow-ups.
- Withdrawal from all follow-ups.
- Missed follow-ups.
- Lost to follow-up.

***24b Timing of withdrawals:*** These will be categorised by days 14, 90, 180 and 365.

***24c Presentation of withdrawals:*** The timing of withdrawals and losses will be summarised in the CONSORT flow diagram. They will be shown by treatment group in a Kaplan-Meier graph and rates compared using Cox proportional hazards regression.

**25 Baseline patient characteristics**

***25a Listing of baseline characteristics:*** These will comprise demographics and clinical measures and are listed in *Supplement 3*.

***25b Summarisation of baseline characteristics:*** Data will be shown as number (%), median [interquartile range] or mean (standard deviation), as appropriate.

### SECTION 6. ANALYSIS

**26 Outcome definitions**

***26a Outcomes***

***Primary outcome:*** The primary outcome is the 14-level dysphagia severity rating scale (DSRS), a composite measure of swallowing impairment comprising three ordered categorical subscales each with score 0-4 so total score is 0-12 *(Supplement Table 1.1).* For PhEAST, the DSRS is extended to include death with total score = 13, as we have done previously.^15^ ^57^ Detailed guidance for scoring patients on oral trials was developed for PhEAST using the DSRS. These are: allocating a score of 11 for patients on minimal oral trials (≤15 teaspoons of any consistency in total per day) and scoring from the food and fluid sub-scales for patients on consistent oral trials (> 15 teaspoons of any consistency in total per day) alongside a score of 3 on the supervision scale. These same definitions were used to operationally define the difference between a FOIS of 2 and 3. These scores reflect the limitations of current scales to accurately capture scores for patients on oral trials which is a common approach used in the management of PSD. Although the FOIS is one of the few scales to include reference to oral trials, it was necessary to operationally define the wording used for the purpose of the study. Originally the intention was to assess the primary outcome at day 14 reflecting the design of earlier PES studies. However, post-stroke recovery paradigms value assessment of outcome at day 90 by which time effective interventions should have had a substantial effect. Additionally, PES studies find that DSRS continues to improve over 90 days whilst some recovery occurs in untreated participants. This is similar to recover trajectories of motor and speech impairments following stroke. Rather than just switching the timing of the primary outcome from day 14 to day 90, we decided to keep the first and add the second and use a repeated measures analysis; this will have the added effect of increasingly statistical power. This decision to change was made during recruitment, prior to data lock and masked to randomisation and is explained in protocol version 11 and the protocol publication.^24^

To avoid observer bias, the primary outcome measure (DSRS) is assessed by a blinded: i) speech & language therapist at day 14 while the participant is in hospital; and ii) follow-up coordinator at day 90 by telephone; both are masked to treatment assignment. Treatment catheters will be removed before the day 14 assessment and replaced by a NGT, as necessary, to maintain blinding in hospital.

All-cause death is included in the extended DSRS for multiple reasons: i) death is a common outcome after severe stroke with dysphagia, e.g. following pneumonia resulting from aspiration; ii) to avoid missing a “kill or cure” effect; iii) to avoid excluding participants from analyses (and so maintain statistical power); iv) to avoid “missingness” which may be informative; and v) to anchor analyses. Many other scales used in stroke or dysphagia include death, e.g. modified Rankin scale, Barthel index, quality of life (EQ-5D), feeding status scale.

Patients with post-stroke dysphagia may receive fluids and food in spite of speech & language therapy advice against this. Specifically, some patients may wish to receive water to drink even if they have thin liquid aspiration, so-called free water protocol.^58^ Equally, patients who are near to end-of-life may wish to feed even though they have liquid and/or food aspiration, known as “eating and drinking with acknowledged risk” (EDAR). (Of note, EDAR present at baseline is ineligible for the trial.) The® main analyses described here will use the “professional” scores derived from SLT advice. A sensitivity analysis will use “pragmatic” scores derived from what the participant is actually doing. For example, a participant may have a professional SLT-scored DSRS of 4 (fluids 1, diet 2, supervision 1) but a pragmatic DSRS of 0 (fluids 0, diet 0, supervision 0), the latter reflecting participant request. The issue of using these latter scores is that they may make the effect of control/no PES look better so that DSRS scores in the two treatment groups, PES and no PES, appear more similar.

***Secondary endpoints:*** These are listed by timing in *S Table 1.4*. Day 90/180/365 assessments will be made centrally by telephone blinded to baseline scores, treatment and in-hospital data (or by post if telephone does not work). If the participant is an inpatient at day 90, 180 or 365, the follow-up may be completed by the hospital site staff.

*Note 1:* These outcomes have all been sensitive to therapeutic change in previous studies.

*Penetration/aspiration:* The use of fibreoptic endoscopic evaluation of swallowing (FEES) or videofluoroscopy (VFS) is not mandated in the protocol but if they are carried out the penetration aspiration scale score ^59^ and date will be.recorded via the eCRF.

***Health economics:*** See *Supplement 2:* health economics analysis protocol (HEAP).

***Safety endpoints:*** PES has an excellent safety record in previous trials ^18^ whilst participants with PSD, who usually have severe stroke, will have multiple adverse events and SAEs. Hence, we will limit recording to:

- SAEs over 0-9 days.
- Procedure/device-related (S)AEs/SADEs/USADEs over days 0-14.
- Discontinuations due to (S)AEs.
- Fatal SAEs over days 10-90.

Events will be adjudicated blinded to participant information and treatment assignment.

***Device deficiencies:*** Device deficiencies will be recorded and forwarded to the company for further investigation. These will be listed as related to:

- The base station
- The treatment catheter

***Cognition:*** Post-stroke cognitive impairment (PSCI) and post-stroke dementia (PSD) follow stroke damage to cognitive neural pathways and/or the presence of concomitant small vessel disease and are common with rates up to 35% at 5 years. However, studies into PSCI/PSD are complicated by multiple issues, notably: a tendency to recruit patients with milder stroke (who are easier to enrol); assessment of cognition in the presence of existing cognitive impairment or concurrent dysphasia; difficulty in follow up, e.g. because the patient is in a care home; or the patient has died. As a result, adequate assessment of cognition and its temporal trajectory in patients with severe ischaemic stroke (IS) or intracerebral haemorrhage (ICH) is often not performed.

We expect most patients with post-stroke dysphagia in the PhEAST trial will have severe stroke, and so the PhEAST cognition study will assess cognition at days 0 (baseline), day 14 (timing of main study primary outcome), day 90 (timing of main study secondary outcomes) and days 180 and 365 after randomisation. Dysphagia measures at day 180 and day 365 will be collected as well as the cognition measures. The measures used for the cognition study will be:

- Cognition (participant): DSM-5-7 level and DSM-5-4-level ordinal cognition scales,^60^ t-MoCA, t-TICS, t-MMSE, semantic verbal fluency (animal naming), phonemic verbal fluency (letter F), clinical diagnosis of dementia (from participants or carers/informants).
- Cognition (from informant): IQCODE (separate informant consent for this).
- Dependency and disability (necessary for diagnosis of dementia): modified Rankin scale (mRS ^44^), and Barthel index (BI).
- Frailty: clinical frailty index (CFI).
- Mood/depression (which complicates PSCI/PSD diagnosis): Zung depression scale.
- Health economics: EQ-5D-5L, EQ-VAS.
- Stroke impact scale.
- Recurrent stroke after index event.

Overall, the cognition study will provide information on cognition and its trajectory over the first year after severe stroke, a neglected research area and of considerable importance to this population and their family and carers. Additionally, it will enhance the main trial itself through providing extended follow-up information on swallowing.

***26b Units:*** Units will be shown in tables.

***26c Calculations/transformations:*** Quality of life (EQ-5D-5L) using UK weightings.

***26d NICE:*** NICE list several key efficacy and safety outcomes to be assessed in research:^18^

- *“Degree of aspiration“* - we assess that as chest infection or pneumonia.
- *“Change in severity of dysphagia“* - we assess that as swallow impairment (DSRS).
- “*Need for nasogastric and percutaneous endoscopic gastrostomy or jejunostomy feeding” - we assess* that as need for any enteral tube (whether NGT, PEG, RIG, J-tube) and need for any permanent feeding tube (whether PEG, RIG, J-tube).
- “Device-related discomfort or injury” - we assess these through reporting of SADE, USADE or device deficiencies.
- **27 Analysis methods**

***27a Methods of analyses:*** Results will be tabulated as number (%), median [interquartile range] or mean (standard deviation) (*S Table 1.5*). Absolute (binary analyses) and relative measures of effect and 95% confidence intervals will be presented for each analysis. In tabulations, a worst score will be assigned at days 14, 90, 180 and 365 for people who die (e.g. DSRS=13, FOIS=0, mRS=6) to avoid losing participants in analyses (and so reduce statistical power), missing a “kill or cure” effect and to anchor analyses.

**S Table 1.5:**
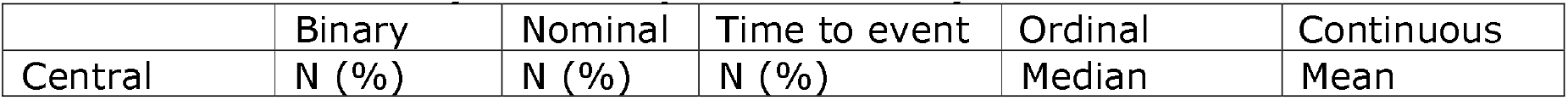
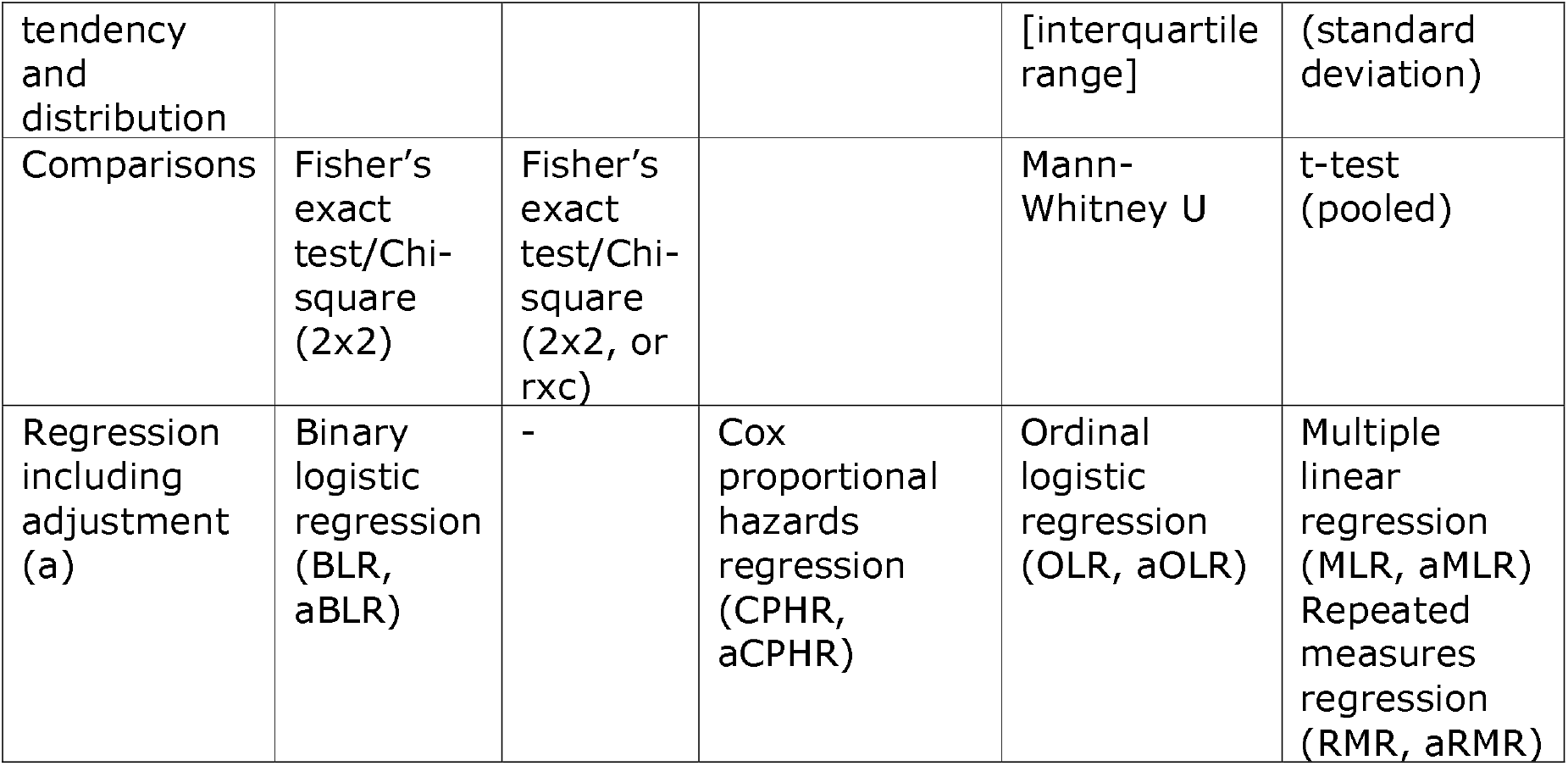
Summary of descriptive and analytical statistics.

***Analyses of Primary endpoint:*** DSRS will be analysed by adjusted repeated measures linear mixed effects regression based on data from days 14 and 90 with covariate adjustment. The model will include fixed effects for treatment group, time (categorical), and the treatment-by-time interaction, with baseline outcome value included as a covariate. A random intercept for participant will be included to account for within-subject correlation. An unstructured covariance matrix will be used to model the repeated measurements; alternative covariance structures will be explored in sensitivity analyses. Effect size will be reported as the average treatment effect with confidence intervals and p-value. The interaction between treatment effect and time will also be reported.

***Analyses of secondary outcomes:*** Secondary outcomes will be compared using continuous repeated measures regression, ordinal repeated measures regression (e.g. FOIS), multiple linear regression (e.g. Barthel index), ordinal logistic regression (e.g. mRS), Cox proportional hazards regression (e.g. time to death) and binary logistic regression (e.g. discharge with PEG/RIG).

***27b Covariate adjustment:*** Analyses will be adjusted for baseline DSRS, age, sex, impairment (NIHSS), stroke type (ischaemic/haemorrhagic), circulation (anterior/posterior) and time to randomisation (days). Non-binary variables will use raw data.

***27c Assumption checking:*** Since the sample size is considered to be large, we will not test for normality, however plots of the residuals will be reviewed to check for potential violations.. The assumption of proportionality for ordinal logistic regression will be tested with the likelihood test and reported.

***27d Alternative methods:*** We will still use OLR if the test of proportionality fails reporting odds ratio and confidence intervals.

***27e Sensitivity analyses:*** The primary outcome of DSRS will be subject to sensitivity analyses as shown in ***S Table 6***. The analyses will assess the impact of imputation of missing DSRS scores at day 14 and/or 90, not adjusting for minimisation and other important baseline variables, only including participants where no key protocol violations occurred (per protocol), accounting for free water administration or EDAR assessing the outcome only at day 14 or day 90, and excluding death.

**S Table 6.** Sensitivity analyses for the primary outcome, DSRS.

| Primary outcome | Day | Population | Imputation | Analysis |
| --- | --- | --- | --- | --- |
| With no imputation | 14/90 | mITT | No | aRMR |
| *Sensitivity analyses* |  |  |  |  |
| With imputation | 14/90 | ITT | Yes | aRMR |
| Unadjusted | 14/90 | mITT | No | RMR |
| Per protocol (#20.4) | 14/90 | PP | No | aRMR |
| Accounting for FWP/EDAR | 14/90 | mITT | No | aRMR |
| With no imputation | 14 | mITT | No | aMLR |
| With no imputation, change | 14 | mITT | No | aMLR |
| With no imputation | 90 | mITT | No | aMLR |
| With no imputation, change | 90 | mITT | No | aMLR |
| Excluding death | 14/90 | mITT | No | aRMR |

a: adjusted; EDAR: eating and drinking with acknowledged risk; FWP: free water protocol; ITT: intention-to-treat; mITT: modified intention-to-treat; PP: per protocol

***27f Subgroup analyses :*** The primary outcome, DSRS, will be assessed in pre-specified subgroups based on minimisation factors using interaction tests to identify responder and non-responder subgroups. Between subgroup effect (95% CI) will be presented:

- Age (years): <75, > = 75
- Sex: female, male
- Reperfusion: yes, no
- Dysphagia severity rating scale (/12): <11, 11, 12
- Functional oral intake scale (/7): 1, 2/3
- Stroke severity, NIHSS (/42): <13, > = 13
- Stroke type: ischaemic, primary intracerebral haemorrhage
- Circulation: anterior, posterior
- Onset to randomisation (days): <12, 12-19, >19
- Lesion side: right, bilateral, left
- Stroke size: small, medium, large
- White matter disease: none, mild, moderate/severe
- Prior instrumental testing: yes, no
- Responder, DSRS change day 90-0: >-3.5, <=-3.5
- Responder, DSRS day 90: <4, >=4
- Experience with PES prior to trial: yes, no
- Site recruitment: >10, < = 10

Since the trial is powered to detect overall differences between groups rather than subgroup interactions, these analyses will be regarded as hypothesis generating. The importance of some of these subgroup analyses is highlighted by similar findings in PHAST-TRAC and PHADER that PES appears to more effective if treatment is started within the first month after stroke.^16,20^

A subgroup analysis will be performed in the PES-only group to assess the relationship between stimulation levels (in milliamps) and effect on primary outcome at day 90. The analysis will follow that of PHADER.^16^

- Threshold, average: <10, 10-13, >13
- Stimulation, average: <30, 30-36, >36
- Tolerability, average: <35, 35-45, >45
- Delta, tolerability - threshold, average: <24, 24-33, >33

**28 Missing data:** Missing data will be imputed using multiple regression imputation for a sensitivity analysis of DSRS. The multiple imputation will be undertaken using the PROC MI procedure in SAS, with 20 burn-in iterations and using the default setting for number of imputations. Data will be imputed at days 14 and/or 90 if missing. If participants have definitely died, they will be assigned a value of DSSR=13; if they are definitely alive but their DSSR is unknown, they will be assigned a DSRS value of 0-12 as determined by regression imputation.

***29 Additional analyses:*** No further analyses planned.

***30 Harms:*** These will be presented in Tables describing serious adverse events.

***31 Statistical software:*** Statistical Analysis System (SAS) version 9.4 (or later), SAS Institute Incorporation, Cary, North Carolina.

### SECTION 7. ADDITIONAL INFORMATION

**32 Governance:** The trial is funded by the National Institute for Health and Care Research (NIHR) Health Technology Assessment (HTA) Programme and sponsored by the University of Nottingham. Phagenesis Ltd, the device manufacturer, lend sites a base station and provide catheters and training for free. The trial is managed by a Trial Management Committee (TMC), supervised by a Trial Steering Committee (TSC) and overseen by an independent Data Monitoring Committee (DMC).

**33 Minimising bias**

Multiple approaches are taken in the design and execution of the trial to minimise bias:

- Central data registration with real-time on-line validation;
- Concealment of allocation;
- Adaptive randomisation/minimisation;
- Inclusion of patients enrolled in other studies (co-enrolment) where feasible;
- Blinded assessment of DSRS at day 14;
- Blinded central telephone (or postal) assessment of outcomes at days 90, 180, 365;
- Analysis by modified intention-to-treat and in pre-specified subgroups.

**34 Study within a trial (SWAT)**

The SWAT will investigate the effects of a package to improve awareness and communication with investigators to improve PES delivery and especially delivered current. Sites are cluster-randomised to the intervention vs control at ratio 1:1. The intervention comprises the Coordinating Centre’s speech & language therapist contacting the treater who under-treated the participant and determining what happened and any root causes. The treater is reminded of the importance of providing adequate treatment in respect of stimulation current and treatment duration.

One interim analysis is planned at 200 PES participants (i.e. 400 total randomised) in the main trial. If there is strong evidence of an effect of the SWAT intervention on treatment current, the strategy showing the greatest treatment current would then be implemented for the remaining participants. Otherwise, the SWAT will continue until the end of the trial. Analyses will include descriptive statistics and between-group comparisons for each strategy using multivariate regression models.

*Success criteria:*

1. Increased treatment current over first three days in participants randomised to SWAT vs no-SWAT.
2. Increased proportion of participants with mean treatment current >20 mA in hospitals randomised to SWAT vs no-SWAT.
3. Increased proportion of participants who only require one PES catheter in hospitals randomised to SWAT vs no-SWAT.

**35 Discrepancies between protocol and statistical analysis plan**

Where there is a difference between the protocol (website and published versions) and this statistical analysis plan (SAP), the SAP takes precedence. This is particularly relevant to the change in analysis of the primary outcome.

**36 Calculations/definitions**

***36a FOIS:*** This is a swallow impairment scale

| Level | Item | Notes |
| --- | --- | --- |
| 1 | Nothing by mouth |  |
| 2 | Tube dependent with minimal attempts of food or liquid | Minimal is defined as ≤15 tea-spoons in total of any consistency per day |
| 3 | Tube dependent with consistent oral intake of food or liquid | Consistent is defined as >15 tea-spoons of any consistency per day in total (but not sufficient for nutritional purposes, i.e. still requires some element of tube feeding) |
| 4 | Total oral diet of a single consistency |  |
| 5 | Total oral diet with multiple consistencies, but requiring special preparation or compensations |  |
| 6 | Total oral diet with multiple consistencies without special preparation, but with specific food limitations |  |
| 7 | Total oral diet with no restrictions |  |

***36b FSS:*** The FSS assesses feeding status based on mode of nutrition. The scale is shown here with the scores as coded incorrectly in the REDCap database and the correct scores, as published,^48^ which will be reported in publications.

| Correct score | Score in REDCap | Level | Item (adapted from <sup>48</sup>) |
| --- | --- | --- | --- |
| 0 | 1 | Oral: Normal | Regular oral diet (IDDSI L7/L7 easy chew).<br>Normal oral fluids (IDDSI L0).<br>Non-oral feeding route may be in situ but is not required to maintain nutrition. |
| 1 | 2 | Oral: Soft | Modified oral diet (IDDSI L6 or below).<br>Oral fluids – normal or modified (IDDSI L0-L4).<br>Non-oral feeding route may be in situ but is not required to maintain nutrition. |
| 2 | 3 | Non-oral: NGT | Nasogastric tube feeding required to<br>maintain nutrition.<br>• May receive oral fluids – normal or modified (IDDSI L0-L4).<br>• May receive tastes/oral trials of diet. |
| 3 | 4 | Non-oral:<br>PEG/RIG/J-tube | • Percutaneous endoscopic gastrostomy feeding (or equivalent) required to maintain nutrition.<br>• May receive oral fluids – normal or modified (IDDSI L0-L4).<br>• May receive tastes/oral trials of diet. |
| 4 | 5 | Non-oral: IV/SC | • Non-oral supplemented fluid intake.<br>• May receive oral fluids – normal or modified (IDDSI L0-L4).<br>• May receive tastes/oral trials of diet. |
| 5 | 6 | Non-oral:<br>nothing | • May receive tastes/free water protocol for comfort.<br>• May be awaiting placement of PEG/NGT. |
| 6 | 0 | Died | • Due to any cause |

***36c IDDSI-FDS:*** Two IDDSI scores rate swallow impairment on the basis of intake of:^49^

- Food : regular (i.e. normal) score =7; soft & bite sized =6; minced & moist =5; pureed =4; liquidised =3; nil by mouth = 2.
- Drinks : thin (i.e. normal) =0; slightly thick =1; mildly thick =2; moderately thick = 3; extremely thick, score =4; nil by mouth =5.

From this, the overall swallow impairment is calculated using the IDDSI functional diet scale (IDDSI-FDS) score which ranges from 1 to 8.^50^ We have modified the IDDSI-FDS score to include values less than 1 to allow scoring of nil by mouth, including minimal and consistent trials. Hence, the modified IDDSI-FDS (m-IDDSI-FDS) ranges from 0 (nil-by-mouth) to 8 (food 7, drinks 0, i.e. normal). Score of 0, 0.25, 0.5 and 0.75 allow differentiation of patients who are nil-by-mouth but may or not be taking trials of feeding:

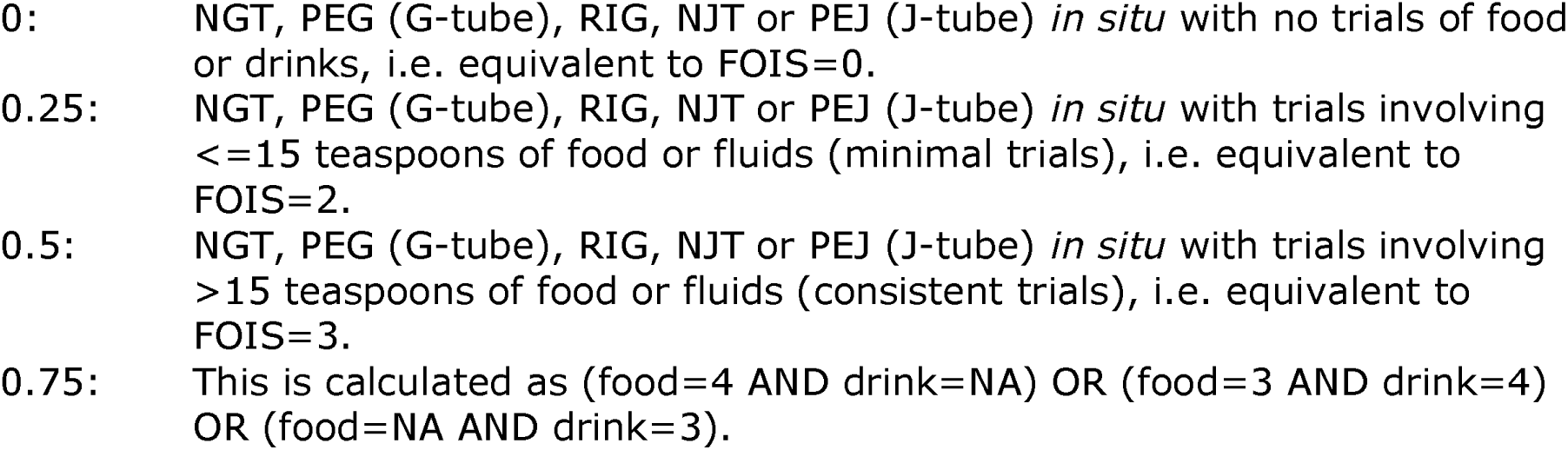

The modification maintains symmetry around the top-right / bottom-left axis.

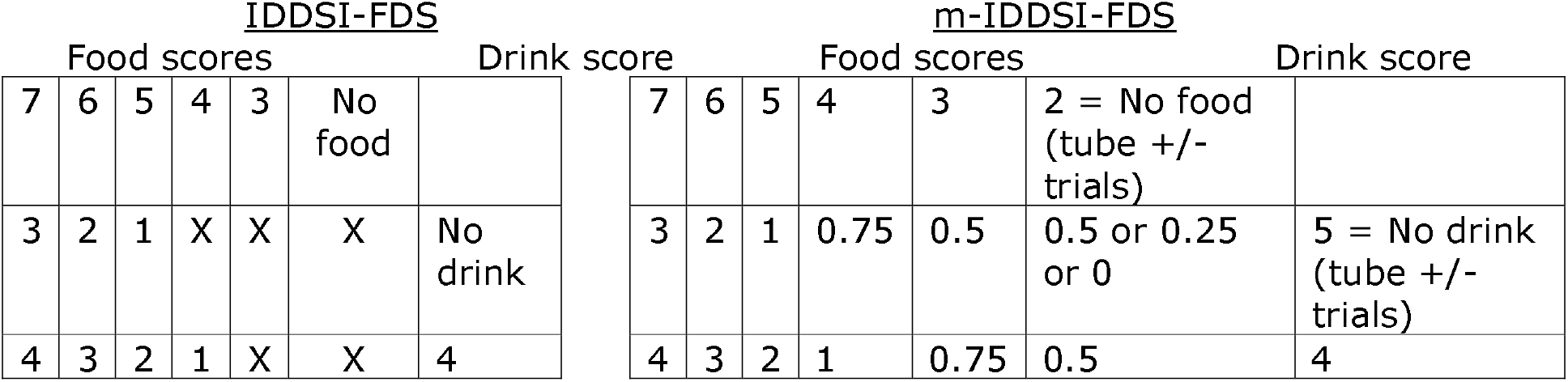
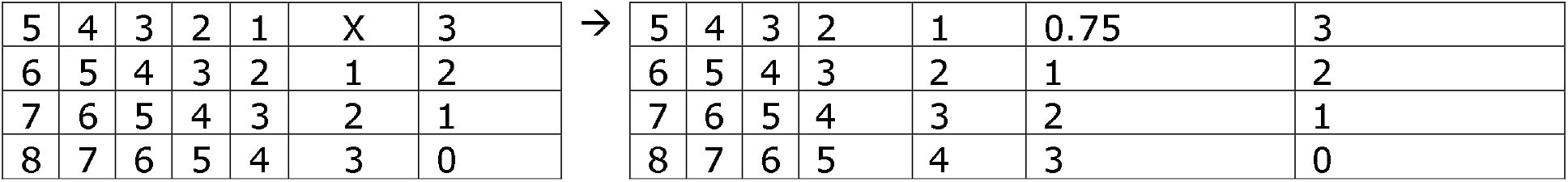

| IDDSI-FDS |  |  |  |  |  |  | m-IDDSI-FDS |  |  |  |  |  |  |
| --- | --- | --- | --- | --- | --- | --- | --- | --- | --- | --- | --- | --- | --- |
| Food scores |  |  |  |  |  | Drink score | Food scores |  |  |  |  |  | Drink score |
| 7 | 6 | 5 | 4 | 3 | No food |  | 7 | 6 | 5 | 4 | 3 | 2 = No food (tube +/- trials) |  |
| 3 | 2 | 1 | X | X | X | No drink | 3 | 2 | 1 | 0.75 | 0.5 | 0.5 or 0.25 or 0 | 5 = No drink (tube +/- trials) |
| 4 | 3 | 2 | 1 | X | X | 4 | 4 | 3 | 2 | 1 | 0.75 | 0.5 | 4 |

|  |  |  |  |  |  |  |  |  |  |  |  |  |  |  |
| --- | --- | --- | --- | --- | --- | --- | --- | --- | --- | --- | --- | --- | --- | --- |
| 5 | 4 | 3 | 2 | 1 | X | 3 | → | 5 | 4 | 3 | 2 | 1 | 0.75 | 3 |
| 6 | 5 | 4 | 3 | 2 | 1 | 2 |  | 6 | 5 | 4 | 3 | 2 | 1 | 2 |
| 7 | 6 | 5 | 4 | 3 | 2 | 1 |  | 7 | 6 | 5 | 4 | 3 | 2 | 1 |
| 8 | 7 | 6 | 5 | 4 | 3 | 0 |  | 8 | 7 | 6 | 5 | 4 | 3 | 0 |

***36d EAT-10:***^51^ This 10-item scale assesses swallowing difficulties when swallowing is still possible. Each item scores from 0 = no problem to 4 = severe problem so that the maximum score is 10 x 4 = 40. Since the scale does not account for non-oral feeding, we have extended it by adding a nil by mouth score = 5 resulting in a maximum score of 50. This score includes participants who are on oral trials.

***36e Death scores:*** Several scales already include a value for death, e.g. mRS=6, BI=-5 and EQ-5D health utility = 0. We have extended other scales to include death, usually with a value more extreme than any living value. The aim is to avoid: losing participants in analyses (and so maintain statistical power), missing a “kill or cure” effect and to anchor analyses.^36,38,39,46^ The death values are given here:

- Swallowing/eating: DSRS=13, FOIS-O, FSS=6, IDDSI-food=1, IDDSI-drink=6, IDDSI-FDS=-1, EAT-10 = 51
- Others: CFS=10, EQ-VAS=-1, NIHSS=43, ZDS=102.5

**36f Global analyses:** Analyses that include a global assessment of multiple scales are usually more efficient statistically. We will use the Wei-Lachin test to perform these for two groupings of scales:

- Swallowing/feeding: DSRS, FOIS, FSS, IDDSI-FDS and EAT-10.
- Stroke impact scale:^52^ 8 domains - strength, hand function, mobility, activities of daily living, memory, communication, handicap.

**36d *Composite of poor outcomes at death or discharge:*** DSRS>6, pneumonia in hospital, PEG ***in situ*** at discharge, died in hospital or length of stay >upper interquartile range in days.

**37 Publications, published and planned**

1. Protocol: On trial website and published.^24^
2. Statistical Analysis Plan (SAP) and Health economics analysis plan: On trial website and this publication.
3. Baseline characteristics.
4. Primary results using data to day 90.
5. Health economics using data to day 90.
6. Prediction of swallowing recovery including responder analysis.
7. Pharyngeal electrical stimulation: experience, safety and device deficiencies.
8. Update to published Cochrane Collaboration systematic review.
9. Update to published individual patient data meta-analysis.^14^
10. Design learnings, including training, weekends, consent, challenges, free water, eating and drinking with acknowledged risks (EDAR), dysphasia, co-enrolment.
11. Validation of PRESS score.^61^
12. Primary swallowing/dysphagia results using data to day 360.
13. Health economics using data to day 360.
14. Cognition and mood trajectory using data to day 360.
15. Prevalence and use of oral trials.
16. Qualitative assessment of PES delivery.
17. Calculating swallowing and eating scales from clinical information.

**38 Data sharing**

Data from PhEAST will be added to the Cochrane Collaboration review of PSD ^4^ and integrated into an updated individual patient data meta-analysis of PES.^62^ Individual patient data will be shared with the ‘Virtual International Stroke Trials Archive’ (VISTA) ^63^ and, ultimately, made available over the web.^34^

### SUPPLEMENT 2

**Health Economic Analysis Plan (HEAP) for the ‘Pharyngeal electrical stimulation for acute stroke dysphagia trial’ (PhEAST)**

Version 5 26.01.2026

Authors: Marilyn James, Cristina Roadevin. Nottingham Clinical Trials Unit

**Signatures**

| **Role** | **Name** | **Signature** | **Date** |
| --- | --- | --- | --- |
| ***Author, Professor of Health Economics*** | **Prof Marilyn James** |  |  |
| ***Author, Health Economist*** | **Dr Cristina Roadevin** |  |  |

**1. Purpose of HEAP**

The aim of this HEAP is to outline the design and analytical methods used to evaluate the cost effectiveness of the PhEAST trial intervention compared to usual care. The methods specified are prospective and an indicative guide for the analysis but are subject to change in line with recommendations of official bodies and/or guidelines of best practice. Similarly, completion rates of health economic measures within the trial may require further assumptions or analyses.

**2. Economic Perspective**

The primary analysis will adopt an NHS and personal social services perspective in accordance with NICE guidance (2022). Parallel analysis will take a broader societal perspective.

**3. Economic Data and Management**

***3.1 Software***

Data will be imported from Microsoft Excel and analysed in StataSE (Release 18.5; StataCorp USA).

***3.2 Data Cleaning***

Plausibility checks will be performed on data fields relevant to the economic evaluation. Any identified issues will be clarified with the data manager or relevant staff.

***3.3 Outcomes***

Primary outcome (Health economics)

- Health-related quality of life (HRQoL) for participant: EuroQoL-5D-5L (EQ-5D-5L) administered at baseline and days: 14 and 90. We hope to later extend this to days 180 and 365.
- ***3.4 Derivation of Indices of HRQoL and Quality Adjusted Life Years***

HRQoL indices will be derived using a relevant population tariff. If unavailable for EQ-5D-5L, responses will be mapped to EQ-5D-3L for analysis. Area under the curve (AUC) will be used to adjust participant HRQoL for the time spent in their respective health states, constructing quality adjusted life-years (QALYs).

***3.5 Health Care and Other Resource Use***

*3.5.1 Patient level resource use*

Patient level resource use data will be collected using a purposely designed patient resource proforma. This will collect health care and out of pocket and patient level data such as medication, hospital visits, rehabilitation, primary care, aids and appliances, social care and time lost from paid employment for the patient or their relatives/carers. The proforma will be used to collect data at day 90. The same is used at day 180 and 365 and these will be used potentially in a secondary analysis.

*3.5.2 Potential intervention-specific costs*

1. PES Equipment Costs:

- Base Station: The durable component of the PES system, which generates, optimises and monitors the delivery of electrical stimulation.
- Catheter: A single-use sterile disposable component. It incorporates electrodes and wiring for delivering electrical stimulation and also functions as a feeding tube. Each patient may require additional catheters to account for potential dislodgement and the need for replacements.

2. Staff Costs:

- Training: Costs associated with training Nurses and Speech and Language Therapists to administer PES.
- Staff type and grade delivering PES: Costs for the time spent by trained staff to administer PES treatments. This includes six daily 10-minute sessions per patient.
- Monitoring and Support: Costs for ongoing monitoring and support, including troubleshooting and ensuring the correct administration of PES.

3. Swallowing Therapy:

Costs related to swallowing therapy contact time (SLT) which might be impacted by the effectiveness of PES.

4. PEG/RIG tube use

The cost of the PEG/RIG tube itself, including any ancillary supplies required for its placement and maintenance. The cost of tube placement, typically involving an endoscopic (or radiologic) procedure performed by a gastroenterologist. Ongoing costs for maintaining the tube, including supplies for cleaning and feeding, and any additional procedures required to manage complications (e.g., bleeding, infections, displacement). Nursing, dietician and other healthcare professional time required to care for patients with tubes, including feeding and managing the tube.

5. Diet costs

Costs related to specialised diets, thickening powders and nutritional supplements provided to patients with dysphagia. Includes the cost of preparing and delivering modified food and thickened liquid consistencies to ensure safe swallowing. For participants who are receiving food via a tube, includes costs of enteral feeding sets

**3.6 Costing of Resource Use**

Costs will be adjusted to current values using inflation indices from PSSRU.^64^

**Table S2-1.** Costing sources.

|  |  |
| --- | --- |
| NHS inpatient admissions | The National Cost Collection for the NHS (https://www.england.nhs.uk/costing-in-the-nhs/national-cost-collection/) |
| NHS outpatient visits, primary/community services, residential care, nursing care | Unit Costs of Health and Social Care (https://www.pssru.ac.uk/).<sup>64</sup> |
| Medication | British National Formulary |
|  | (https://bnf.nice.org.uk/) |
| Special equipment | https://www.essentialaids.com/mobility/walking-frames-accessories.html |
| Informal care | The costs of absenteeism will be estimated through the lost wages approach. Reports of health-related time taken off work to help with patient's care will be combined with population-level gross weekly salaries from the Annual Survey of Hours and Earnings (Office for National Statistics, 2024). |

**Table S2-2.** Costing sources, direct intervention costs.

|  |  |
| --- | --- |
| PES Equipment Costs | Company (Phagenyx®, Phagenesis Ltd) |
| Staff costs(training, administration, monitoring) | Company (Phagenyx®, Phagenesis Ltd) |
| Speech and Language therapy | Unit Costs of Health and Social Care <sup>64</sup> (https://www.pssru.ac.uk). The National Cost Collection for the NHS Duration from sites |
| PEG/RIG tube | The National Cost Collection for the NHS |
| Diet costs | Sites/Pharma |

We will apply the following assumption when costing the ‘other’ category, where patients have provided free-text responses: the average cost of items within that category will be imputed. For example, if a drug is listed under ‘other’, we will assign it the average cost of all medications.

**4. Procedures for Missing Data**

Handling of missing data will follow guidelines for intention-to-treat analysis with incomplete observations.^65^ Multiple imputation methods will be applied using available case data, such as Multiple Imputation of Chained Equations (MICE), which build into their models the inherent uncertainty associated with the missing data.^66^

**5. Within-trial Analysis**

***5.1 Population and Time Horizon***

The economic evaluation will use an incremental approach between the two groups with an intention-to-treat (ITT) population and a 90-day time horizon. We hope to extend this to a 365-day horizon subject to funding.

***5.2 Discount Rates***

No discounting will be applied to derived QALYs or costs within the 3 month period.

***5.3 Analysis of Outcomes and Resource Use***

The economic evaluation will adopt both a healthcare perspective (NHS and Personal Social Services) and a broader societal perspective. Costs, QALYs, and cost-effectiveness estimates will be derived using multiply imputed datasets to address missing data and improve analytical robustness.

Cost-effectiveness will be assessed through incremental cost-effectiveness ratios (ICERs) and incremental net monetary benefits (INMBs). In the base-case analysis, differences in costs and outcomes between trial arms will be estimated using seemingly unrelated regression (SUR) models applied to the imputed datasets. This method enables the joint estimation of cost and QALY equations, allowing for correlation between their error terms while adjusting for baseline characteristics and other relevant covariates.

Regression models will estimate treatment-specific mean costs and QALYs, adjusting for age, sex, and other potentially important trial variables. To account for baseline differences in health-related quality of life, QALY models will also include baseline EQ-5D preference scores as a covariate.

In addition to the cost-utility analysis, we will perform a cost effectiveness analysis focusing on the trial’s primary outcome, the Dysphagia Severity Rating Scale (DSRS), provided that the outcome favours the intervention and is statistically significant. Using the clinical outcome analysis provided by the statistics team, we will calculate incremental cost effectiveness ratios (ICERs) by dividing the mean difference in costs by the mean difference in DSRS scores between the PES and no-PES group. If the clinical outcome favours standard care, a cost effectiveness analysis will not be undertaken, as the intervention would be dominated by standard care and the calculation of an ICER would not be informative for decision making.

***5.4 Sensitivity Analyses***

Uncertainty surrounding the estimated treatment effects will be addressed through probabilistic sensitivity analysis (PSA). This will involve simulating the joint distribution of regression coefficients using 10,000 Monte Carlo iterations. This approach will enable us to quantify how uncertainty in both costs and outcomes translates into the likelihood of overall cost effectiveness.

Cost Effectiveness Acceptability Curves (CEACs) will be generated using the net monetary benefit framework, which expresses the value to the NHS of the intervention for a given willingness to pay threshold (λ) for the outcome of interest. The incremental net monetary benefit is defined as:

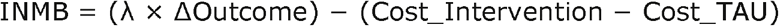

Under this framework, the decision rule is that an intervention is considered cost effective when the incremental net monetary benefit is greater than zero, which is equivalent to the incremental cost effectiveness ratio being below the willingness to pay threshold. Using joint uncertainty in costs and outcomes derived from the SUR based probabilistic sensitivity analysis, we will calculate the INMB across a range of willingness to pay thresholds for each simulation. The CEAC will then show the probability that the intervention is cost effective at different willingness to pay thresholds per QALY. In addition, sensitivity analysis will be used to explore key cost drivers if necessary.

**6. Reporting/Publishing**

***6.1 Reporting Standards***

The final report will be submitted for peer review alongside a CHEERS checklist.

***6.2 Reporting Deviations from the HEAP***

Any deviations from the HEAP will be reported in full and communicated with the study team.

**7. References**

National Institute for Health and Care Excellence. NICE health technology evaluations: the manual 2022. https://www.nice.org.uk/process/pmg36/chapter/introduction-to-health-technology-evaluation

Office for National Statistics. Dataset: Consumer price inflation tables. 2024 (https://www.ons.gov.uk/economy/inflationandpriceindices/datasets/consumerpriceinflation)

Office for National Statistics. Dataset: Earnings and hours worked, all employees: ASHE Table 1. 2024 (https://www.ons.gov.uk/employmentandlabourmarket/peopleinwork/earninqsandworkinqhours/datasets/allemployeesashetable1)

Example tabulated results at 12 months are shown in Supplement 5.

### SUPPLEMENT 3

**PHARYNGEAL ELECTRICAL STIMULATION FOR ACUTE/SUB-ACUTE POST-STROKE DYSPHAGIA: MAIN RESULTS FROM THE PHEAST TRIAL**

**1. DESCRIPTION OF MAIN PAPER**

**Title**

Pharyngeal electrical stimulation for acute post-stroke dysphagia: main results from the PhEAST trial

**Authors**

PhEAST Investigators

**Aim**

Describe the primary outcome and key secondary outcomes.

**Participants**

All in PhEAST.

**Outcomes**

*Primary outcome, days 14/90:* DSRS.

*Secondary outcomes, days 14/90:*

*Safety, day 14/90:* serious adverse events, serious adverse device effects, unexpected serious adverse device effects, device deficiencies.

**Analyses**

Descriptive statistics: number (%), median [interquartile range] or mean (standard deviation). Comparisons using repeated measures regression, multiple linear regression, ordinal logistic regression, Cox proportional hazards regression, binary logistic regression and Wei-Lachin test.

**Table S3-1.**
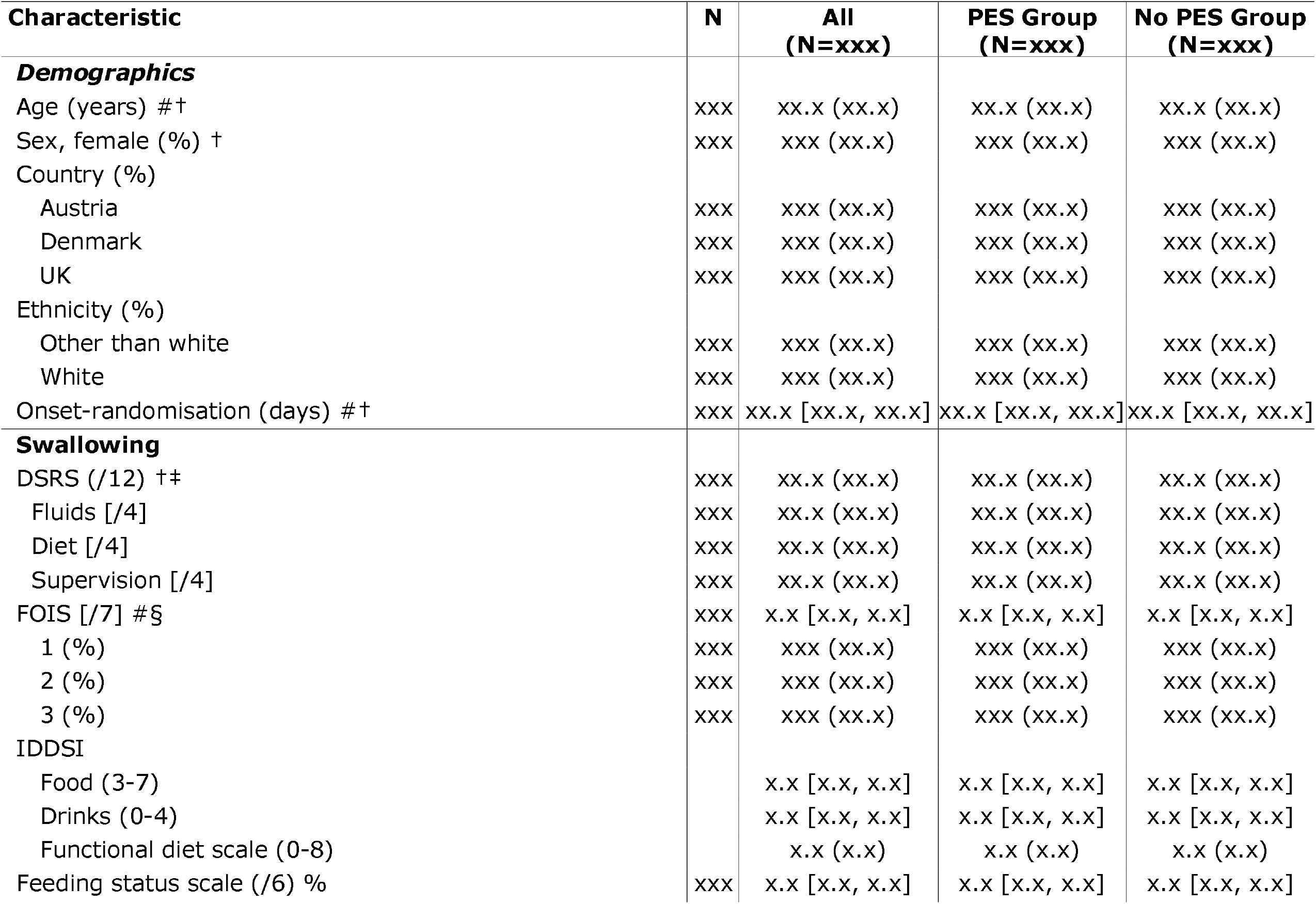
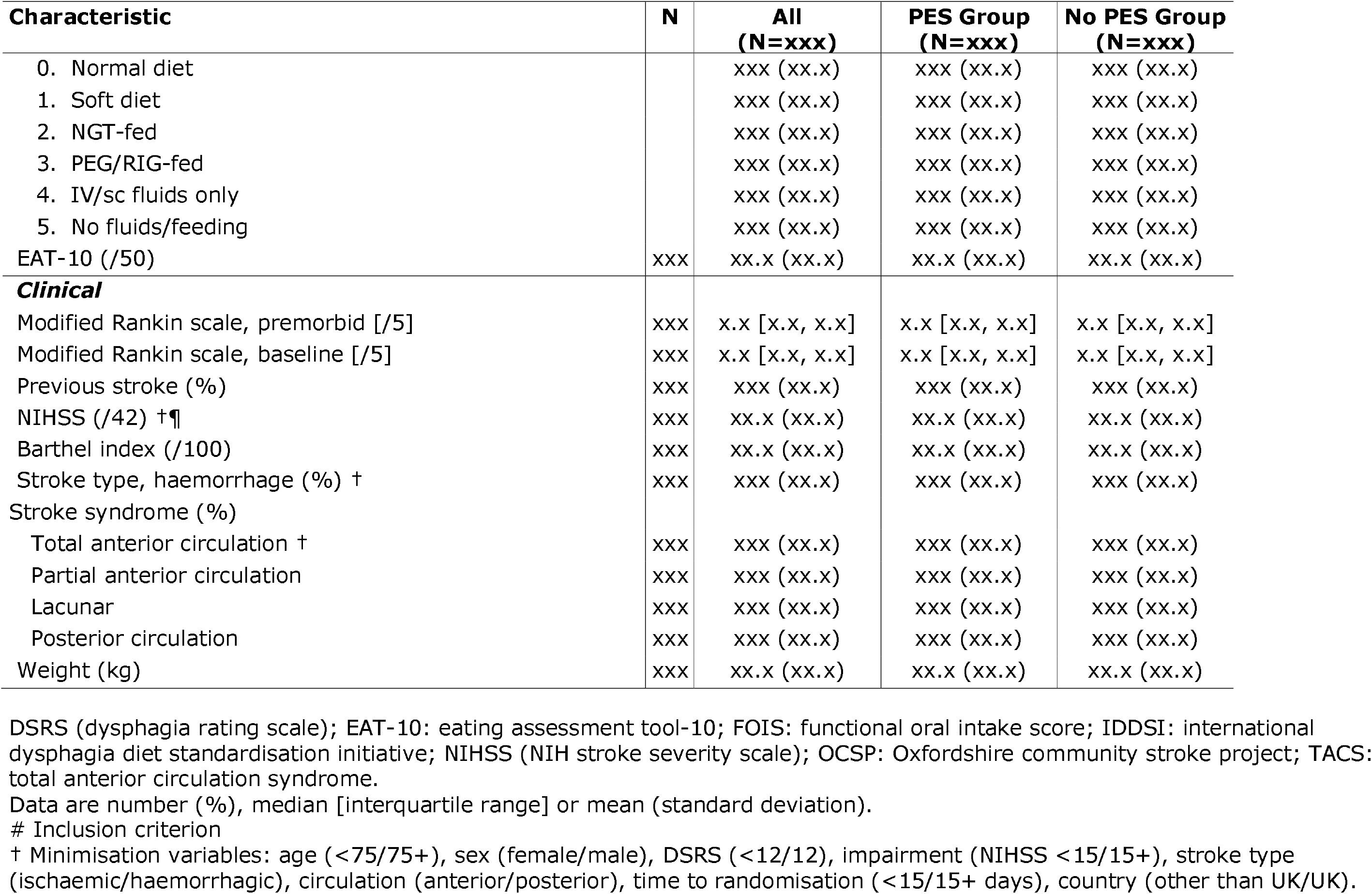
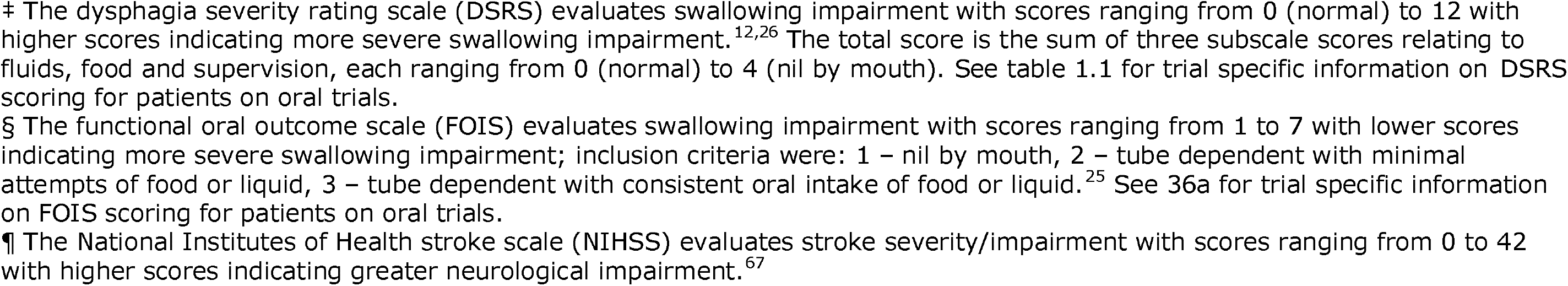
Characteristics of the patients at baseline.

| Characteristic | N | All<br>(N=xxx) | PES Group<br>(N=xxx) | No PES Group<br>(N=xxx) |
| --- | --- | --- | --- | --- |
| **Demographics** |  |  |  |  |
| Age (years) #† | xxx | xx.x (xx.x) | xx.x (xx.x) | xx.x (xx.x) |
| Sex, female (%) † | xxx | xxx (xx.x) | xxx (xx.x) | xxx (xx.x) |
| Country (%) |  |  |  |  |
| Austria | xxx | xxx (xx.x) | xxx (xx.x) | xxx (xx.x) |
| Denmark | xxx | xxx (xx.x) | xxx (xx.x) | xxx (xx.x) |
| UK | xxx | xxx (xx.x) | xxx (xx.x) | xxx (xx.x) |
| Ethnicity (%) |  |  |  |  |
| Other than white | xxx | xxx (xx.x) | xxx (xx.x) | xxx (xx.x) |
| White | xxx | xxx (xx.x) | xxx (xx.x) | xxx (xx.x) |
| Onset-randomisation (days) #† | xxx | xx.x [xx.x, xx.x] | xx.x [xx.x, xx.x] | xx.x [xx.x, xx.x] |
| **Swallowing** |  |  |  |  |
| DSRS (/12) †‡ | xxx | xx.x (xx.x) | xx.x (xx.x) | xx.x (xx.x) |
| Fluids [/4] | xxx | xx.x (xx.x) | xx.x (xx.x) | xx.x (xx.x) |
| Diet [/4] | xxx | xx.x (xx.x) | xx.x (xx.x) | xx.x (xx.x) |
| Supervision [/4] | xxx | xx.x (xx.x) | xx.x (xx.x) | xx.x (xx.x) |
| FOIS [/7] #§ | xxx | x.x [x.x, x.x] | x.x [x.x, x.x] | x.x [x.x, x.x] |
| 1 (%) | xxx | xxx (xx.x) | xxx (xx.x) | xxx (xx.x) |
| 2 (%) | xxx | xxx (xx.x) | xxx (xx.x) | xxx (xx.x) |
| 3 (%) | xxx | xxx (xx.x) | xxx (xx.x) | xxx (xx.x) |
| IDDSI |  |  |  |  |
| Food (3-7) |  | x.x [x.x, x.x] | x.x [x.x, x.x] | x.x [x.x, x.x] |
| Drinks (0-4) |  | x.x [x.x, x.x] | x.x [x.x, x.x] | x.x [x.x, x.x] |
| Functional diet scale (0-8) |  | x.x (x.x) | x.x (x.x) | x.x (x.x) |
| Feeding status scale (/6) % | xxx | x.x [x.x, x.x] | x.x [x.x, x.x] | x.x [x.x, x.x] |
| 0. Normal diet |  | xxx (xx.x) | xxx (xx.x) | xxx (xx.x) |
| 1. Soft diet |  | xxx (xx.x) | xxx (xx.x) | xxx (xx.x) |
| 2. NGT-fed |  | xxx (xx.x) | xxx (xx.x) | xxx (xx.x) |
| 3. PEG/RIG-fed |  | xxx (xx.x) | xxx (xx.x) | xxx (xx.x) |
| 4. IV/sc fluids only |  | xxx (xx.x) | xxx (xx.x) | xxx (xx.x) |
| 5. No fluids/feeding |  | xxx (xx.x) | xxx (xx.x) | xxx (xx.x) |
| EAT-10 (/50) | xxx | xx.x (xx.x) | xx.x (xx.x) | xx.x (xx.x) |
| **Clinical** |  |  |  |  |
| Modified Rankin scale, premorbid [/5] | xxx | x.x [x.x, x.x] | x.x [x.x, x.x] | x.x [x.x, x.x] |
| Modified Rankin scale, baseline [/5] | xxx | x.x [x.x, x.x] | x.x [x.x, x.x] | x.x [x.x, x.x] |
| Previous stroke (%) | xxx | xxx (xx.x) | xxx (xx.x) | xxx (xx.x) |
| NIHSS (/42) †¶ | xxx | xx.x (xx.x) | xx.x (xx.x) | xx.x (xx.x) |
| Barthel index (/100) | xxx | xx.x (xx.x) | xx.x (xx.x) | xx.x (xx.x) |
| Stroke type, haemorrhage (%) † | xxx | xxx (xx.x) | xxx (xx.x) | xxx (xx.x) |
| Stroke syndrome (%) |  |  |  |  |
| Total anterior circulation † | xxx | xxx (xx.x) | xxx (xx.x) | xxx (xx.x) |
| Partial anterior circulation | xxx | xxx (xx.x) | xxx (xx.x) | xxx (xx.x) |
| Lacunar | xxx | xxx (xx.x) | xxx (xx.x) | xxx (xx.x) |
| Posterior circulation | xxx | xxx (xx.x) | xxx (xx.x) | xxx (xx.x) |
| Weight (kg) | xxx | xx.x (xx.x) | xx.x (xx.x) | xx.x (xx.x) |

DSRS (dysphagia rating scale); EAT-10: eating assessment tool-10; FOIS: functional oral intake score; IDDSI: international dysphagia diet standardisation initiative; NIHSS (NIH stroke severity scale); OCSP: Oxfordshire community stroke project; TACS: total anterior circulation syndrome.
Data are number (%), median [interquartile range] or mean (standard deviation).

**Table S3-2.**
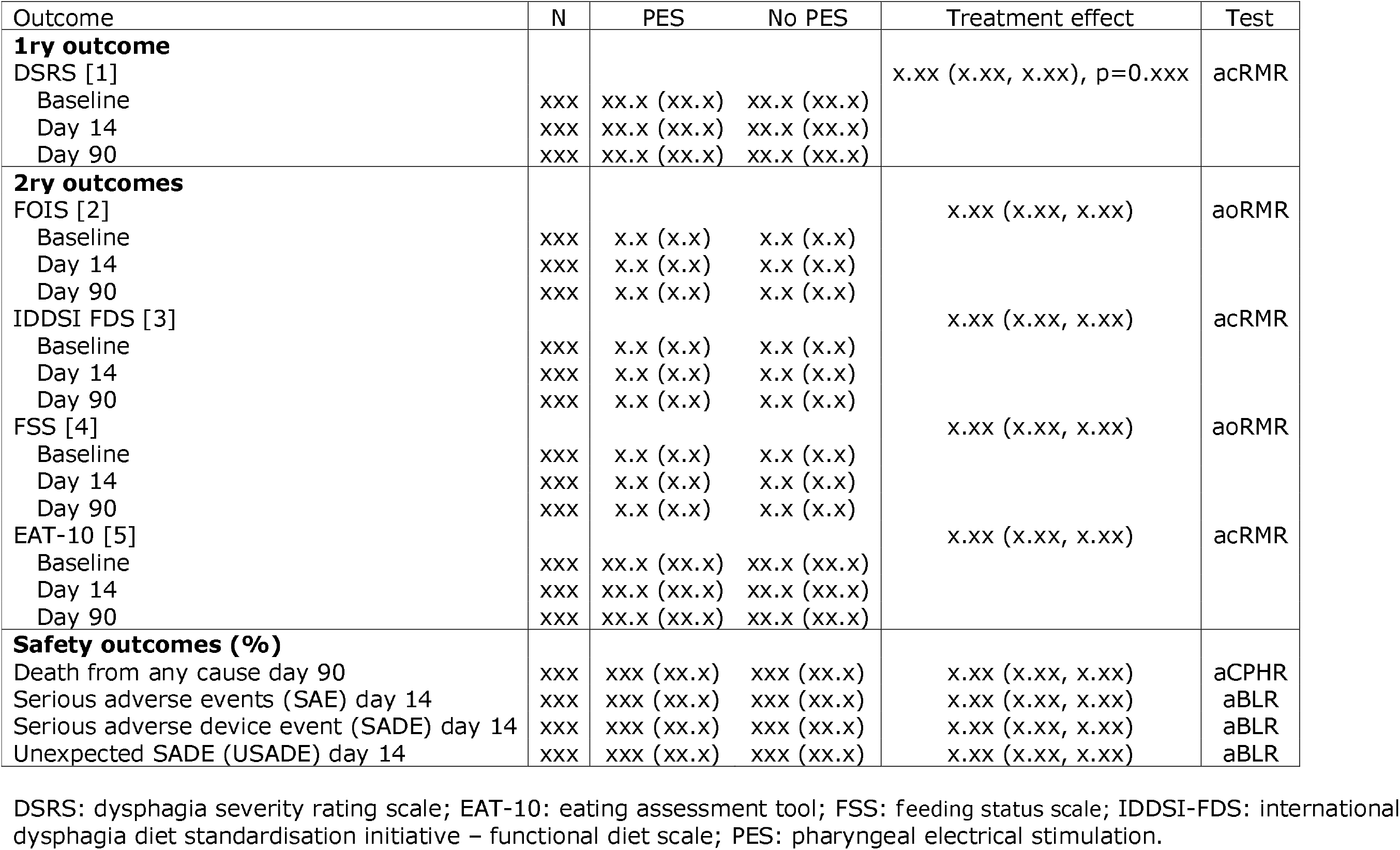
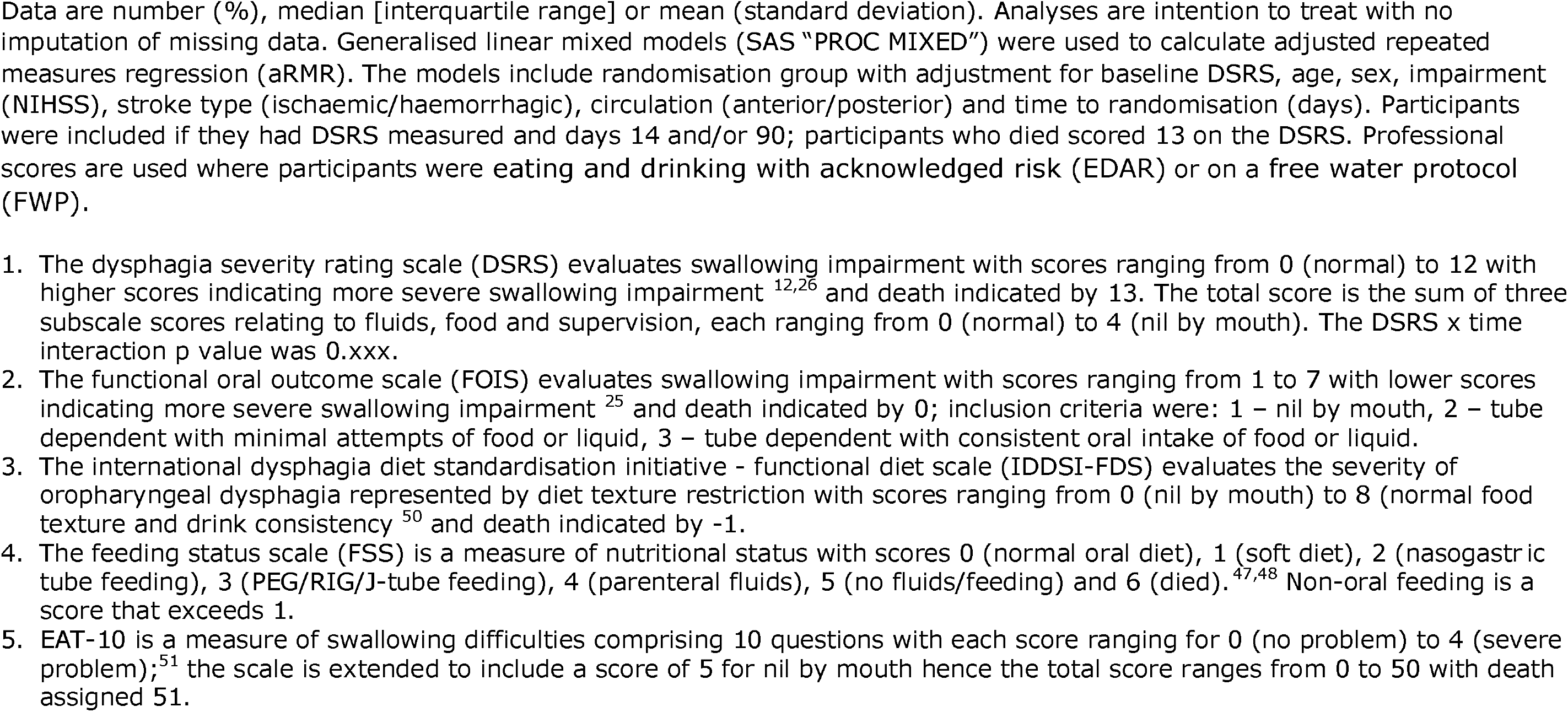
Primary and secondary swallowing and feeding outcomes at days 14 and 90 with no imputation. Analyses by adjusted continuous repeated measures regression (acRMR), adjusted ordinal repeated measures regression (aoRMR), adjusted Cox proportional hazards regression (aCPHR). Or adjusted binary logistic regression (aBLR)

| Outcome | N | PES | No PES | Treatment effect | Test |
| --- | --- | --- | --- | --- | --- |
| **1ry outcome** |  |  |  |  |  |
| DSRS [1] |  |  |  | x.xx (x.xx, x.xx), p=0.xxx | acRMR |
| Baseline | xxx | xx.x (xx.x) | xx.x (xx.x) |  |  |
| Day 14 | xxx | xx.x (xx.x) | xx.x (xx.x) |  |  |
| Day 90 | xxx | xx.x (xx.x) | xx.x (xx.x) |  |  |
| **2ry outcomes** |  |  |  |  |  |
| FOIS [2] |  |  |  | x.xx (x.xx, x.xx) | aoRMR |
| Baseline | xxx | x.x (x.x) | x.x (x.x) |  |  |
| Day 14 | xxx | x.x (x.x) | x.x (x.x) |  |  |
| Day 90 | xxx | x.x (x.x) | x.x (x.x) |  |  |
| IDDSI FDS [3] |  |  |  | x.xx (x.xx, x.xx) | acRMR |
| Baseline | xxx | x.x (x.x) | x.x (x.x) |  |  |
| Day 14 | xxx | x.x (x.x) | x.x (x.x) |  |  |
| Day 90 | xxx | x.x (x.x) | x.x (x.x) |  |  |
| FSS [4] |  |  |  | x.xx (x.xx, x.xx) | aoRMR |
| Baseline | xxx | x.x (x.x) | x.x (x.x) |  |  |
| Day 14 | xxx | x.x (x.x) | x.x (x.x) |  |  |
| Day 90 | xxx | x.x (x.x) | x.x (x.x) |  |  |
| EAT-10 [5] |  |  |  | x.xx (x.xx, x.xx) | acRMR |
| Baseline | xxx | xx.x (xx.x) | xx.x (xx.x) |  |  |
| Day 14 | xxx | xx.x (xx.x) | xx.x (xx.x) |  |  |
| Day 90 | xxx | xx.x (xx.x) | xx.x (xx.x) |  |  |
| **Safety outcomes (%)** |  |  |  |  |  |
| Death from any cause day 90 | xxx | xxx (xx.x) | xxx (xx.x) | x.xx (x.xx, x.xx) | aCPHR |
| Serious adverse events (SAE) day 14 | xxx | xxx (xx.x) | xxx (xx.x) | x.xx (x.xx, x.xx) | aBLR |
| Serious adverse device event (SADE) day 14 | xxx | xxx (xx.x) | xxx (xx.x) | x.xx (x.xx, x.xx) | aBLR |
| Unexpected SADE (USADE) day 14 | xxx | xxx (xx.x) | xxx (xx.x) | x.xx (x.xx, x.xx) | aBLR |

DSRS: dysphagia severity rating scale; EAT-10: eating assessment tool; FSS: feeding status scale; IDDSI-FDS: international dysphagia diet standardisation initiative – functional diet scale; PES: pharyngeal electrical stimulation.
Data are number (%), median [interquartile range] or mean (standard deviation). Analyses are intention to treat with no imputation of missing data. Generalised linear mixed models (SAS "PROC MIXED") were used to calculate adjusted repeated measures regression (aRMR). The models include randomisation group with adjustment for baseline DSRS, age, sex, impairment (NIHSS), stroke type (ischaemic/haemorrhagic), circulation (anterior/posterior) and time to randomisation (days). Participants were included if they had DSRS measured and days 14 and/or 90; participants who died scored 13 on the DSRS. Professional scores are used where participants were eating and drinking with acknowledged risk (EDAR) or on a free water protocol (FWP).

**Figure 1. Swallowing impairment at 14 and 90 days in the two groups according to scores on the dysphagia severity rating scale (primary outcome).**

| Shift | Score | 0 | 1 | 2 | 3 | 4 | 5 | 6 | 7 | 8 | 9 | 10 | 11 | 12 | 13 |
| --- | --- | --- | --- | --- | --- | --- | --- | --- | --- | --- | --- | --- | --- | --- | --- |
|  | Day |  |  |  |  |  |  |  |  |  |  |  |  |  |  |
| PES | 00 | xx | xx | xx | xx | xx | xx | xx | xx | xx | xx | xx | xx | xx | 0 |
|  | 14 | xx | xx | xx | xx | xx | xx | xx | xx | xx | xx | xx | xx | xx | xx |
|  | 90 | xx | xx | xx | xx | xx | xx | xx | xx | xx | xx | xx | xx | xx | xx |
| No PES | 90 | xx | xx | xx | xx | xx | xx | xx | xx | xx | xx | xx | xx | xx | xx |
|  | 14 | xx | xx | xx | xx | xx | xx | xx | xx | xx | xx | xx | xx | xx | xx |
|  | 00 | xx | xx | xx | xx | xx | xx | xx | xx | xx | xx | xx | xx | xx | 0 |

Scores on the dysphagia severity rating scale range from 0 to 13 with 0 indicating no swallowing impairment/normal diet, higher scores indicating more severe swallowing impairment, 12 indicating nil by mouth and 13 indicating death.^12,26^ The total score is the sum of three subscales relating to fluids, food and supervision, each ranging from 0 (normal) to 4 (nil by mouth).

**Figure 2. Cumulative incidence of death out to 90 days with analysis by Cox proportional hazards regression.**

Panel shows inset with an enlarged y axis.

**Figure 3. Subgroup analysis of the primary outcome.**

Shown are the difference in means for swallowing impairment in pre-specified subgroups including minimisation factors. The widths of the confidence intervals have not been adjusted for multiplicity and should not be used in place of hypothesis testing. DSRS scores range from 0 to 12, with 13 indicating death and higher scores indicating worse swallowing impairment. FOIS scores range from 1 to 7, with 0 indicating death and lower scores indicating worse swallowing impairment. NIHSS scores range from 0 to 42, with 43 indicting death and higher scores indicating greater neurological impairment. Age, FOIS, DSRS, NIHSS and time are dichotomised as shown.

- Age (years): <75, >=75
- Sex: female, male
- Reperfusion: yes, no
- Dysphagia severity rating scale (/12): <11, 11, 12
- Functional oral intake scale (/7): 1, 2/3
- Stroke severity, NIHSS (/42): <13, > = 13
- Stroke type: ischaemic, primary intracerebral haemorrhage
- Circulation: anterior, posterior
- Onset to randomisation (days): <11, 11-20, >20
- Lesion side: right, bilateral, left
- Stroke size: very small/small, medium, large/very large
- White matter disease: none, mild, moderate, severe
- Prior instrumental testing (FEES/VFS): yes, no
- Responder, DSRS change day 90-0: >-3.5, <=-3.5
- Responder, DSRS day 90: <4, >=4
- Experience with PES prior to trial: yes, no
- Site recruitment: >10, < = 10

FOREST PLOT WITH INTERACTION TERMS

### SUPPLEMENT FOR MAIN PAPER

**Supplement to main paper**

**Table S1.** Reasons for excluding from participating in the trial.

| **Reason †** | **N (%)** |
| --- | --- |
| FOIS >3 | XXXX (xx.x) |
| Premorbid dysphagia | XXXX (xx.x) |
| Palliative/end-of-life care | XXXX (xx.x) |
| Repatriated | XXXX (xx.x) |
| Frequent removal of nasogastric tubes | XXXX (xx.x) |
| Declined consent | XXXX (xx.x) |
| Pacemaker/cochlear implant/ICD | XXXX (xx.x) |
| Out of time window | XXXX (xx.x) |
| No clinical indication for NGT or PEG | XXXX (xx.x) |
| Non-stroke dysphagia | XXXX (xx.x) |
| Pre-stroke mRS of 4 or 5 | XXXX (xx.x) |
| Ongoing/planned ventilation, intubation or tracheostomy | XXXX (xx.x) |
| Planned electrical/magnetic stimulation for dysphagia | XXXX (xx.x) |
| Clinical indication for >35% oxygen | XXXX (xx.x) |
| Patient unlikely to tolerate PES catheter/treatment | XXXX (xx.x) |
| Active malignant MCA syndrome | XXXX (xx.x) |
| Recruited into another clinical trial | XXXX (xx.x) |
| No staff available to recruit or complete procedures | XXXX (xx.x) |
| Patient or next of kin declined | XXXX (xx.x) |
| Patient unfit for research due to other comorbidities | XXXX (xx.x) |
| Language barrier | XXXX (xx.x) |
| Eating & drinking with acknowledged risk (EDAR) | XXXX (xx.x) |
| Not appropriate for swallow assessment(s) | XXXX (xx.x) |
| Pregnant | XXXX (xx.x) |
| Presence of pharyngeal pouch | XXXX (xx.x) |
| NIHSS score 1a = >2 | XXXX (xx.x) |
| Patient from overseas so unable to follow-up | XXXX (xx.x) |
| Dysphagia considered to be short-term only | XXXX (xx.x) |
| Total | XXXX |

MCA: middle cerebral artery
† Items are not mutually exclusive, i.e. patients may have been excluded for more than one reason.
TO BE ORDERED BY FREQUENCY

**Table S2.** Clinical examination of swallowing at baseline and prognostication for recovery.

| **Characteristic** | **N** | **All** | **PES Group**<br>**(N=xxx)** | **No PES Group**<br>**(N=xxx)** |
| --- | --- | --- | --- | --- |
| Dysphonia (%) | xxx | xxx (xx.x) | xxx (xx.x) | xxx (xx.x) |
| Dysarthria (%) | xxx | xxx (xx.x) | xxx (xx.x) | xxx (xx.x) |
| Abnormal gag reflex (%) | xxx | xxx (xx.x) | xxx (xx.x) | xxx (xx.x) |
| Abnormal volitional cough (%) | xxx | xxx (xx.x) | xxx (xx.x) | xxx (xx.x) |
| Cough after swallow (%) | xxx | xxx (xx.x) | xxx (xx.x) | xxx (xx.x) |
| Voice change after swallow (%) | xxx | xxx (xx.x) | xxx (xx.x) | xxx (xx.x) |
| Frontal operculum involved † | xxx | xxx (xx.x) | xxx (xx.x) | xxx (xx.x) |
| Calculated PRESS score ‡ | xxx | xx.x (xx.x) | xxx (xx.x) | xxx (xx.x) |

† Brain scans before or at baseline were not collected and adjudicated; admission neuroradiology reports (see Table S3 below) were entered by recruiting sites; some of these reported the presence of a stroke lesion (infarct or haemorrhage) at the frontal operculum although this item may have been under-reported and so the total PRESS score will be underestimated by one point for some participants.
‡ The predictive swallowing score (PRESS<sup>61</sup>) is a prognostic instrument that predicts swallowing recovery by 30 days after stroke and ranges from 0 (no features present) to 10 (10 features present) based on age $\geq 70$ years; NIHSS score; stroke location includes lesion at the frontal operculum; presence of dysphonia, dysarthria, abnormal gag reflex, abnormal volitional cough, cough after swallow and voice change after swallow; and FOIS score. Scoring is given in reference.<sup>61</sup> Brain scans before or at baseline were not collected and adjudicated. Instead, relevant neuroradiology reports were entered by recruiting sites but few made any specific comment about the presence of a stroke lesion at the frontal operculum. Hence, this item may have been under-reported and so the total PRESS score is likely an underestimate.

**Table S3.** Neuroimaging features as recorded by site’s (neuro)-radiologist imaging report between admission to hospital and randomisation.

| **Feature** | **N (%)** | **All**<br>**(N=xxx)** | **PES**<br>**(n1=xxx)** | **No PES**<br>**(n2=xxx)** |
| --- | --- | --- | --- | --- |
| CT scan | xxx (xx.x) | xxx (xx.x) | xxx (xx.x) | xxx (xx.x) |
| MR scan | xxx (xx.x) | xxx (xx.x) | xxx (xx.x) | xxx (xx.x) |
| ***Reported by site (%)*** | xxx (xx.x) |  |  |  |
| Normal scan |  | xxx (xx.x) | xxx (xx.x) | xxx (xx.x) |
| Infarct |  | xxx (xx.x) | xxx (xx.x) | xxx (xx.x) |
| Infarct with HTI |  | xxx (xx.x) | xxx (xx.x) | xxx (xx.x) |
| ICH |  | xxx (xx.x) | xxx (xx.x) | xxx (xx.x) |
| SAH |  | xxx (xx.x) | xxx (xx.x) | xxx (xx.x) |
| Other lesion |  | xxx (xx.x) | xxx (xx.x) | xxx (xx.x) |
| ***Ischaemic stroke*** | xxx (xx.x) | xxx (xx.x) | xxx (xx.x) | xxx (xx.x) |
| Side | xxx (xx.x) |  |  |  |
| Right |  | xxx (xx.x) | xxx (xx.x) | xxx (xx.x) |
| Left |  | xxx (xx.x) | xxx (xx.x) | xxx (xx.x) |
| Bilateral/midline |  | xxx (xx.x) | xxx (xx.x) | xxx (xx.x) |
| Size | xxx (xx.x) |  |  |  |
| Very large |  | xxx (xx.x) | xxx (xx.x) | xxx (xx.x) |
| Large |  | xxx (xx.x) | xxx (xx.x) | xxx (xx.x) |
| Moderate |  | xxx (xx.x) | xxx (xx.x) | xxx (xx.x) |
| Small |  | xxx (xx.x) | xxx (xx.x) | xxx (xx.x) |
| Very small |  | xxx (xx.x) | xxx (xx.x) | xxx (xx.x) |
| Hyperdense artery | xxx (xx.x) | xxx (xx.x) | xxx (xx.x) | xxx (xx.x) |
| Site | xxx (xx.x) |  |  |  |
| Frontal |  | xxx (xx.x) | xxx (xx.x) | xxx (xx.x) |
| Parietal |  | xxx (xx.x) | xxx (xx.x) | xxx (xx.x) |
| Temporal |  | xxx (xx.x) | xxx (xx.x) | xxx (xx.x) |
| Subcortical |  | xxx (xx.x) | xxx (xx.x) | xxx (xx.x) |
| Basal ganglia |  | xxx (xx.x) | xxx (xx.x) | xxx (xx.x) |
| Internal capsule |  | xxx (xx.x) | xxx (xx.x) | xxx (xx.x) |
| Pontine |  | xxx (xx.x) | xxx (xx.x) | xxx (xx.x) |
| Brain stem |  | xxx (xx.x) | xxx (xx.x) | xxx (xx.x) |
| Cerebellum |  | xxx (xx.x) | xxx (xx.x) | xxx (xx.x) |
| Local sites | xxx (xx.x) |  |  |  |
| Insula |  | xxx (xx.x) | xxx (xx.x) | xxx (xx.x) |
| Frontal operculum |  | xxx (xx.x) | xxx (xx.x) | xxx (xx.x) |
| Small vessel disease | xxx (xx.x) | xxx (xx.x) | xxx (xx.x) | xxx (xx.x) |
| Lacunar infarction |  | xxx (xx.x) | xxx (xx.x) | xxx (xx.x) |
| Lacune(s) |  | xxx (xx.x) | xxx (xx.x) | xxx (xx.x) |
| White matter change |  | xxx (xx.x) | xxx (xx.x) | xxx (xx.x) |
| SVD score [/3] | xxx (xx.x) | x.x (x.x) | x.x (x.x) | x.x (x.x) |
| Brain frailty score [/3] | xxx (xx.x) | x.x (x.x) | x.x (x.x) | x.x (x.x) |
| ***ICH*** | xxx (xx.x) | xxx (xx.x) | xxx (xx.x) | xxx (xx.x) |
| Side | xxx (xx.x) |  |  |  |
| Right |  | xxx (xx.x) | xxx (xx.x) | xxx (xx.x) |
| Left |  | xxx (xx.x) | xxx (xx.x) | xxx (xx.x) |
| Bilateral/midline |  | xxx (xx.x) | xxx (xx.x) | xxx (xx.x) |
| Size | xxx (xx.x) | xxx (xx.x) | xxx (xx.x) | xxx (xx.x) |
| Very large |  | xxx (xx.x) | xxx (xx.x) | xxx (xx.x) |
| Large |  | xxx (xx.x) | xxx (xx.x) | xxx (xx.x) |
| Moderate |  | xxx (xx.x) | xxx (xx.x) | xxx (xx.x) |
| Small |  | xxx (xx.x) | xxx (xx.x) | xxx (xx.x) |
| Very small |  | xxx (xx.x) | xxx (xx.x) | xxx (xx.x) |
| Site | xxx (xx.x) |  |  |  |
| Basal ganglia |  | xxx (xx.x) | xxx (xx.x) | xxx (xx.x) |
| Brainstem |  | xxx (xx.x) | xxx (xx.x) | xxx (xx.x) |
| Cerebellum |  | xxx (xx.x) | xxx (xx.x) | xxx (xx.x) |
| Centrum semiovale |  | xxx (xx.x) | xxx (xx.x) | xxx (xx.x) |
| Corona radiata |  | xxx (xx.x) | xxx (xx.x) | xxx (xx.x) |
| Frontal |  | xxx (xx.x) | xxx (xx.x) | xxx (xx.x) |
| Midbrain |  | xxx (xx.x) | xxx (xx.x) | xxx (xx.x) |
| Occipital |  | xxx (xx.x) | xxx (xx.x) | xxx (xx.x) |
| Parietal |  | xxx (xx.x) | xxx (xx.x) | xxx (xx.x) |
| Temporal |  | xxx (xx.x) | xxx (xx.x) | xxx (xx.x) |
| Other sites |  |  |  |  |
| Frontal operculum |  | xxx (xx.x) | xxx (xx.x) | xxx (xx.x) |
| **Background** | xxx (xx.x) |  |  |  |
| Atrophy |  | xxx (xx.x) | xxx (xx.x) | xxx (xx.x) |
| Old infarct |  | xxx (xx.x) | xxx (xx.x) | xxx (xx.x) |
| **Other diagnoses** |  |  |  |  |
| Intraventricular haemorrhage | xxx (xx.x) | xxx (xx.x) | xxx (xx.x) | xxx (xx.x) |
| Subarachnoid haemorrhage | xxx (xx.x) | xxx (xx.x) | xxx (xx.x) | xxx (xx.x) |
| Subdural | xxx (xx.x) | xxx (xx.x) | xxx (xx.x) | xxx (xx.x) |
| Arteriovenous malformation | xxx (xx.x) | xxx (xx.x) | xxx (xx.x) | xxx (xx.x) |
| Aneurysm | xxx (xx.x) | xxx (xx.x) | xxx (xx.x) | xxx (xx.x) |

† Participants may have multiple features. Data are n (%)
Radiology reports were not available for all participants so numbers (%) may differ from those reported by investigators.
Imaging report interpreted by algorithmic natural language processing: accuracy depends on the structure, detail, completeness and quality of the radiologist report and robustness of the NLP program to non-standard English and missing data. Hence the numbers should be considered approximate and incomplete.

**Table S4.** Hospital care before and after randomisation.

| **Procedure** | **N** | **All** | **PES** | **No PES** | **aDIM/acOR/aOR**<br>**(95% CI)<sup>†</sup>** | **Analysis** |
| --- | --- | --- | --- | --- | --- | --- |
| ***Pre-randomisation (%)*** |  | xxx | xxx | xxx | / | / |
| Intravenous thrombolysis | xxx | xxx (xx.x) | xxx (xx.x) | xxx (xx.x) |  |  |
| Mechanical thrombectomy | xxx | xxx (xx.x) | xxx (xx.x) | xxx (xx.x) |  |  |
| Hemicraniectomy | xxx | xxx (xx.x) | xxx (xx.x) | xxx (xx.x) |  |  |
| Neurosurgery, other | xxx | xxx (xx.x) | xxx (xx.x) | xxx (xx.x) |  |  |
| Vascular surgery | xxx | xxx (xx.x) | xxx (xx.x) | xxx (xx.x) |  |  |
| Intensive care unit admission | xxx | xxx (xx.x) | xxx (xx.x) | xxx (xx.x) |  |  |
| Ventilation | xxx | xxx (xx.x) | xxx (xx.x) | xxx (xx.x) |  |  |
| Days of ventilation | xxx | xxx (xx.x) | xxx (xx.x) | xxx (xx.x) |  |  |
| Tracheostomy | xxx | xxx (xx.x) | xxx (xx.x) | xxx (xx.x) |  |  |
| Days with a tracheostomy | xxx | xxx (xx.x) | xxx (xx.x) | xxx (xx.x) |  |  |
| ***Post-randomisation (%)*** |  | xxx | xxx | xxx |  |  |
| Neurosurgery | xxx | xxx (xx.x) | xxx (xx.x) | xxx (xx.x) | x.xx (x.xx, x.xx) | aBLR |
| Vascular surgery | xxx | xxx (xx.x) | xxx (xx.x) | xxx (xx.x) | x.xx (x.xx, x.xx) | aBLR |
| Intensive care unit admission | xxx | xxx (xx.x) | xxx (xx.x) | xxx (xx.x) | x.xx (x.xx, x.xx) | aBLR |
| Ventilation | xxx | xxx (xx.x) | xxx (xx.x) | xxx (xx.x) | x.xx (x.xx, x.xx) | aBLR |
| Tracheostomy | xxx | xxx (xx.x) | xxx (xx.x) | xxx (xx.x) | x.xx (x.xx, x.xx) | aBLR |
| ***At discharge/death in hospital (%)*** |  | xxx | xxx | xxx |  |  |
| Antithrombotic | xxx | xxx (xx.x) | xxx (xx.x) | xxx (xx.x) | x.xx (x.xx, x.xx) | aBLR |
| Antiplatelet | xxx | xxx (xx.x) | xxx (xx.x) | xxx (xx.x) |  |  |
| Anticoagulant, oral | xxx | xxx (xx.x) | xxx (xx.x) | xxx (xx.x) |  |  |
| Statin | xxx | xxx (xx.x) | xxx (xx.x) | xxx (xx.x) | x.xx (x.xx, x.xx) | aBLR |
| Antihypertensive | xxx | xxx (xx.x) | xxx (xx.x) | xxx (xx.x) | x.xx (x.xx, x.xx) | aBLR |
| Gastroprotection | xxx | xxx (xx.x) | xxx (xx.x) | xxx (xx.x) | x.xx (x.xx, x.xx) | aBLR |

aDIM: adjusted difference in means; acOR: adjusted common odds ratio; aOR: adjusted odds ratio; CI: confidence intervals; PES: pharyngeal electrical stimulation

**Table S5.** Adherence to trial allocation.

| **Assessment** | **PES** |
| --- | --- |
| Participants | xxx |
| ***PES doses received (%)*** |  |
| First dose received | xxx (xx.x) |
| Doses received, at least |  |
| 1 | xxx (xx.x) |
| 2 | xxx (xx.x) |
| 3 | xxx (xx.x) |
| 4 | xxx (xx.x) |
| 5 | xxx (xx.x) |
| 6, all | xxx (xx.x) |
| Number of doses, median [IQR] | x.x [x.x, x.x] |
| ***Reasons for non-adherence to first three treatments (%)*** |  |
| Withdrawal from treatment | xx (xx) |
| Withdrawal from trial | xx (xx) |
| Clinical decision by hospital staff | xx (xx) |
| Participant discharged | xx (xx) |
| Lost to follow-up | xx (xx) |
| PES failure | xx (xx) |
| Died | xx (xx) |
| Other | xx (xx) |

**Table S6.** Treatment with pharyngeal electrical stimulation: threshold, tolerance, calculated stimulation, actual stimulation and delta (tolerability-threshold), all in milliamps (mA), and treatment duration in participants randomised to PES. Data are number (%), median [interquartile range] or mean (standard deviation).

| Treatment | N | Threshold | Tolerance | Calculated stimulation | Stimulation | Delta | Duration (secs) |
| --- | --- | --- | --- | --- | --- | --- | --- |
| 1 | xxx | xx.x (xx.x) | xx.x (xx.x) | xx.x (xx.x) | xx.x (xx.x) | xx.x (xx.x) | xxx.x [xxx.x, xxx.x] |
| 2 | xxx | xx.x (xx.x) | xx.x (xx.x) | xx.x (xx.x) | xx.x (xx.x) | xx.x (xx.x) | xxx.x [xxx.x, xxx.x] |
| 3 | xxx | xx.x (xx.x) | xx.x (xx.x) | xx.x (xx.x) | xx.x (xx.x) | xx.x (xx.x) | xxx.x [xxx.x, xxx.x] |
| 4 | xxx | xx.x (xx.x) | xx.x (xx.x) | xx.x (xx.x) | xx.x (xx.x) | xx.x (xx.x) | xxx.x [xxx.x, xxx.x] |
| 5 | xxx | xx.x (xx.x) | xx.x (xx.x) | xx.x (xx.x) | xx.x (xx.x) | xx.x (xx.x) | xxx.x [xxx.x, xxx.x] |
| 6 | xxx | xx.x (xx.x) | xx.x (xx.x) | xx.x (xx.x) | xx.x (xx.x) | xx.x (xx.x) | xxx.x [xxx.x, xxx.x] |
| Average | xxx | xx.x (xx.x) | xx.x (xx.x) | xx.x (xx.x) | xx.x (xx.x) | xx.x (xx.x) | xxx.x [xxx.x, xxx.x] |

**Table S7.** Treatment with pharyngeal electrical stimulation: participants receiving adequate stimulation.

| | N | Stimulation $\geq 20$mA | Delta $> 20$mA<sup>†</sup> | Time $\geq 9$m50s | Catheters used (SD) |
| --- | --- | --- | --- | --- | --- |
| Participants (%) | xxx | xxx (xx.x) | xxx (xx.x) | xxx (xx.x) | x.x (x.x) |
| Treatments (%) | xxxx | xxxx (xx.x) | xxxx (xx.x) | xxxx (xx.x) | / |

<sup>†</sup> Delta = tolerance - threshold

**Table S8.**
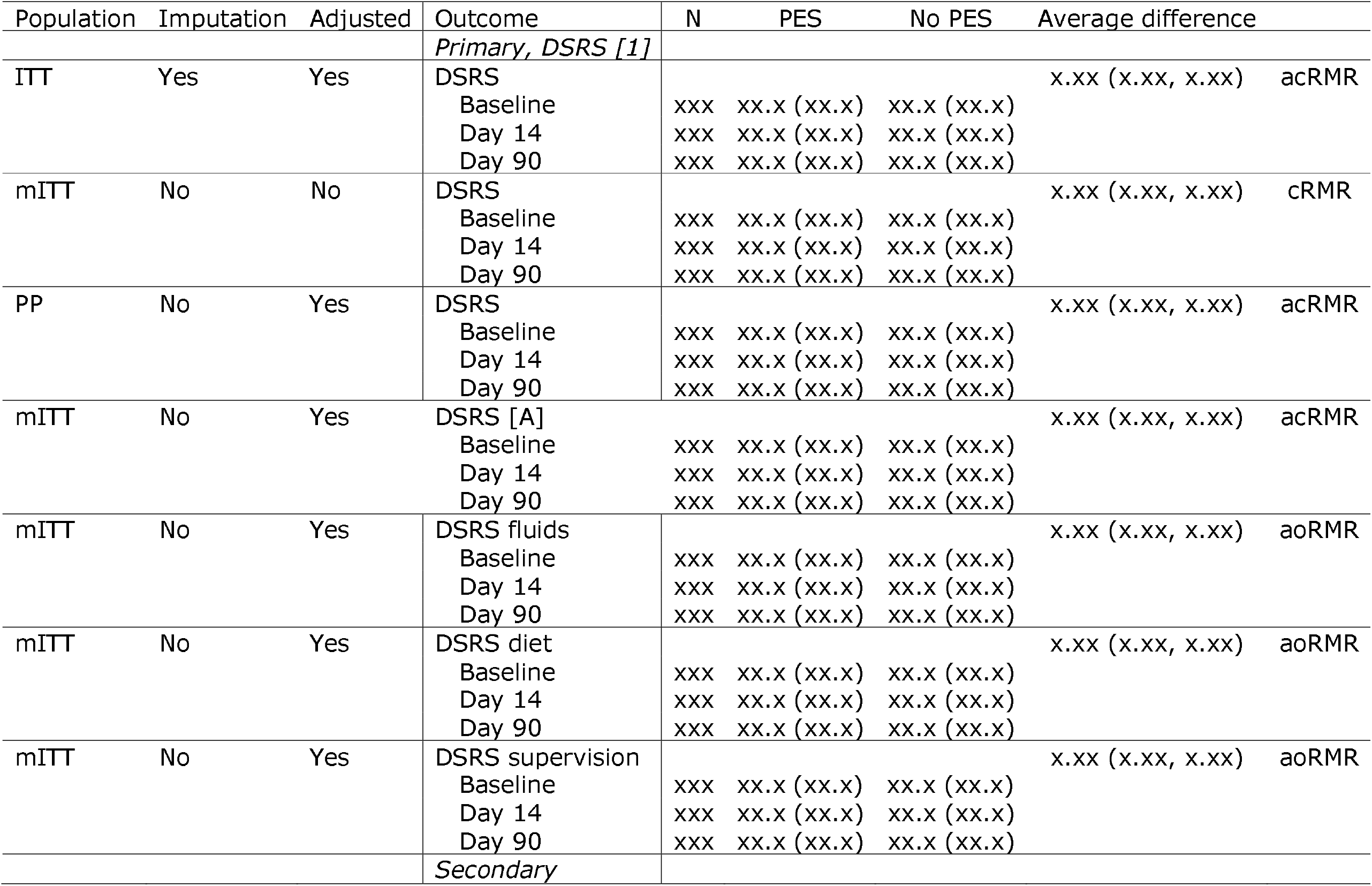
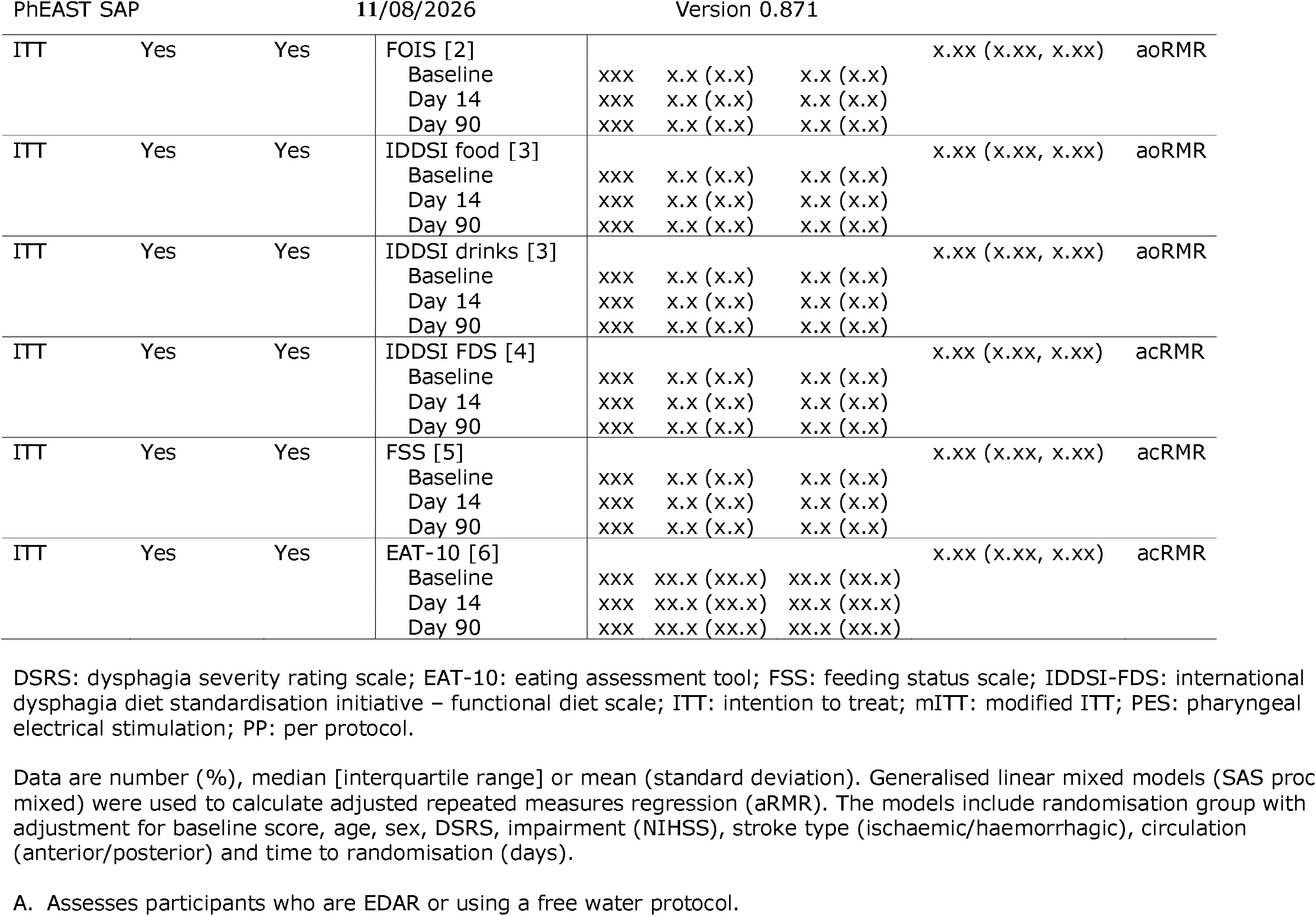
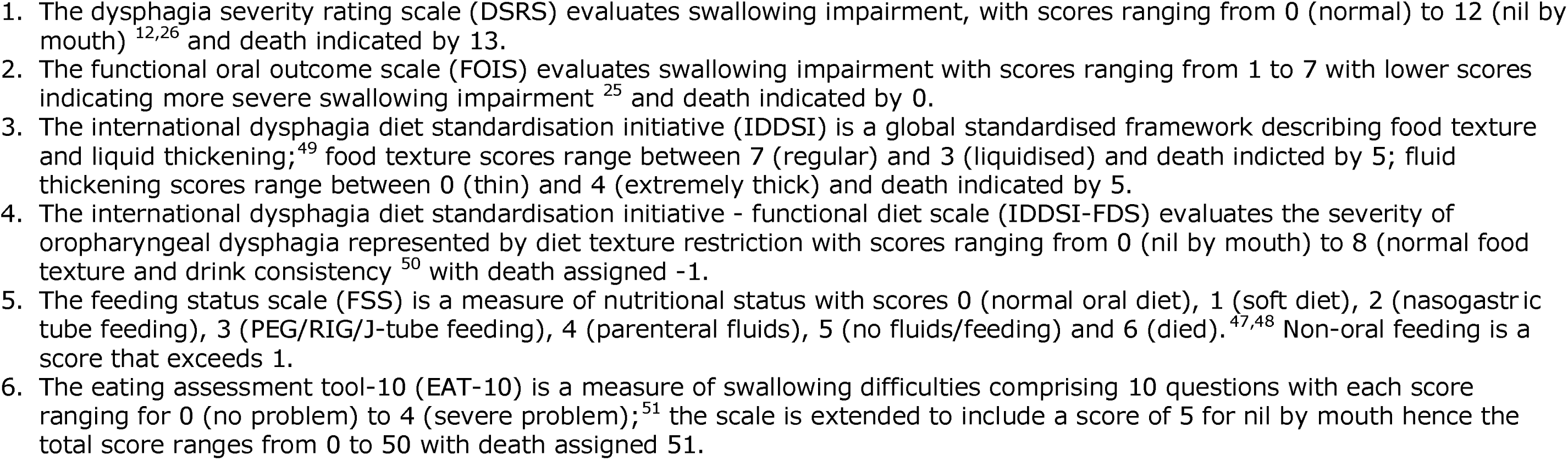
Swallowing and feeding outcomes at days 14 and 90. Sensitivity analyses by continuous repeated measures regression (cRMR) or ordinal repeated measures regression (oRMR) with adjustment.

| Population | Imputation | Adjusted | Outcome | N | PES | No PES | Average difference |  |
| --- | --- | --- | --- | --- | --- | --- | --- | --- |
|  |  |  | *Primary, DSRS [1]* |  |  |  |  |  |
| ITT | Yes | Yes | DSRS |  |  |  | x.xx (x.xx, x.xx) | acRMR |
|  |  |  | Baseline | xxx | xx.x (xx.x) | xx.x (xx.x) |  |  |
|  |  |  | Day 14 | xxx | xx.x (xx.x) | xx.x (xx.x) |  |  |
|  |  |  | Day 90 | xxx | xx.x (xx.x) | xx.x (xx.x) |  |  |
| mITT | No | No | DSRS |  |  |  | x.xx (x.xx, x.xx) | cRMR |
|  |  |  | Baseline | xxx | xx.x (xx.x) | xx.x (xx.x) |  |  |
|  |  |  | Day 14 | xxx | xx.x (xx.x) | xx.x (xx.x) |  |  |
|  |  |  | Day 90 | xxx | xx.x (xx.x) | xx.x (xx.x) |  |  |
| PP | No | Yes | DSRS |  |  |  | x.xx (x.xx, x.xx) | acRMR |
|  |  |  | Baseline | xxx | xx.x (xx.x) | xx.x (xx.x) |  |  |
|  |  |  | Day 14 | xxx | xx.x (xx.x) | xx.x (xx.x) |  |  |
|  |  |  | Day 90 | xxx | xx.x (xx.x) | xx.x (xx.x) |  |  |
| mITT | No | Yes | DSRS [A] |  |  |  | x.xx (x.xx, x.xx) | acRMR |
|  |  |  | Baseline | xxx | xx.x (xx.x) | xx.x (xx.x) |  |  |
|  |  |  | Day 14 | xxx | xx.x (xx.x) | xx.x (xx.x) |  |  |
|  |  |  | Day 90 | xxx | xx.x (xx.x) | xx.x (xx.x) |  |  |
| mITT | No | Yes | DSRS fluids |  |  |  | x.xx (x.xx, x.xx) | aoRMR |
|  |  |  | Baseline | xxx | xx.x (xx.x) | xx.x (xx.x) |  |  |
|  |  |  | Day 14 | xxx | xx.x (xx.x) | xx.x (xx.x) |  |  |
|  |  |  | Day 90 | xxx | xx.x (xx.x) | xx.x (xx.x) |  |  |
| mITT | No | Yes | DSRS diet |  |  |  | x.xx (x.xx, x.xx) | aoRMR |
|  |  |  | Baseline | xxx | xx.x (xx.x) | xx.x (xx.x) |  |  |
|  |  |  | Day 14 | xxx | xx.x (xx.x) | xx.x (xx.x) |  |  |
|  |  |  | Day 90 | xxx | xx.x (xx.x) | xx.x (xx.x) |  |  |
| mITT | No | Yes | DSRS supervision |  |  |  | x.xx (x.xx, x.xx) | aoRMR |
|  |  |  | Baseline | xxx | xx.x (xx.x) | xx.x (xx.x) |  |  |
|  |  |  | Day 14 | xxx | xx.x (xx.x) | xx.x (xx.x) |  |  |
|  |  |  | Day 90 | xxx | xx.x (xx.x) | xx.x (xx.x) |  |  |
|  |  |  | *Secondary* |  |  |  |  |  |
| ITT | Yes | Yes | FOIS [2]<br>Baseline<br>Day 14<br>Day 90 | xxx<br>xxx<br>xxx | x.x (x.x)<br>x.x (x.x)<br>x.x (x.x) | x.x (x.x)<br>x.x (x.x)<br>x.x (x.x) | x.xx (x.xx, x.xx) | aoRMR |
| ITT | Yes | Yes | IDDSI food [3]<br>Baseline<br>Day 14<br>Day 90 | xxx<br>xxx<br>xxx | x.x (x.x)<br>x.x (x.x)<br>x.x (x.x) | x.x (x.x)<br>x.x (x.x)<br>x.x (x.x) | x.xx (x.xx, x.xx) | aoRMR |
| ITT | Yes | Yes | IDDSI drinks [3]<br>Baseline<br>Day 14<br>Day 90 | xxx<br>xxx<br>xxx | x.x (x.x)<br>x.x (x.x)<br>x.x (x.x) | x.x (x.x)<br>x.x (x.x)<br>x.x (x.x) | x.xx (x.xx, x.xx) | aoRMR |
| ITT | Yes | Yes | IDDSI FDS [4]<br>Baseline<br>Day 14<br>Day 90 | xxx<br>xxx<br>xxx | x.x (x.x)<br>x.x (x.x)<br>x.x (x.x) | x.x (x.x)<br>x.x (x.x)<br>x.x (x.x) | x.xx (x.xx, x.xx) | acRMR |
| ITT | Yes | Yes | FSS [5]<br>Baseline<br>Day 14<br>Day 90 | xxx<br>xxx<br>xxx | x.x (x.x)<br>x.x (x.x)<br>x.x (x.x) | x.x (x.x)<br>x.x (x.x)<br>x.x (x.x) | x.xx (x.xx, x.xx) | acRMR |
| ITT | Yes | Yes | EAT-10 [6]<br>Baseline<br>Day 14<br>Day 90 | xxx<br>xxx<br>xxx | xx.x (xx.x)<br>xx.x (xx.x)<br>xx.x (xx.x) | xx.x (xx.x)<br>xx.x (xx.x)<br>xx.x (xx.x) | x.xx (x.xx, x.xx) | acRMR |

DSRS: dysphagia severity rating scale; EAT-10: eating assessment tool; FSS: feeding status scale; IDDSI-FDS: international dysphagia diet standardisation initiative – functional diet scale; ITT: intention to treat; mITT: modified ITT; PES: pharyngeal electrical stimulation; PP: per protocol.
Data are number (%), median [interquartile range] or mean (standard deviation). Generalised linear mixed models (SAS proc mixed) were used to calculate adjusted repeated measures regression (aRMR). The models include randomisation group with adjustment for baseline score, age, sex, DSRS, impairment (NIHSS), stroke type (ischaemic/haemorrhagic), circulation (anterior/posterior) and time to randomisation (days).
A. Assesses participants who are EDAR or using a free water protocol.

**Table S9.**
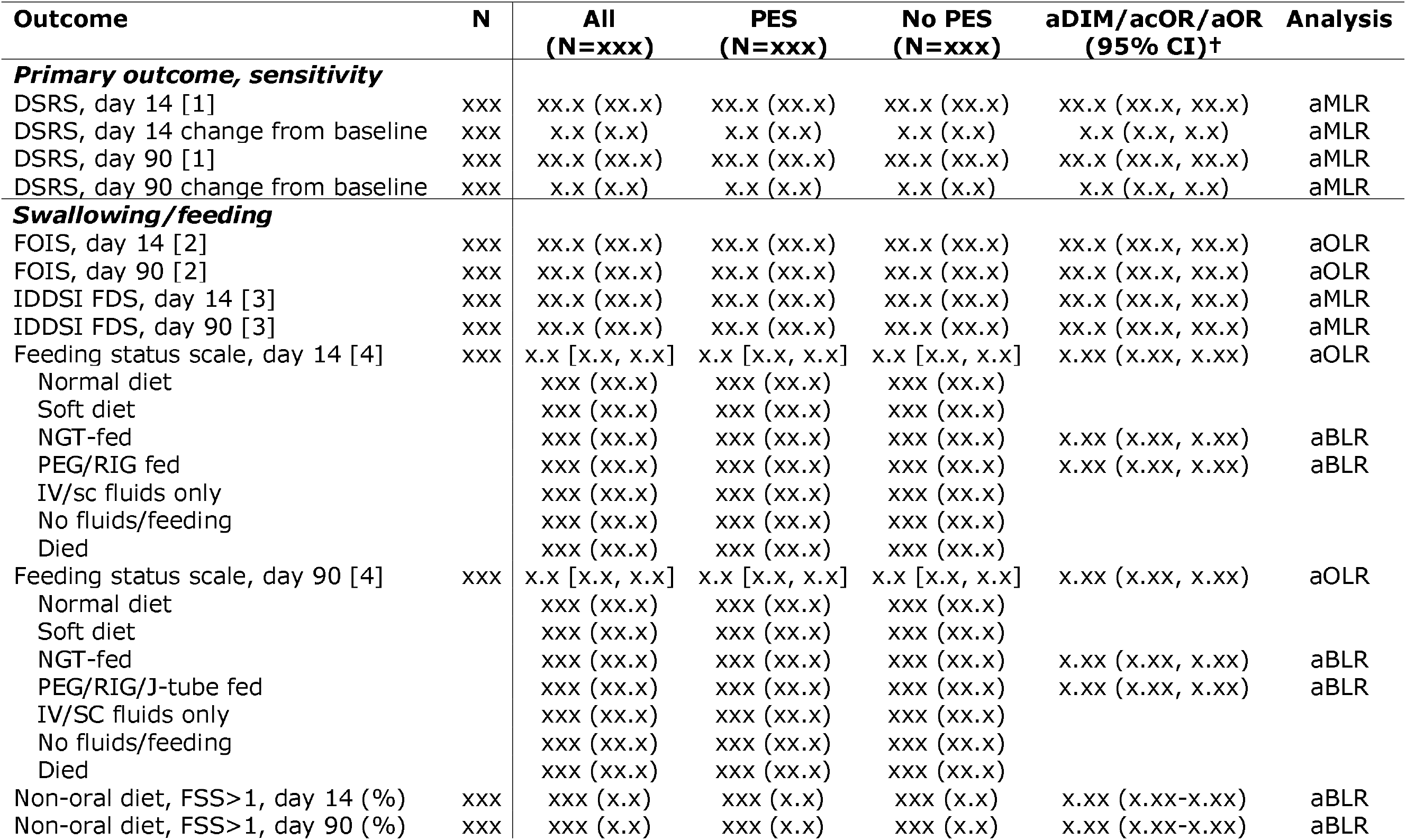
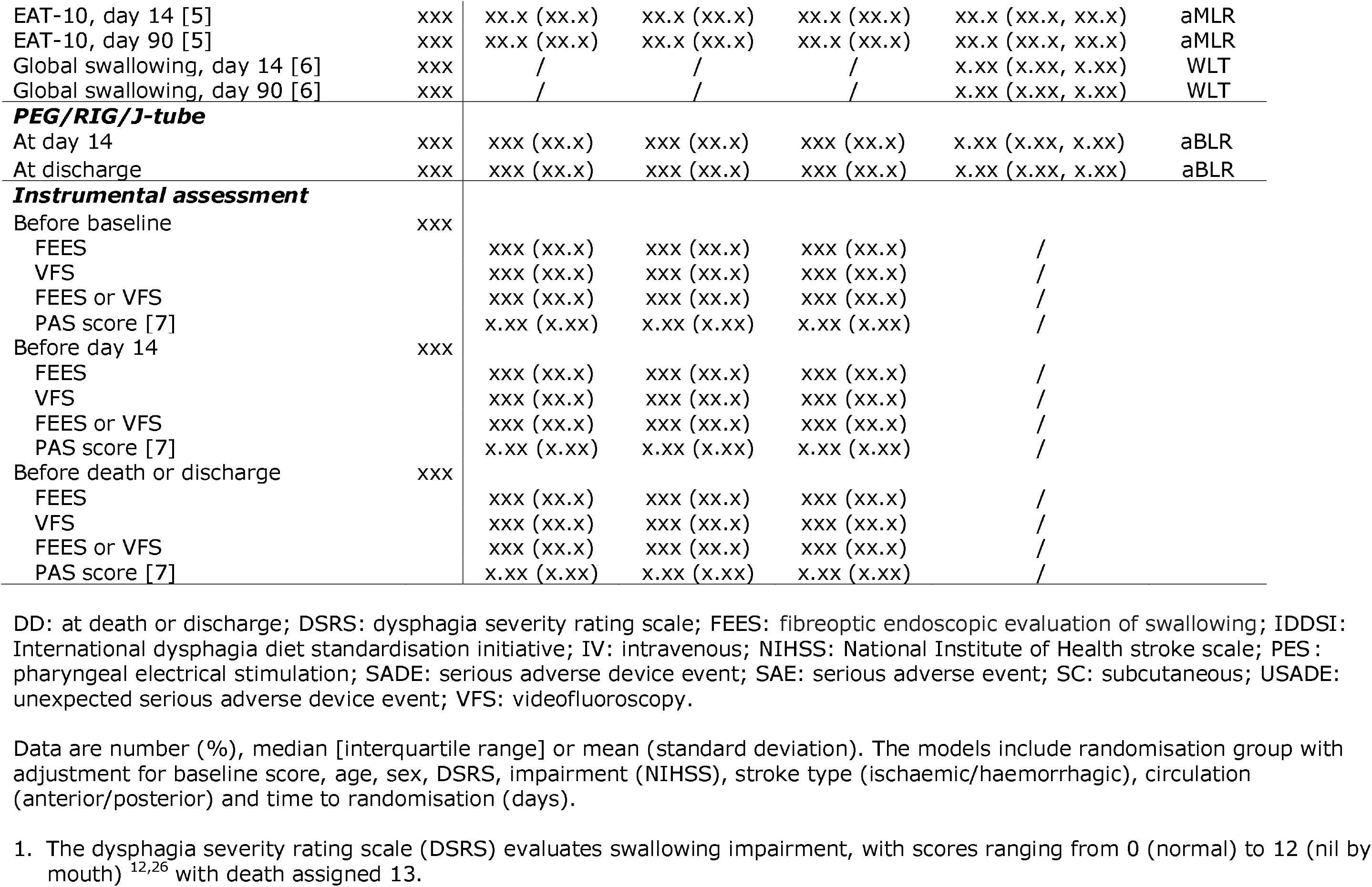
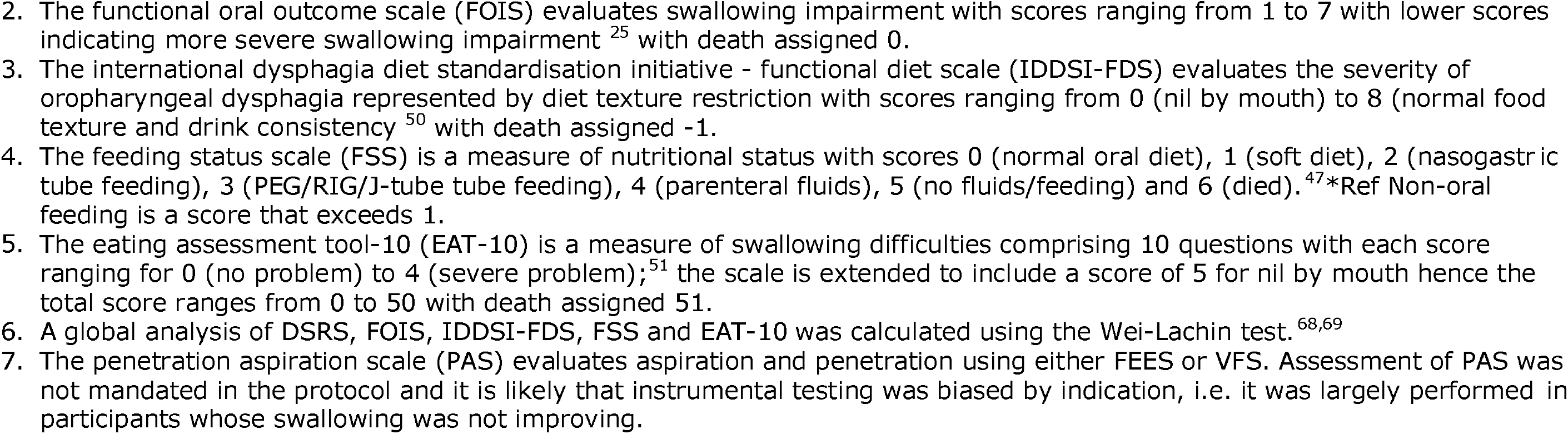
Secondary outcomes related to dysphagia, swallowing and feeding at day 14, discharge and day 90. Analyses used adjusted binary logistic regression (aBLR), adjusted ordinal logistic regression (aOLR) or adjusted multiple linear regression (aMLR).

| Outcome | N | All<br>(N=xxx) | PES<br>(N=xxx) | No PES<br>(N=xxx) | aDIM/acOR/aOR<br>(95% CI)<sup>†</sup> | Analysis |
| --- | --- | --- | --- | --- | --- | --- |
| **Primary outcome, sensitivity** |  |  |  |  |  |  |
| DSRS, day 14 [1] | xxx | xx.x (xx.x) | xx.x (xx.x) | xx.x (xx.x) | xx.x (xx.x, xx.x) | aMLR |
| DSRS, day 14 change from baseline | xxx | x.x (x.x) | x.x (x.x) | x.x (x.x) | x.x (x.x, x.x) | aMLR |
| DSRS, day 90 [1] | xxx | xx.x (xx.x) | xx.x (xx.x) | xx.x (xx.x) | xx.x (xx.x, xx.x) | aMLR |
| DSRS, day 90 change from baseline | xxx | x.x (x.x) | x.x (x.x) | x.x (x.x) | x.x (x.x, x.x) | aMLR |
| **Swallowing/feeding** |  |  |  |  |  |  |
| FOIS, day 14 [2] | xxx | xx.x (xx.x) | xx.x (xx.x) | xx.x (xx.x) | xx.x (xx.x, xx.x) | aOLR |
| FOIS, day 90 [2] | xxx | xx.x (xx.x) | xx.x (xx.x) | xx.x (xx.x) | xx.x (xx.x, xx.x) | aOLR |
| IDDSI FDS, day 14 [3] | xxx | xx.x (xx.x) | xx.x (xx.x) | xx.x (xx.x) | xx.x (xx.x, xx.x) | aMLR |
| IDDSI FDS, day 90 [3] | xxx | xx.x (xx.x) | xx.x (xx.x) | xx.x (xx.x) | xx.x (xx.x, xx.x) | aMLR |
| Feeding status scale, day 14 [4] | xxx | x.x [x.x, x.x] | x.x [x.x, x.x] | x.x [x.x, x.x] | x.xx (x.xx, x.xx) | aOLR |
| Normal diet |  | xxx (xx.x) | xxx (xx.x) | xxx (xx.x) |  |  |
| Soft diet |  | xxx (xx.x) | xxx (xx.x) | xxx (xx.x) |  |  |
| NGT-fed |  | xxx (xx.x) | xxx (xx.x) | xxx (xx.x) | x.xx (x.xx, x.xx) | aBLR |
| PEG/RIG fed |  | xxx (xx.x) | xxx (xx.x) | xxx (xx.x) | x.xx (x.xx, x.xx) | aBLR |
| IV/sc fluids only |  | xxx (xx.x) | xxx (xx.x) | xxx (xx.x) |  |  |
| No fluids/feeding |  | xxx (xx.x) | xxx (xx.x) | xxx (xx.x) |  |  |
| Died |  | xxx (xx.x) | xxx (xx.x) | xxx (xx.x) |  |  |
| Feeding status scale, day 90 [4] | xxx | x.x [x.x, x.x] | x.x [x.x, x.x] | x.x [x.x, x.x] | x.xx (x.xx, x.xx) | aOLR |
| Normal diet |  | xxx (xx.x) | xxx (xx.x) | xxx (xx.x) |  |  |
| Soft diet |  | xxx (xx.x) | xxx (xx.x) | xxx (xx.x) |  |  |
| NGT-fed |  | xxx (xx.x) | xxx (xx.x) | xxx (xx.x) | x.xx (x.xx, x.xx) | aBLR |
| PEG/RIG/J-tube fed |  | xxx (xx.x) | xxx (xx.x) | xxx (xx.x) | x.xx (x.xx, x.xx) | aBLR |
| IV/SC fluids only |  | xxx (xx.x) | xxx (xx.x) | xxx (xx.x) |  |  |
| No fluids/feeding |  | xxx (xx.x) | xxx (xx.x) | xxx (xx.x) |  |  |
| Died |  | xxx (xx.x) | xxx (xx.x) | xxx (xx.x) |  |  |
| Non-oral diet, FSS>1, day 14 (%) | xxx | xxx (x.x) | xxx (x.x) | xxx (x.x) | x.xx (x.xx-x.xx) | aBLR |
| Non-oral diet, FSS>1, day 90 (%) | xxx | xxx (x.x) | xxx (x.x) | xxx (x.x) | x.xx (x.xx-x.xx) | aBLR |

| PhEAST SAP | 11/08/2026 | Version 0.871 |  |  |  |  |
| --- | --- | --- | --- | --- | --- | --- |
| EAT-10, day 14 [5] | xxx | xx.x (xx.x) | xx.x (xx.x) | xx.x (xx.x) | xx.x (xx.x, xx.x) | aMLR |
| EAT-10, day 90 [5] | xxx | xx.x (xx.x) | xx.x (xx.x) | xx.x (xx.x) | xx.x (xx.x, xx.x) | aMLR |
| Global swallowing, day 14 [6] | xxx | / | / | / | x.xx (x.xx, x.xx) | WLT |
| Global swallowing, day 90 [6] | xxx | / | / | / | x.xx (x.xx, x.xx) | WLT |
| **PEG/RIG/J-tube** |  |  |  |  |  |  |
| At day 14 | xxx | xxx (xx.x) | xxx (xx.x) | xxx (xx.x) | x.xx (x.xx, x.xx) | aBLR |
| At discharge | xxx | xxx (xx.x) | xxx (xx.x) | xxx (xx.x) | x.xx (x.xx, x.xx) | aBLR |
| **Instrumental assessment** |  |  |  |  |  |  |
| Before baseline | xxx |  |  |  |  |  |
| FEES |  | xxx (xx.x) | xxx (xx.x) | xxx (xx.x) | / |  |
| VFS |  | xxx (xx.x) | xxx (xx.x) | xxx (xx.x) | / |  |
| FEES or VFS |  | xxx (xx.x) | xxx (xx.x) | xxx (xx.x) | / |  |
| PAS score [7] |  | x.xx (x.xx) | x.xx (x.xx) | x.xx (x.xx) | / |  |
| Before day 14 | xxx |  |  |  |  |  |
| FEES |  | xxx (xx.x) | xxx (xx.x) | xxx (xx.x) | / |  |
| VFS |  | xxx (xx.x) | xxx (xx.x) | xxx (xx.x) | / |  |
| FEES or VFS |  | xxx (xx.x) | xxx (xx.x) | xxx (xx.x) | / |  |
| PAS score [7] |  | x.xx (x.xx) | x.xx (x.xx) | x.xx (x.xx) | / |  |
| Before death or discharge | xxx |  |  |  |  |  |
| FEES |  | xxx (xx.x) | xxx (xx.x) | xxx (xx.x) | / |  |
| VFS |  | xxx (xx.x) | xxx (xx.x) | xxx (xx.x) | / |  |
| FEES or VFS |  | xxx (xx.x) | xxx (xx.x) | xxx (xx.x) | / |  |
| PAS score [7] |  | x.xx (x.xx) | x.xx (x.xx) | x.xx (x.xx) | / |  |

DD: at death or discharge; DSRS: dysphagia severity rating scale; FEES: fiberoptic endoscopic evaluation of swallowing; IDDSI: International dysphagia diet standardisation initiative; IV: intravenous; NIHSS: National Institute of Health stroke scale; PES: pharyngeal electrical stimulation; SADE: serious adverse device event; SAE: serious adverse event; SC: subcutaneous; USADE: unexpected serious adverse device event; VFS: videofluoroscopy.
Data are number (%), median [interquartile range] or mean (standard deviation). The models include randomisation group with adjustment for baseline score, age, sex, DSRS, impairment (NIHSS), stroke type (ischaemic/haemorrhagic), circulation (anterior/posterior) and time to randomisation (days).

**Table S10.**
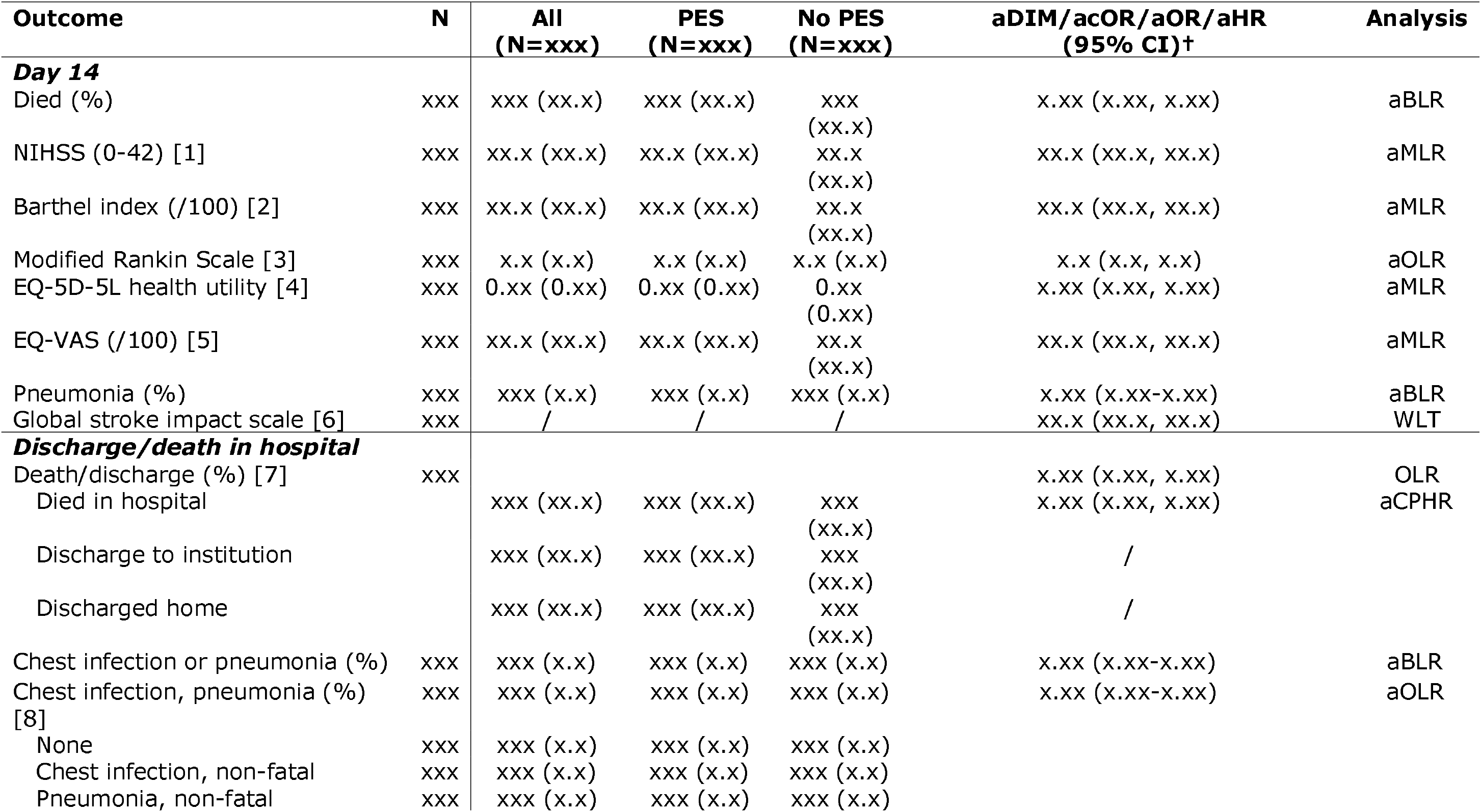
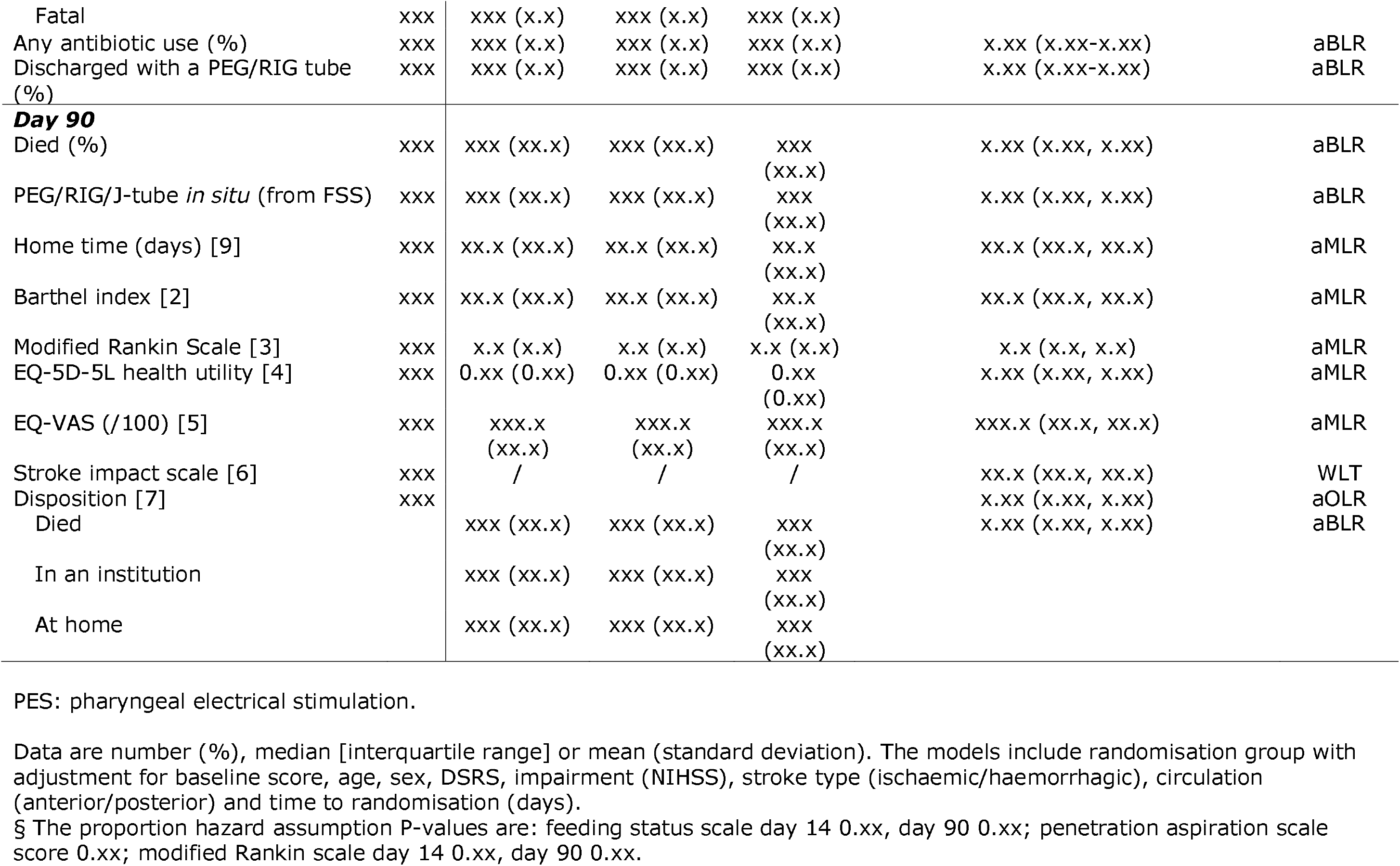
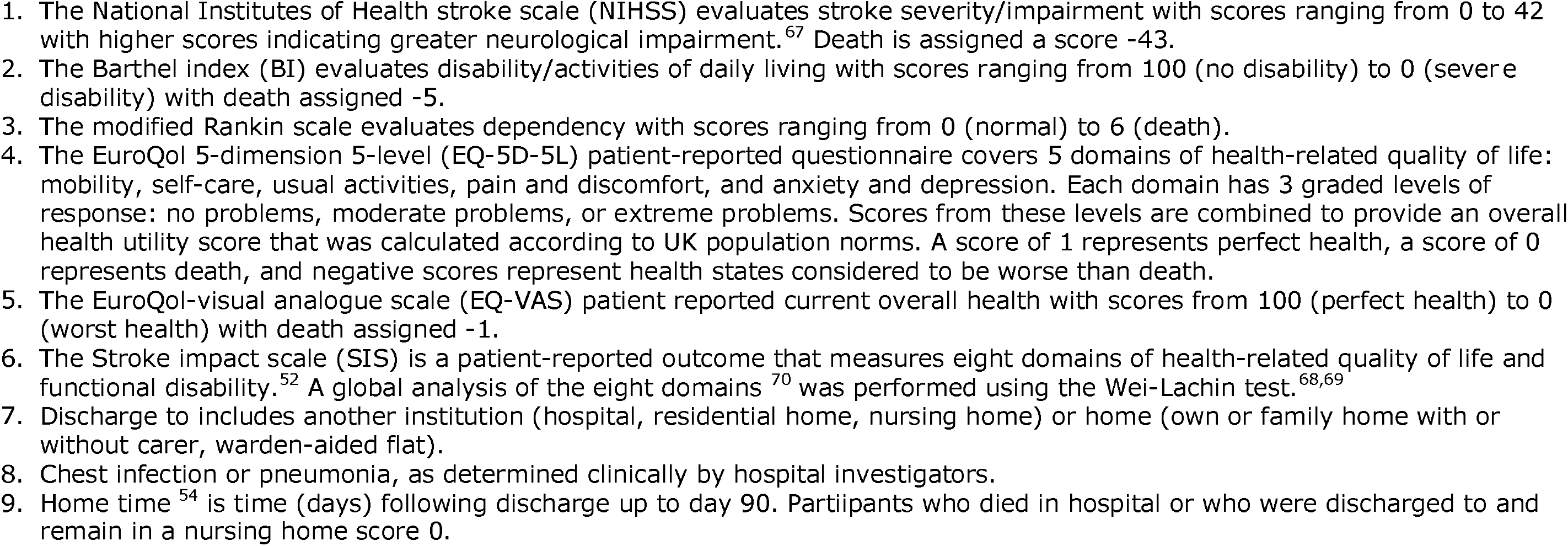
Other secondary outcomes at day 14, discharge or hospital death and day 90. Analyses used adjusted binary logistic regression, adjusted Cox proportional hazards regression, adjusted ordinal logistic regression, adjusted multiple linear regression or Wei-Lachin test.

**Table S11.** Safety end points: causality, events (up to day 14) and fatal (up to day 90). Data are number (frequency), central tendency (95% confidence intervals) and probability.

| End point | N | All | PES Group<br>(N=xxx) | No PES Group<br>(N=xxx) | aDIM/acOR/aOR<br>(95% CI)<sup>†</sup> | Test |
| --- | --- | --- | --- | --- | --- | --- |
| **Events to day 14** |  | xxx | xxx | xxx |  |  |
| Relationship to treatment, all |  |  |  |  | x.xx (x.xx-x.xx) | aOLR |
| Not related (SAE) | xxx | xxx (x.x) | xxx (x.x) | xxx (x.x) |  |  |
| Unlikely to be related (SAE) | xxx | xxx (x.x) | xxx (x.x) | xxx (x.x) |  |  |
| Possibly related (SADE/USADE) | xxx | xxx (x.x) | xxx (x.x) | xxx (x.x) |  |  |
| Probably related (SADE/USADE) | xxx | xxx (x.x) | xxx (x.x) | xxx (x.x) |  |  |
| Definitely related (SADE/USADE) | xxx | xxx (x.x) | xxx (x.x) | xxx (x.x) |  |  |
| **Participants** |  | xxx | xxx | xxx |  |  |
| Event/effect by day 14 |  |  |  |  |  |  |
| SAE | xxx | xxx (xx.x) | xxx (xx.x) | xxx (xx.x) | x.xx (x.xx-x.xx) | BLR |
| Non-fatal | xxx | xxx (xx.x) | xxx (xx.x) | xxx (xx.x) | x.xx (x.xx-x.xx) | BLR |
| Fatal | xxx | xxx (xx.x) | xxx (xx.x) | xxx (xx.x) | x.xx (x.xx-x.xx) | BLR |
| SADE | xxx | xxx (xx.x) | xxx (xx.x) | / | / | / |
| Non-fatal | xxx | xxx (xx.x) | xxx (xx.x) | / | / | / |
| Fatal | xxx | xxx (xx.x) | xxx (xx.x) | / | / | / |
| USADE | xxx | xxx (xx.x) | xxx (xx.x) | / | / | / |
| Non-fatal | xxx | xxx (xx.x) | xxx (xx.x) | / | / | / |
| Fatal | xxx | xxx (xx.x) | xxx (xx.x) | / | / | / |
| Fatal SAE by day 90 | xxx | xxx (xx.x) | xxx (xx.x) | xxx (xx.x) | x.xx (x.xx-x.xx) | CPHR |

SAE: serious adverse event; SADE: serious adverse device effect; USADE: unexpected serious adverse device effect
✕ The proportion hazard assumption P-value is 0.xx.

**Table S12.**
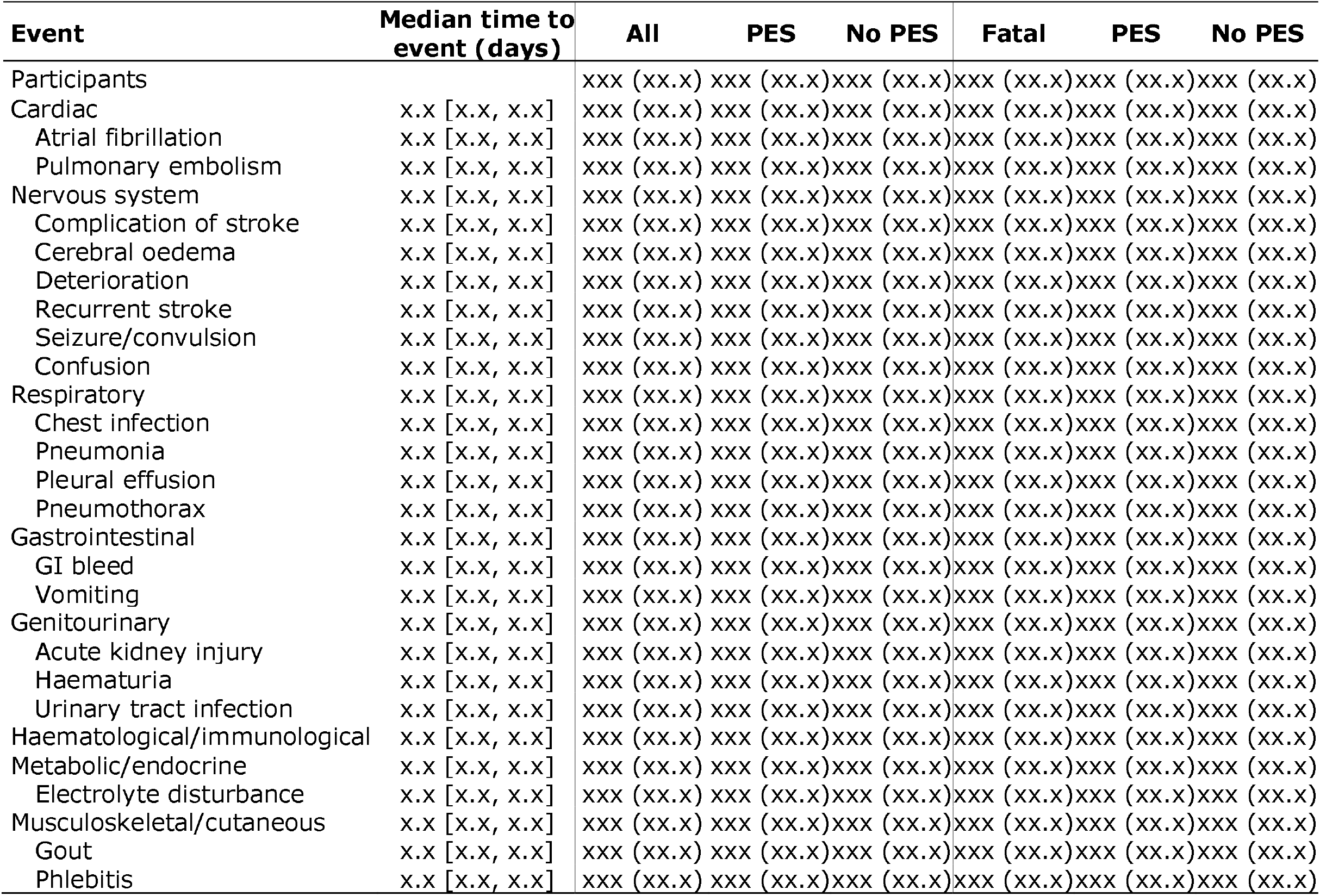
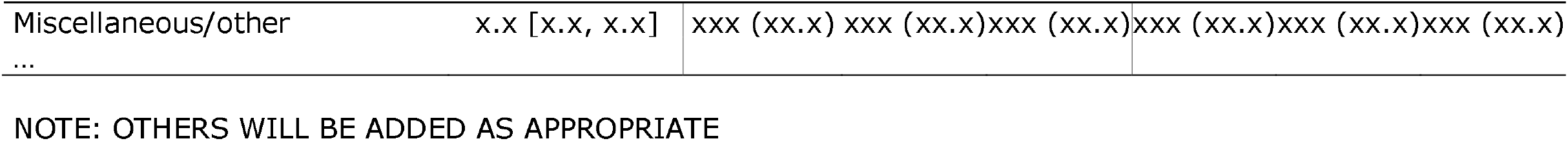
Fatal and non-fatal serious adverse events, serious adverse device events and unexpected serious adverse device events by day 14.

**Table S13.** Protocol violations.

| Violation | All | PES | No PES |
| --- | --- | --- | --- |
| Participants | N | n1 | n2 |
| No consent | xxx (xx.x) | xxx (xx.x) | xxx (xx.x) |
| Ineligible | xxx (xx.x) | xxx (xx.x) | xxx (xx.x) |
| Non stroke dysphagia | xxx (xx.x) | xxx (xx.x) | xxx (xx.x) |
| Pre-stroke dysphagia | xxx (xx.x) | xxx (xx.x) | xxx (xx.x) |
| NIHSS item 1a score 2 or 3 | xxx (xx.x) | xxx (xx.x) | xxx (xx.x) |
| Ongoing/anticipated ventilation/intubation | xxx (xx.x) | xxx (xx.x) | xxx (xx.x) |
| Use of non-trial device for dysphagia | xxx (xx.x) | xxx (xx.x) | xxx (xx.x) |
| Malignant MCA syndrome | xxx (xx.x) | xxx (xx.x) | xxx (xx.x) |
| Pacemaker, cochlear implant, defibrillator | xxx (xx.x) | xxx (xx.x) | xxx (xx.x) |
| On >35% oxygen | xxx (xx.x) | xxx (xx.x) | xxx (xx.x) |
| To be repatriated to a separate organisation | xxx (xx.x) | xxx (xx.x) | xxx (xx.x) |
| To be rehabilitated at a separate organisation | xxx (xx.x) | xxx (xx.x) | xxx (xx.x) |
| Not expected to be at enrolling hospital for 14 days | xxx (xx.x) | xxx (xx.x) | xxx (xx.x) |
| Not expected to tolerate PES catheter | xxx (xx.x) | xxx (xx.x) | xxx (xx.x) |
| To be transferred to a non-participating site | xxx (xx.x) | xxx (xx.x) | xxx (xx.x) |
| Known to be pregnant at enrolment | xxx (xx.x) | xxx (xx.x) | xxx (xx.x) |
| In another trial with no co-enrolment agreement | xxx (xx.x) | xxx (xx.x) | xxx (xx.x) |
| On palliative care | xxx (xx.x) | xxx (xx.x) | xxx (xx.x) |
| Known pharyngeal pouch | xxx (xx.x) | xxx (xx.x) | xxx (xx.x) |
| Dysphagia expected to resolve rapidly | xxx (xx.x) | xxx (xx.x) | xxx (xx.x) |
| EDAR at enrolment | xxx (xx.x) | xxx (xx.x) | xxx (xx.x) |
| PES started $\geq 3$ days after randomisation | xxx (xx.x) | xxx (xx.x) | xxx (xx.x) |
| Non-reported primary outcome | xxx (xx.x) | xxx (xx.x) | xxx (xx.x) |
| Early/late reporting of primary outcome | xxx (xx.x) | xxx (xx.x) | xxx (xx.x) |
| Non-reported SAE, SADE or USADE | xxx (xx.x) | xxx (xx.x) | xxx (xx.x) |
| ... | xxx (xx.x) | xxx (xx.x) | xxx (xx.x) |
| Total number of participants with $\geq 1$ violation | xxx (xx.x) | xxx (xx.x) | xxx (xx.x) |

† Participants may have one or more violations
NOTE: OTHERS WILL BE ADDED AS APPROPRIATE

**Table S14.**
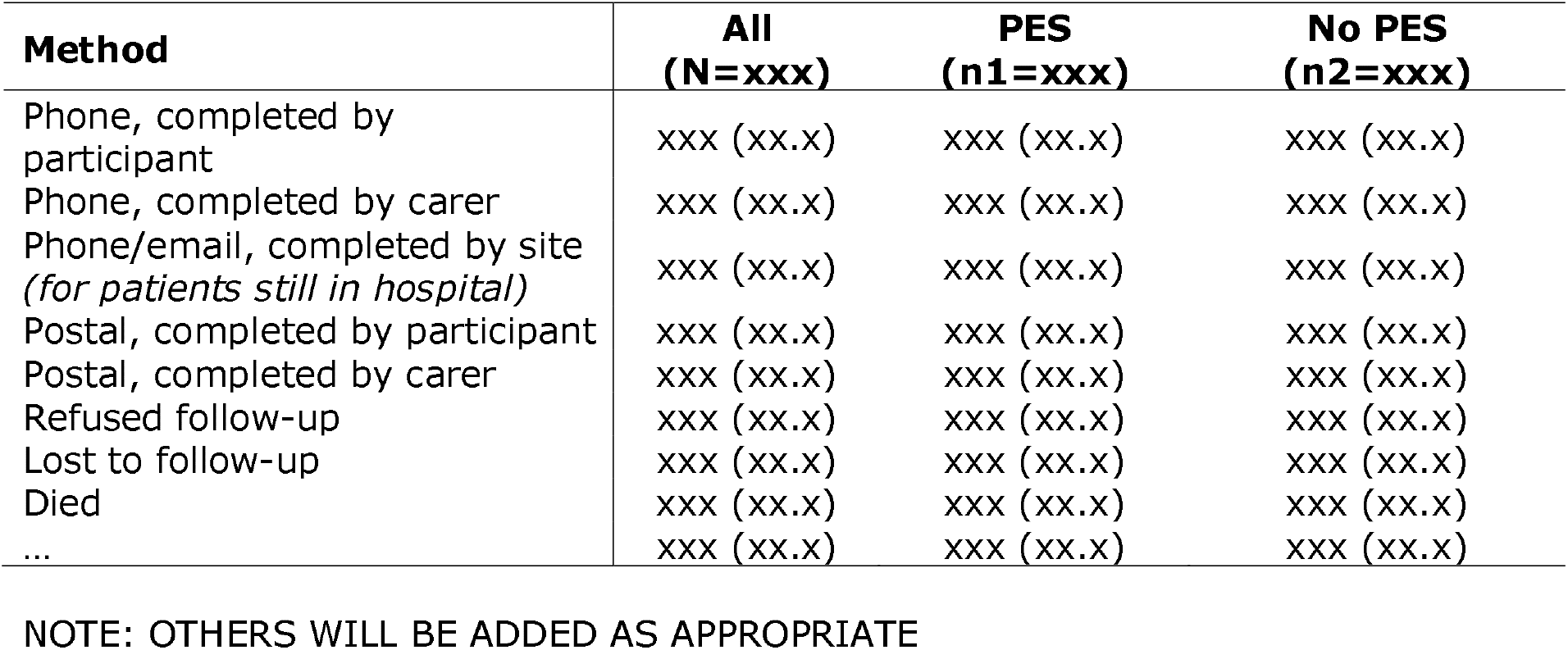
Method of follow-up at day 90.

| **Method** | **All**<br>**(N=xxx)** | **PES**<br>**(n1=xxx)** | **No PES**<br>**(n2=xxx)** |
| --- | --- | --- | --- |
| Phone, completed by participant | xxx (xx.x) | xxx (xx.x) | xxx (xx.x) |
| Phone, completed by carer | xxx (xx.x) | xxx (xx.x) | xxx (xx.x) |
| Phone/email, completed by site<br>(for patients still in hospital) | xxx (xx.x) | xxx (xx.x) | xxx (xx.x) |
| Postal, completed by participant | xxx (xx.x) | xxx (xx.x) | xxx (xx.x) |
| Postal, completed by carer | xxx (xx.x) | xxx (xx.x) | xxx (xx.x) |
| Refused follow-up | xxx (xx.x) | xxx (xx.x) | xxx (xx.x) |
| Lost to follow-up | xxx (xx.x) | xxx (xx.x) | xxx (xx.x) |
| Died | xxx (xx.x) | xxx (xx.x) | xxx (xx.x) |
| ... | xxx (xx.x) | xxx (xx.x) | xxx (xx.x) |

NOTE: OTHERS WILL BE ADDED AS APPROPRIATE

**Table S15.**
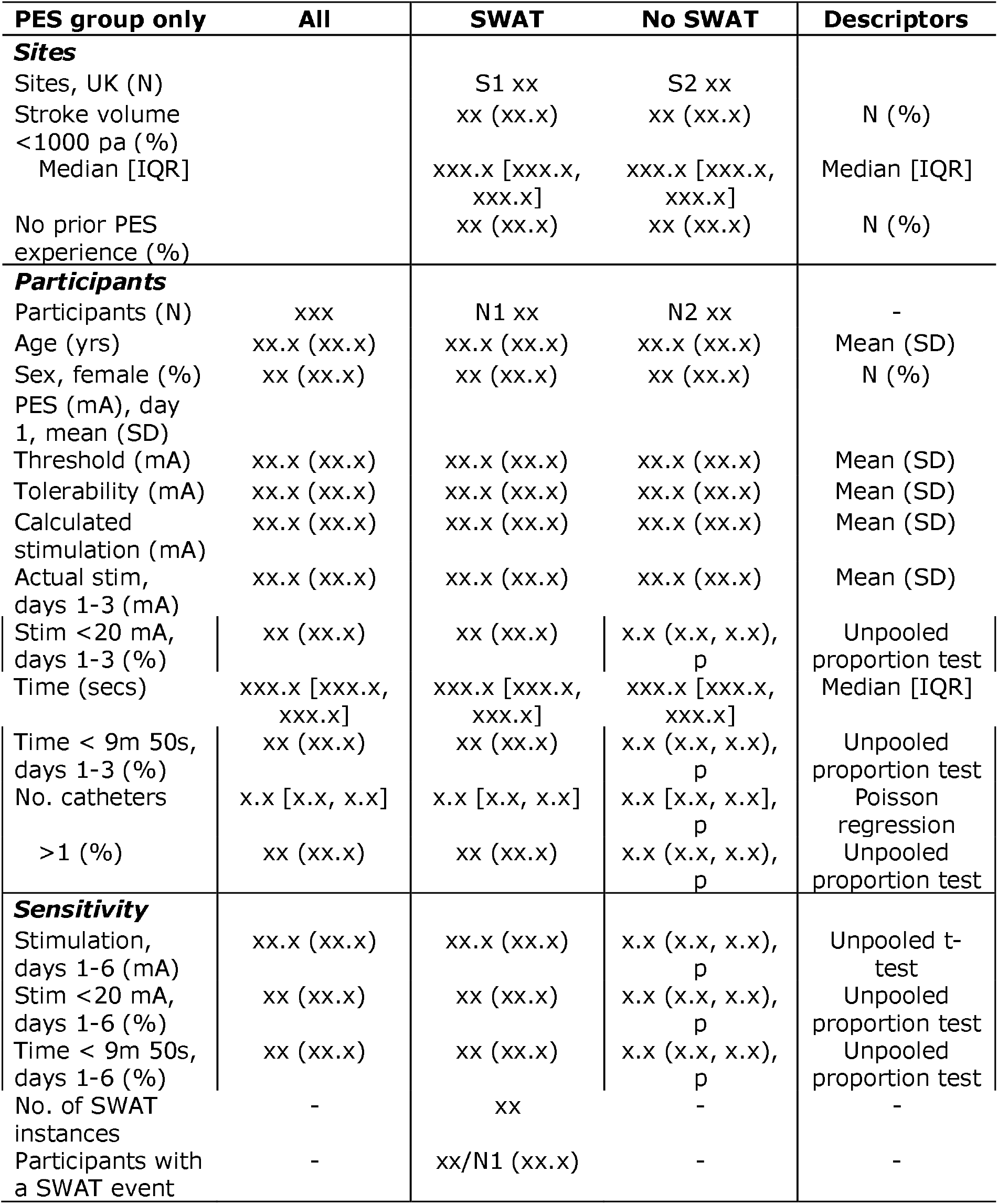
Analysis of the study within a trial (SWAT) assessment of undertreatment in participants randomised to PES.

| **PES group only** | **All** | **SWAT** | **No SWAT** | **Descriptors** |
| --- | --- | --- | --- | --- |
| **Sites** |  |  |  |  |
| Sites, UK (N) |  | S1 xx | S2 xx |  |
| Stroke volume<br><1000 pa (%) |  | xx (xx.x) | xx (xx.x) | N (%) |
| Median [IQR] |  | xxx.x [xxx.x,<br>xxx.x] | xxx.x [xxx.x,<br>xxx.x] | Median [IQR] |
| No prior PES<br>experience (%) |  | xx (xx.x) | xx (xx.x) | N (%) |
| **Participants** |  |  |  |  |
| Participants (N) | xxx | N1 xx | N2 xx | - |
| Age (yrs) | xx.x (xx.x) | xx.x (xx.x) | xx.x (xx.x) | Mean (SD) |
| Sex, female (%) | xx (xx.x) | xx (xx.x) | xx (xx.x) | N (%) |
| PES (mA), day<br>1, mean (SD) |  |  |  |  |
| Threshold (mA) | xx.x (xx.x) | xx.x (xx.x) | xx.x (xx.x) | Mean (SD) |
| Tolerability (mA) | xx.x (xx.x) | xx.x (xx.x) | xx.x (xx.x) | Mean (SD) |
| Calculated<br>stimulation (mA) | xx.x (xx.x) | xx.x (xx.x) | xx.x (xx.x) | Mean (SD) |
| Actual stim,<br>days 1-3 (mA) | xx.x (xx.x) | xx.x (xx.x) | xx.x (xx.x) | Mean (SD) |
| Stim <20 mA,<br>days 1-3 (%) | xx (xx.x) | xx (xx.x) | x.x (x.x, x.x),<br>p | Unpooled<br>proportion test |
| Time (secs) | xxx.x [xxx.x,<br>xxx.x] | xxx.x [xxx.x,<br>xxx.x] | xxx.x [xxx.x,<br>xxx.x] | Median [IQR] |
| Time < 9m 50s,<br>days 1-3 (%) | xx (xx.x) | xx (xx.x) | x.x (x.x, x.x),<br>p | Unpooled<br>proportion test |
| No. catheters | x.x [x.x, x.x] | x.x [x.x, x.x] | x.x [x.x, x.x],<br>p | Poisson<br>regression |
| >1 (%) | xx (xx.x) | xx (xx.x) | x.x (x.x, x.x),<br>p | Unpooled<br>proportion test |
| **Sensitivity** |  |  |  |  |
| Stimulation,<br>days 1-6 (mA) | xx.x (xx.x) | xx.x (xx.x) | x.x (x.x, x.x),<br>p | Unpooled t-test |
| Stim <20 mA,<br>days 1-6 (%) | xx (xx.x) | xx (xx.x) | x.x (x.x, x.x),<br>p | Unpooled<br>proportion test |
| Time < 9m 50s,<br>days 1-6 (%) | xx (xx.x) | xx (xx.x) | x.x (x.x, x.x),<br>p | Unpooled<br>proportion test |
| No. of SWAT<br>instances | - | xx | - | - |
| Participants with<br>a SWAT event | - | xx/N1 (xx.x) | - | - |

**Table S16.** Device deficiencies as reported by site investigators. Data apply only to PES group and are number (%) or mean (standard deviation) with [range].

| **PES group only** | **N (%)** |
| --- | --- |
| ***Base-station*** |  |
| Software issue - system showing incomplete treatments and unable to increase threshold/tolerance | xxx (xx.x) |
| ... | xxx (xx.x) |
| ***Catheter insertion*** |  |
| Guidewire not removed (user error) | xxx (xx.x) |
| Treatment attempted but catheter in incorrect place (user error) | xxx (xx.x) |
| ... | xxx (xx.x) |
| ***Catheter*** |  |
| Catheter securing plaster found in catheter box | xxx (xx.x) |
| Catheter losing contact despite being in position | xxx (xx.x) |
| Naso-gastric tube slippage despite clip in place | xxx (xx.x) |
| Blocked catheter, needed to be removed | xxx (xx.x) |
| ... | xxx (xx.x) |

NOTE: OTHERS WILL BE ADDED AS APPROPRIATE

**Figure S1. CONSORT diagram.**

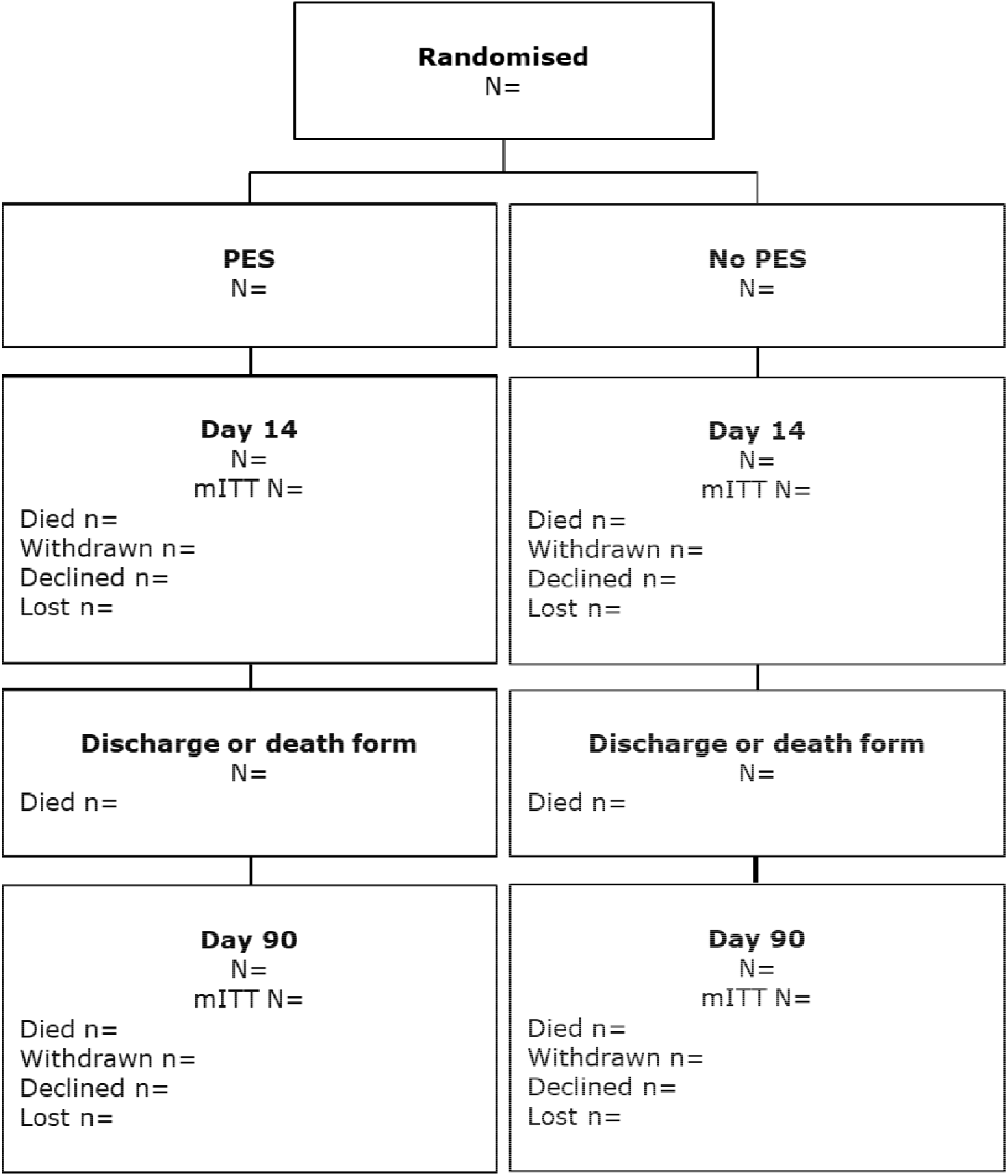

Note: Sites variably screened all stroke admissions, only those referred to SLTs with dysphagia or just those where patients were given an information sheet. As a results, screening data are incomplete and an underestimate; of those reported, XXX patients were screened.

**Figure S2. Sankey diagram of change in DSRS between days 0-14 and days 14 - 90 shown by treatment group.**

**Figure S3. Swallowing impairment at 14 and 90 days in the two groups according to scores on the functional oral intake scale.**

| Shift | Score | 0 | 1 | 2 | 3 | 4 | 5 | 6 | 7 |
| --- | --- | --- | --- | --- | --- | --- | --- | --- | --- |
|  | Day |  |  |  |  |  |  |  |  |
| PES | 00 | 0 | xx | xx | xx | xx | xx | xx | xx |
|  | 14 | xx | xx | xx | xx | xx | xx | xx | xx |
|  | 90 | xx | xx | xx | xx | xx | xx | xx | xx |
| No PES | 90 | xx | xx | xx | xx | xx | xx | xx | xx |
|  | 14 | xx | xx | xx | xx | xx | xx | xx | xx |
|  | 00 | 0 | xx | xx | xx | xx | xx | xx | xx |

The functional oral outcome scale (FOIS) evaluates swallowing impairment with scores ranging from 1 to 7 with lower scores indicating more severe swallowing impairment;^25^ inclusion criteria were: 0 - death, 1 - nil by mouth, 2 - tube dependent with minimal attempts of food or liquid, 3 - tube dependent with consistent oral intake of food or liquid.

**Figure S4. Subgroup analyses of the primary outcome within the PES group.**

Shown are the difference in means for swallowing impairment at day 90 in pre­specified subgroups of stimulation levels in milliamps. The widths of the confidence intervals have not been adjusted for multiplicity and should not be used in place of hypothesis testing. The analysis follows that of PHADER.^16^

- Threshold, average: <10, 10-13, >13
- Stimulation, average: <30, 30-36, >36
- Tolerability, average: <35, 35-45, >45
- Delta, tolerability - threshold, average: <24, 24-33, >33

### SUPPLEMENT 4

**BASELINE PAPER**

**Title**

Baseline characteristics of participants enrolled into the pharyngeal electrical stimulation for acute stroke dysphagia trial (PhEAST) trial.

**Authors**

TBD

**Aim**

Describe the baseline characteristics of participants enrolled into the PhEAST trial.

**Participants**

All in PhEAST

**Outcomes**

None.

**Analyses**

Descriptive statistics: number (%), median [interquartile range] or mean (standard deviation).

**Table S4-1.**
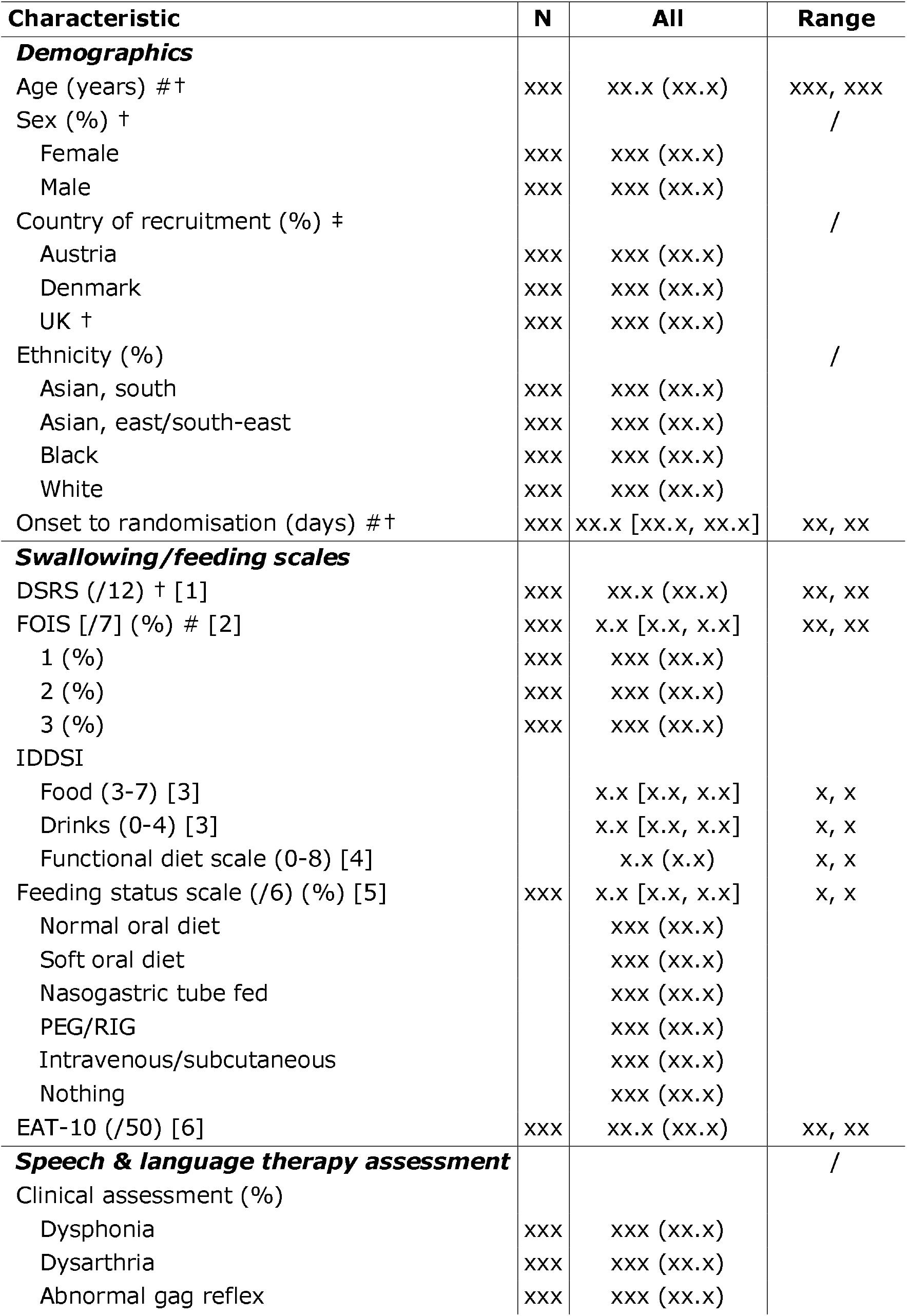
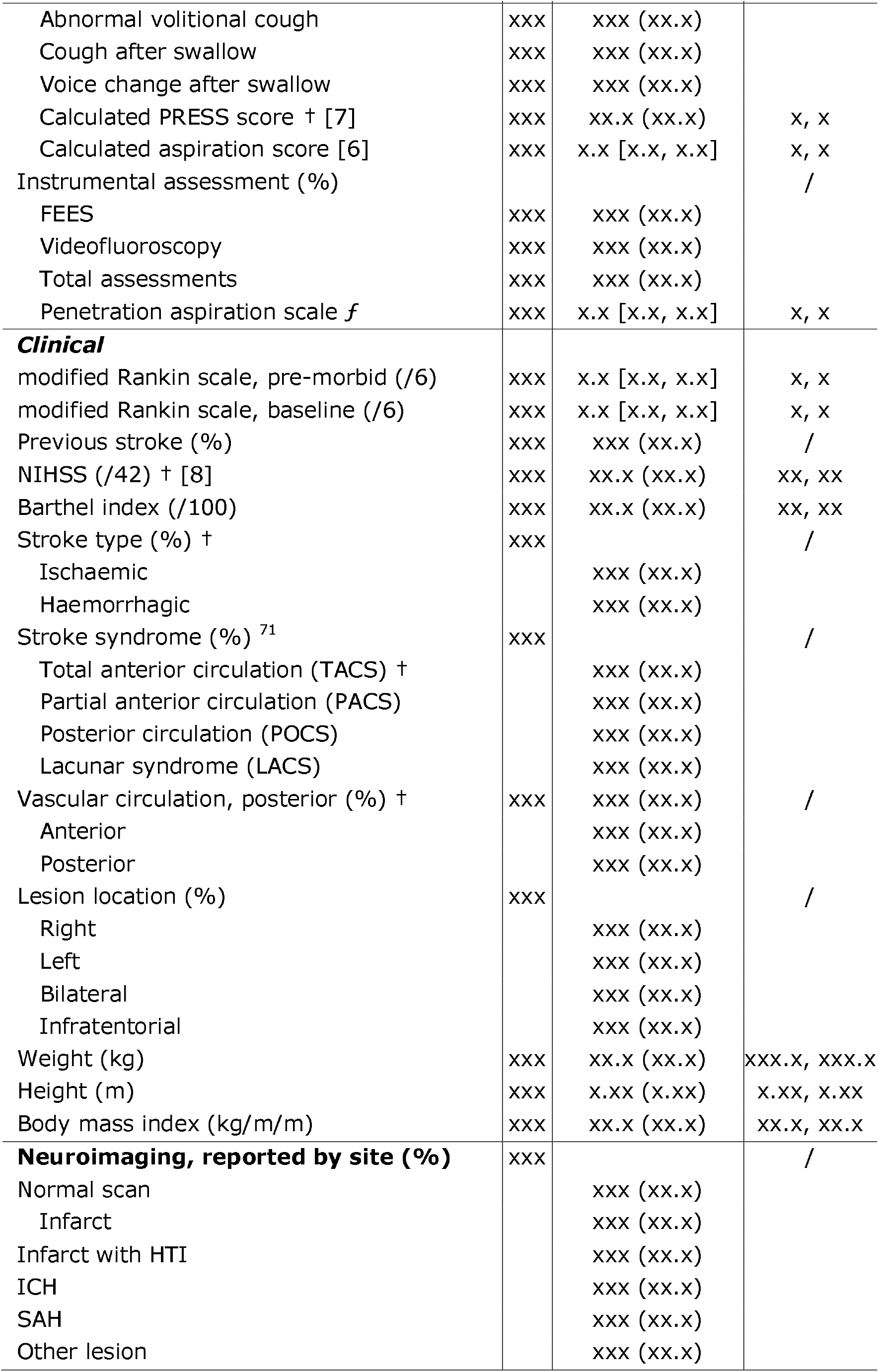
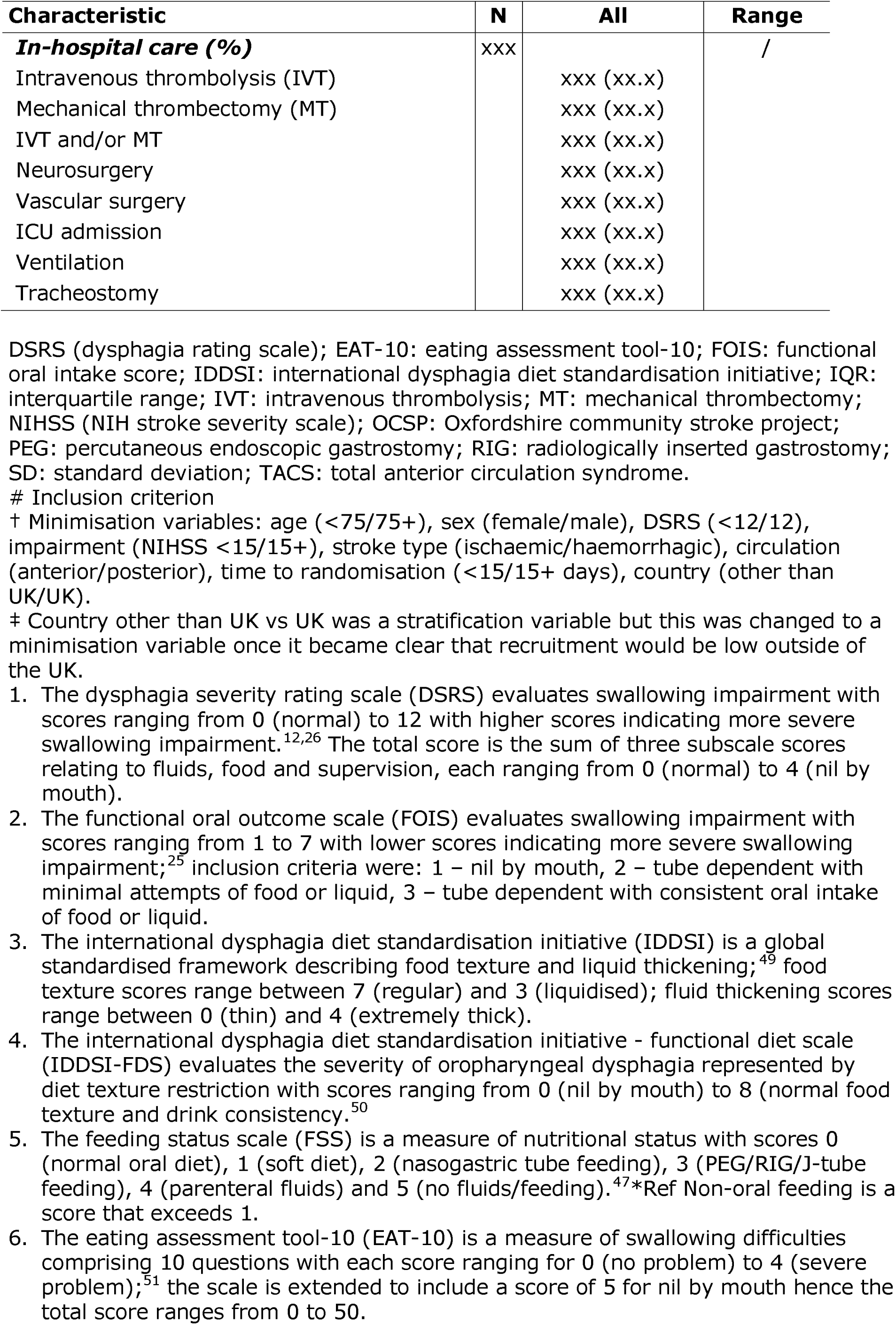
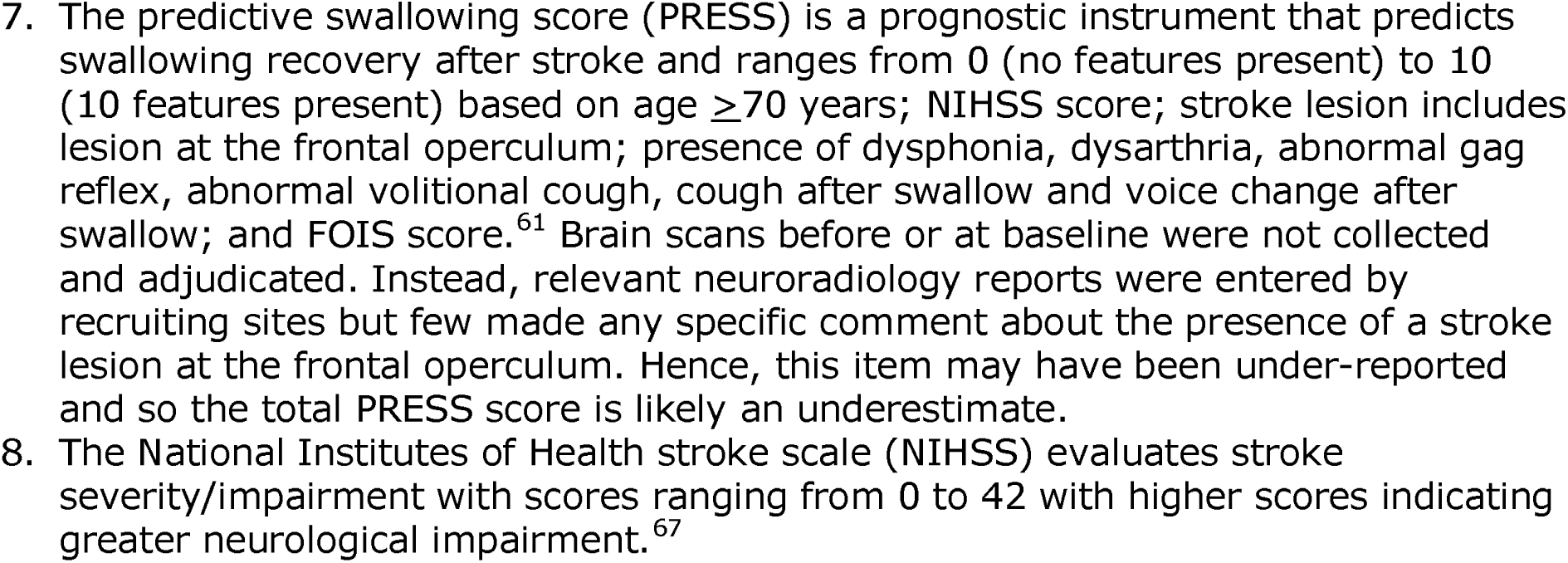
Baseline characteristics for all recorded variables at baseline excluding cognition. Data are number (%), median [interquartile range] or mean (standard deviation) with minimum/maximum.

**Table S4-2.** Sources of consent accounting for country of randomisation and type of consent.

| Variable | All |
| --- | --- |
| Participants with data | N |
| Participant consent | xxx (xx.x) |
| Consultee declaration: England, Denmark, Northern Ireland, Wales | xxx (xx.x) |
| Personal legal representative: Austria, Scotland | xxx (xx.x) |
| Independent physician | xxx (xx.x) |

**Table S4-3.** Co-enrolment into another trial.

| Variable | PhEAST order | N |
| --- | --- | --- |
| Participants |  | xxx |
| ENOS-2 (ISRCTN17654248) | Second | xxx (xx.x) |
| MAPS-2 (ISRCTN40512746) | Second | xxx (xx.x) |
| RECAST-3 (ISRCTN63231313) | Second | xxx (xx.x) |
| TICH-3 (ISRCTN97695350) | Second | xxx (xx.x) |
| ProFATE <sup>72</sup> | Second | xxx (xx.x) |
| GEKO (ISRCTN11175235) | Second | xxx (xx.x) |
| ... |  |  |
| ... |  |  |

**Table S4-4.** Univariate correlations between baseline characteristics for age, sex, swallowing severity (DSRS), stroke severity (NIHSS), stroke type (ischaemic, haemorrhagic), time for onset to randomisation (OTR), and year of randomisation. Correlations by point biserial or Spearman’s tests; data are regression coefficient (P value).

|  | Age | Sex | DSRS | FOIS | IDDSI-RDS | EAT-10 | NIHSS | IS/ICH | OTR | Year |
| --- | --- | --- | --- | --- | --- | --- | --- | --- | --- | --- |
| Age | X | r (p) | r (p) | r (p) | r (p) | r (p) | r (p) | r (p) | r (p) | r (p) |
| Sex | r (p) | X | r (p) | r (p) | r (p) | r (p) | r (p) | r (p) | r (p) | r (p) |
| DSRS | r (p) | r (p) | X | r (p) | r (p) | r (p) | r (p) | r (p) | r (p) | r (p) |
| FOIS | r (p) | r (p) | r (p) | X | r (p) | r (p) | r (p) | r (p) | r (p) | r (p) |
| IDDSI-RDS | r (p) | r (p) | r (p) | r (p) | X | r (p) | r (p) | r (p) | r (p) | r (p) |
| EAT-10 | r (p) | r (p) | r (p) | r (p) | r (p) | X | r (p) | r (p) | r (p) | r (p) |
| NIHSS | r (p) | r (p) | r (p) | r (p) | r (p) | r (p) | X | r (p) | r (p) | r (p) |
| IS/ICH | r (p) | r (p) | r (p) | r (p) | r (p) | r (p) | r (p) | X | r (p) | r (p) |
| OTR | r (p) | r (p) | r (p) | r (p) | r (p) | r (p) | r (p) | r (p) | X | r (p) |
| Year | r (p) | r (p) | r (p) | r (p) | r (p) | r (p) | r (p) | r (p) | r (p) | X |

**Figure 1. Cumulative recruitment of participants and sites during the trial.**

Nb: Place the site line below the participant line.

**Figure 2. Monthly recruitment of participants and sites during the trial.**

Nb: Bars for each month

**Figure 3. Histogram of time from stroke onset to randomisation (days).**

Nb: Bars for each day 2-31.

**Figure 4. Histogram of recruitment by sites.**

Nb: Bars for each site in order of recruitment with sites identified as A, B, C…

### SUPPLEMENT 5

**Title**

Cost effectiveness of pharyngeal electrical stimulation for post stroke dysphagia as compared to usual care. Data from the PhEAST trial.

**Authors**

Cristina Roadevin, Philip M Bath, Marilyn James.

**Aim**

Evaluate the cost effectiveness of the PhEAST trial intervention (PES) compared to usual care.

**Participants**

All in PhEAST

**Outcomes**

Primary: Health-related quality of life (HRQoL) for participant: EuroQoL-5D-5L (EQ-5D-5L) administered at baseline, and days: 14, 90.

Secondary: *Cristina

**Analyses**

Descriptive statistics: number (%), median [interquartile range] or mean (standard deviation).

Cost-effectiveness assessed through incremental cost-effectiveness ratios (ICERs) and incremental net monetary benefits (INMBs).

Exemplar tables follow.

**Table S5-1.** Estimated and imputed average QALYs per patient at 12 months. Data are mean (standard error, SE).

|  | **PES**<br>**Mean (SE)** | **Usual Care**<br>**Mean (SE)** | **Difference in**<br>**means between**<br>**groups (SE)** |
| --- | --- | --- | --- |
| QALYs | xx.x (xx.x) | xx.x (xx.x) | xx.x (xx.x) |

SE: standard error

**Table S5-2.** Average costs per patient and difference in means between groups at 12 months.

|  | **PES + usual care (n=)** |  | **Usual Care (n=)** |  | **Difference in means between groups (£)** |
| --- | --- | --- | --- | --- | --- |
|  | Mean (£) | SE | Mean (£) | SE | Mean (SE) |
| Health service perspective costs | xx.x | xx.x | xx.x | xx.x | xx.x |
| Intervention cost | xx.x | xx.x | xx.x | xx.x | xx.x |
| Inpatient services | xx.x | xx.x | xx.x | xx.x | xx.x |
| Outpatient services | xx.x | xx.x | xx.x | xx.x | xx.x |
| Primary and community care | xx.x | xx.x | xx.x | xx.x | xx.x |
| Medication | xx.x | xx.x | xx.x | xx.x | xx.x |
| Aids and Equipment | xx.x | xx.x | xx.x | xx.x | xx.x |
| Broader Costs | xx.x | xx.x | xx.x | xx.x | xx.x |
| Informal care | xx.x | xx.x | xx.x | xx.x | xx.x |
| Aids and Equipment | xx.x | xx.x | xx.x | xx.x | xx.x |
| Total costs | xx.x | xx.x | xx.x | xx.x | xx.x |
| Health Service Perspective Costs | xx.x | xx.x | xx.x | xx.x | xx.x |
| Societal Perspective Costs | xx.x | xx.x | xx.x | xx.x | xx.x |

**Table S5-3.** Cost Effectiveness Results.

|  | **Incremental costs (95% CI)** | **Incremental QALYs (95% CI)** | **ICER (95% CI)** |
| --- | --- | --- | --- |
| NHS perspective | xx.x (xx.x, xx.x) | xx.x (xx.x, xx.x) | xx.x (xx.x, xx.x) |
| Societal perspective | xx.x (xx.x, xx.x) | xx.x (xx.x, xx.x) | xx.x (xx.x, xx.x) |

**Table S5-4.** Net Monetary Benefit Results.

| **WTP thresholds** | **£20,000** | **£30,000** | **£40,000** | **£50,000** | **£60,000** | **£100,000** |
| --- | --- | --- | --- | --- | --- | --- |
| NMB values | xx.x | xx.x | xx.x | xx.x | xx.x | xx.x |

### SUPPLEMENT 6

**SWALLOWING/DYSPHAGIA UP TO ONE YEAR**

**Title**

Trajectory of swallowing over 1 year in patients with severe ischaemic or haemorrhagic stroke and dysphagia. Data from the PhEAST trial

**Authors**

TBD

**Aim**

Describe the trajectory of swallowing at 5 time points over 1 year in patients with severe ischaemic or haemorrhagic stroke and dysphagia defined by tube-feeding at baseline. Compare swallowing impairment by PES versus no PES

**Participants**

All in PhEAST

**Outcomes**

Primary: DSRS.

Secondary: At all timepoints - FOIS, FSS, IDDSI, IDDSI-FDS, NGT or PEG in situ, death, discharge disposition.

Others: mRS, Barthel, EQ-5D, EQ-VAS, global outcome

**Analyses**

Descriptive statistics: number (%), median [interquartile range] or mean (standard deviation).

Comparison by adjusted repeated measures regression.

Data will be tabulated in all patients whether alive for not, and in alive patients only. The analyses will follow those in the main paper but cover outcomes out to one year. Analyses present in the main paper but not of relevance at one year, e.g. PES stimulation levels, will not be re-presented.

**Table S6-1.** Baseline characteristics by baseline FOIS score.

| Characteristic | Statistic | N | All<br>(N=xxx) | FOIS=1<br>(N=xxx) | FOIS=2<br>(N=xxx) | FOIS=3<br>(N=xxx) | 2p trend |
| --- | --- | --- | --- | --- | --- | --- | --- |
| **Demographics** |  |  |  |  |  |  |  |
| Age (years) #† | Mean (SD) | xxx | xxx (xx.x) | xxx (xx.x) | xxx (xx.x) | xxx (xx.x) | Jonckheere test |
| Sex, female (%) | n (%) | xxx | xxx (xx.x) | xxx (xx.x) | xxx (xx.x) | xxx (xx.x) | CAT 3x2 |
| Country, other than UK | n (%) | xxx | xxx (xx.x) | xxx (xx.x) | xxx (xx.x) | xxx (xx.x) | CAT 3x2 |
| Ethnicity, other than white (%) | n (%) | xxx | xxx (xx.x) | xxx (xx.x) | xxx (xx.x) | xxx (xx.x) | CAT 3x2 |
| Onset to randomisation (days) #† | Median [IQR] | xxx | x.x [x.x, x.x] | x.x [x.x, x.x] | x.x [x.x, x.x] | x.x [x.x, x.x] | Jonckheere test |
| **Swallowing** |  |  |  |  |  |  |  |
| DSRS (/12) † | Mean (SD) | xxx | xx.x (xx.x) | xx.x (xx.x) | xx.x (xx.x) | xx.x (xx.x) | Jonckheere test |
| FOIS [/7] # | Median [IQR] | xxx | x.x [x.x, x.x] | x.x [x.x, x.x] | x.x [x.x, x.x] | x.x [x.x, x.x] | Jonckheere test |
| IDDSI |  |  |  |  |  |  |  |
| Food (3-7) | Median [IQR] | xxx | x.x [x.x, x.x] | x.x [x.x, x.x] | x.x [x.x, x.x] | x.x [x.x, x.x] | Jonckheere test |
| Drinks (0-4) | Median [IQR] | xxx | x.x [x.x, x.x] | x.x [x.x, x.x] | x.x [x.x, x.x] | x.x [x.x, x.x] | Jonckheere test |
| Functional diet scale (0-8) | Mean (SD) | xxx | xx.x (xx.x) | xx.x (xx.x) | xx.x (xx.x) | xx.x (xx.x) | Jonckheere test |
| Feeding status scale, FSS>1 (%) | n (%) | xxx | xxx (xx.x) | xxx (xx.x) | xxx (xx.x) | xxx (xx.x) | CAT 3x2 |
| EAT-10 (/50) | Mean (SD) | xxx | xx.x (xx.x) | xx.x (xx.x) | xx.x (xx.x) | xx.x (xx.x) | Jonckheere test |
| **Clinical** |  |  |  |  |  |  |  |
| Modified Rankin scale, premorbid [/6] | Median [IQR] | xxx | x.x [x.x, x.x] | x.x [x.x, x.x] | x.x [x.x, x.x] | x.x [x.x, x.x] | Jonckheere test |
| Modified Rankin scale, baseline [/6] | Median [IQR] | xxx | x.x [x.x, x.x] | x.x [x.x, x.x] | x.x [x.x, x.x] | x.x [x.x, x.x] | Jonckheere test |
| Previous stroke | n (%) | xxx | xxx (xx.x) | xxx (xx.x) | xxx (xx.x) | xxx (xx.x) | CAT 3x2 |
| NIHSS (/42) † | Mean (SD) | xxx | xx.x (xx.x) | xx.x (xx.x) | xx.x (xx.x) | xx.x (xx.x) | Jonckheere test |
| Barthel index (/100) | Mean (SD) | xxx | xxx (xx.x) | xxx (xx.x) | xxx (xx.x) | xxx (xx.x) | Jonckheere test |
| Stroke type, haemorrhage (%) † | n (%) | xxx | xxx (xx.x) | xxx (xx.x) | xxx (xx.x) | xxx (xx.x) | CAT 3x2 |
| Stroke syndrome, TACS (%) † | n (%) | xxx | xxx (xx.x) | xxx (xx.x) | xxx (xx.x) | xxx (xx.x) | CAT 3x2 |
| Vascular circulation, posterior (%) † | n (%) | xxx | xxx (xx.x) | xxx (xx.x) | xxx (xx.x) | xxx (xx.x) | CAT 3x2 |
| ***In-hospital care***<br>***pre-randomisation*** | n (%) | xxx | xxx (xx.x) | xxx (xx.x) | xxx (xx.x) | xxx (xx.x) |  |
| Intravenous thrombolysis | n (%) | xxx | xxx (xx.x) | xxx (xx.x) | xxx (xx.x) | xxx (xx.x) | CAT 3x2 |
| Mechanical thrombectomy | n (%) | xxx | xxx (xx.x) | xxx (xx.x) | xxx (xx.x) | xxx (xx.x) | CAT 3x2 |
| Hemicraniectomy | n (%) | xxx | xxx (xx.x) | xxx (xx.x) | xxx (xx.x) | xxx (xx.x) | CAT 3x2 |
| Intensive care unit admission | n (%) | xxx | xxx (xx.x) | xxx (xx.x) | xxx (xx.x) | xxx (xx.x) | CAT 3x2 |
| Ventilation | n (%) | xxx | xxx (xx.x) | xxx (xx.x) | xxx (xx.x) | xxx (xx.x) | CAT 3x2 |

DSRS (dysphagia rating scale); EAT-10: eating assessment tool-10; FOIS: functional oral intake score; FSS: feeding status scale; IDDSI: international dysphagia diet standardisation initiative; NIHSS (NIH stroke severity scale); OSCP: Oxfordshire community stroke project; TACS: total anterior circulation syndrome.
Data are number (%), median [interquartile range] or mean (standard deviation). Comparisons by Cochrane-Armitage test (CAT), or Jonckheere test for ordered alternatives.

**Table S6-2.**
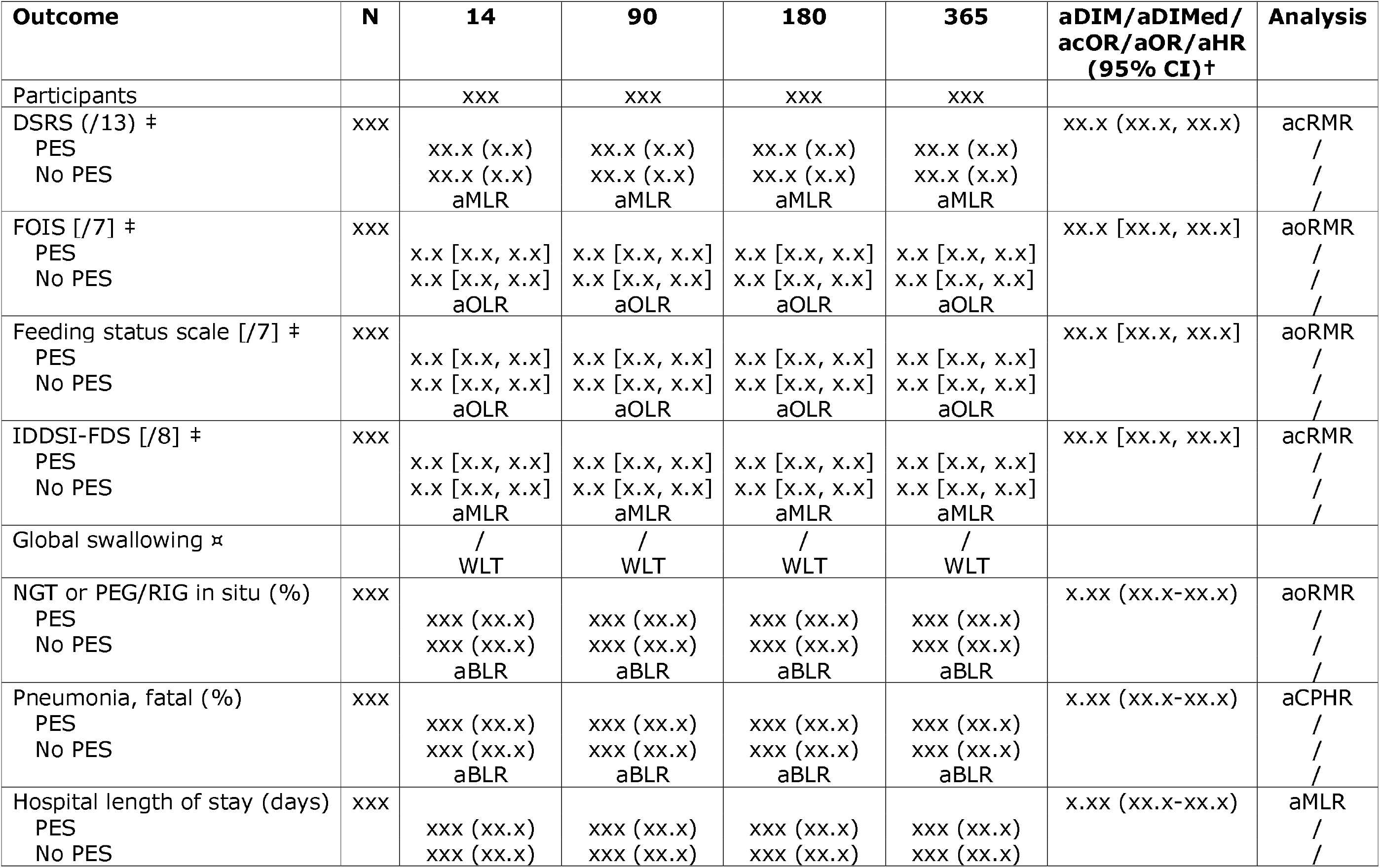
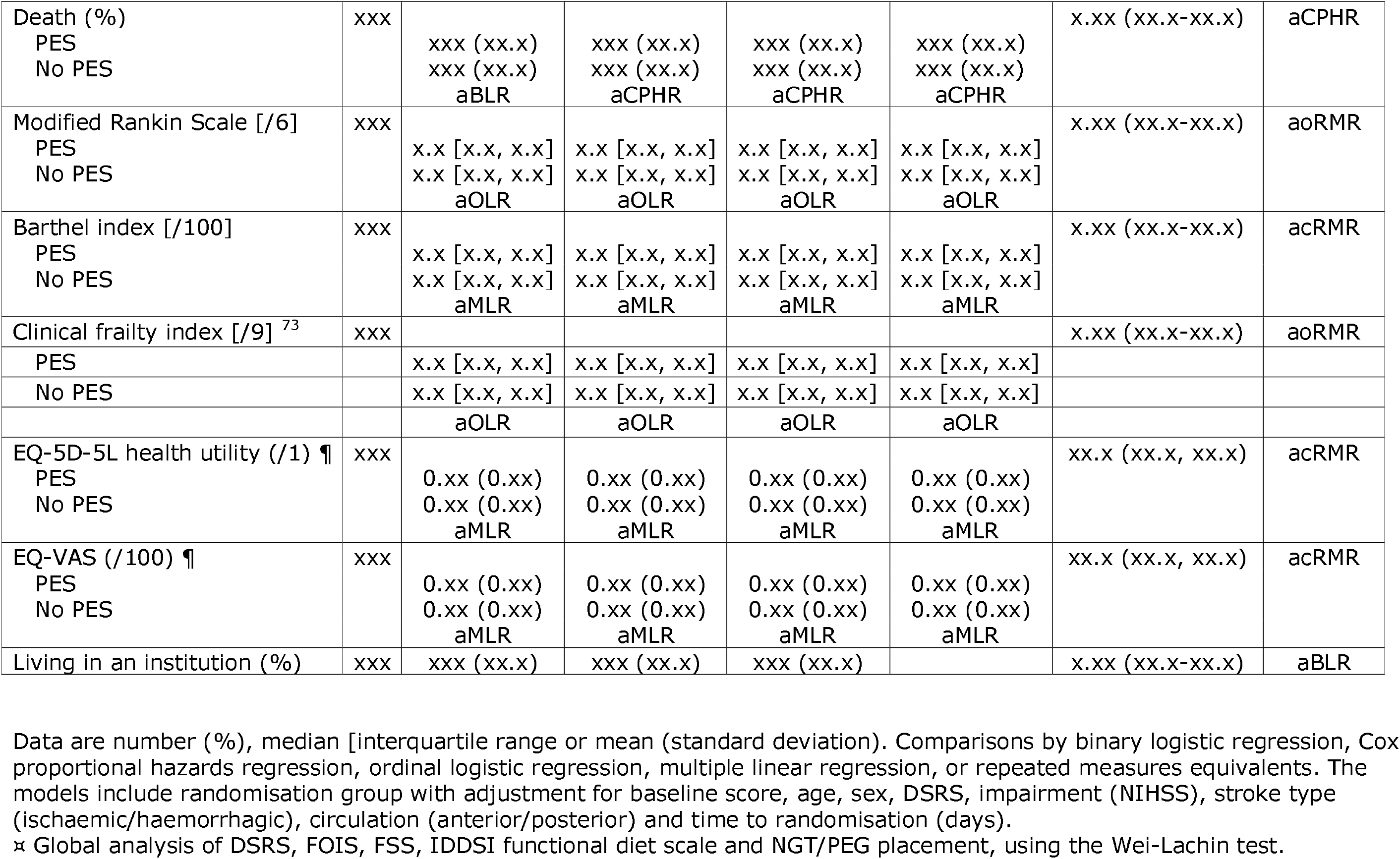
Outcomes at days 14, 90, 180 and 365 in all participants.

**Table S6-3.**
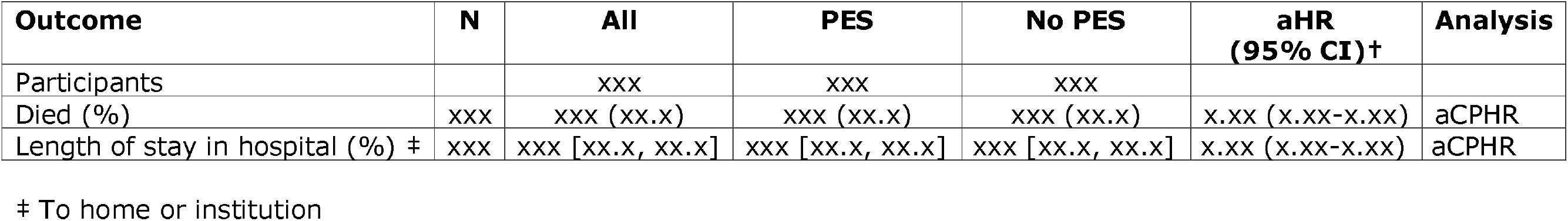
Time to event analyses for length of stay and removal of enteral feeding.

### SUPPLEMENT 7

**COGNITION/DEMENTIA UP TO ONE YEAR**

**Title**

Trajectory of cognition over 1 year in patients with severe ischaemic or haemorrhagic stroke. Data from the PhEAST trial.

**Authors**

TBD

**Aim**

Describe the trajectory of cognition at 5 time points over 1 year in patients with severe ischaemic or haemorrhagic stroke.

**Participants**

All in PhEAST

**Outcomes**

Primary: DSM-5 7-level ordinal cognition.

Secondary: At days 0 and 14 - NIHSS-Cog-4, MoCA and Free-Cog

Secondary: At all timepoints - DSM-5 4-level, t-MoCA, TICS-M, t-Free-Cog, 6-CIT, phenomic fluency and semantic fluency.

Informant: IQCODE.

Others: Clinical frailty index, Zung depression index.

**Analyses**

Descriptive statistics: number (%), median [interquartile range] or mean (standard deviation).

Data will be tabulated in all patients whether alive for not, and in alive patients only.

**Table S7-1.**
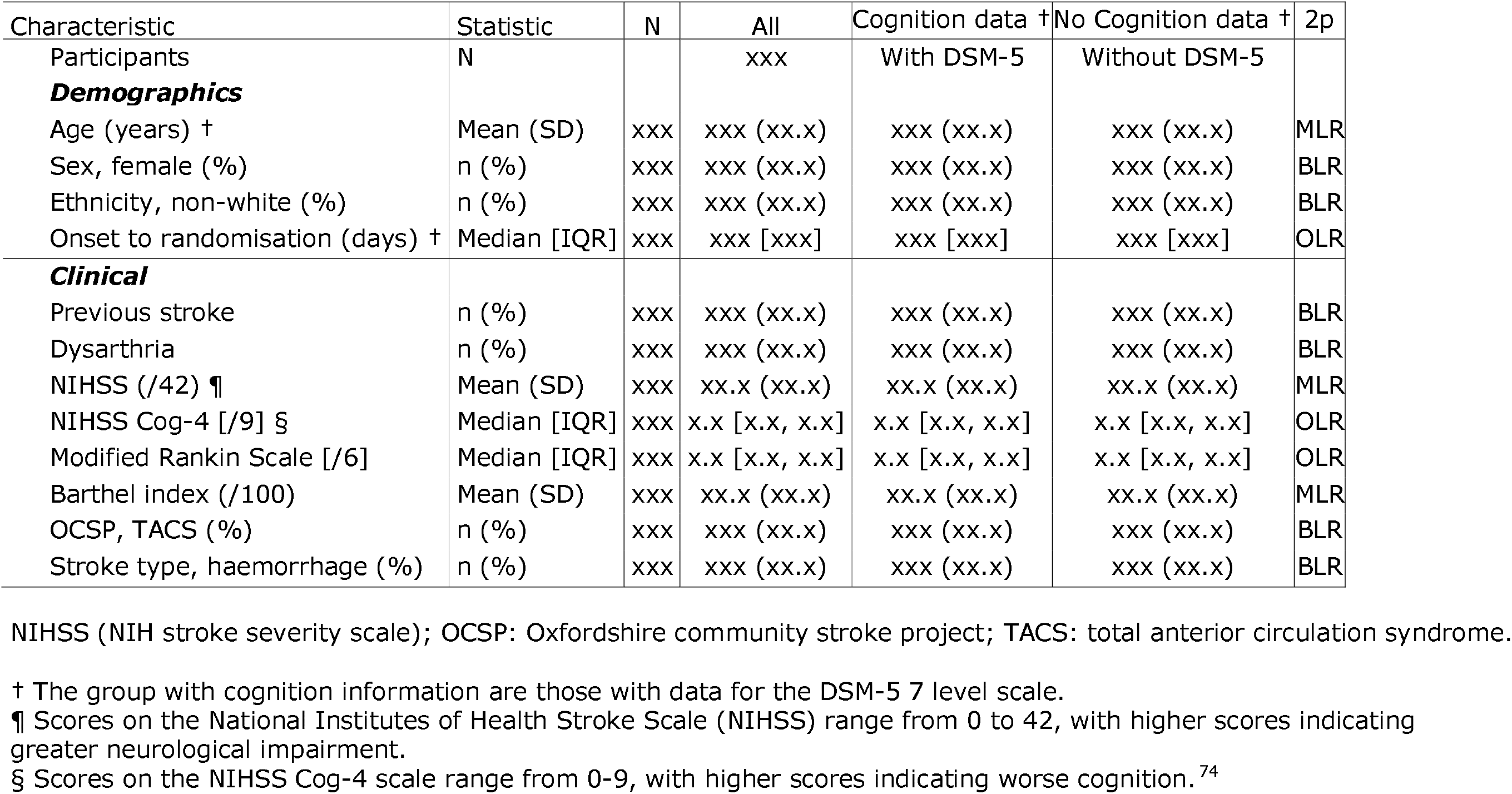
Baseline characteristics. Data are number (%), median [interquartile range or mean (standard deviation).

**Table S7-2.** Cognition variables at baseline and days 14, 90,180 and 365. Results include participants who died with death assigned a score worse than any living score.

| Outcome | N | 0 | 14 | 90 | 180 | 365 | Treatment effect | Test |
| --- | --- | --- | --- | --- | --- | --- | --- | --- |
| Participants |  | xxx | xxx | xxx | xxx | xxx |  |  |
| NIHSS COG-4 [/9] <sup>74</sup><br>§ | xxx |  |  |  |  |  | xx.x (xx.x, xx.x) | aoRMR |
| PES |  | x.x (x.x) | x.x (x.x) | x.x (x.x) | NR | NR |  |  |
| No PES |  | x.x (x.x)<br>/ | x.x (x.x)<br>aMLR | x.x (x.x)<br>aMLR | NR | NR |  |  |
| MOCA (/30) <sup>75</sup> | xxx |  |  |  |  |  | xx.x [xx.x, xx.x] | acRMR |
| PES |  | xx.x (x.x)<br>(N) | xx.x (x.x)<br>(N) | xx.x (x.x)<br>(N) | NR | NR |  |  |
| No PES |  | xx.x (x.x)<br>(N)<br>/ | xx.x (x.x)<br>(N)<br>aMLR | xx.x (x.x)<br>(N)<br>aMLR | NR | NR |  |  |
| t-MOCA (/22) * | xxx |  |  |  |  |  | xx.x [xx.x, xx.x] | acRMR |
| PES |  | xx.x (x.x)<br>(N) | xx.x (x.x)<br>(N) | xx.x (x.x)<br>(N) | xx.x (x.x)<br>(N) | xx.x (x.x)<br>(N) |  | / |
| No PES |  | xx.x (x.x)<br>(N)<br>/ | xx.x (x.x)<br>(N)<br>aMLR | xx.x (x.x)<br>(N)<br>aMLR | xx.x (x.x)<br>(N)<br>aMLR | xx.x (x.x)<br>(N)<br>aMLR |  | / |
| Phonemic fluency<br>(Letter F) <sup>76</sup> | xxx |  |  |  |  |  | xx.x [xx.x, xx.x] | acRMR |
| PES |  | xx.x (x.x)<br>(N) | xx.x (x.x)<br>(N) | xx.x (x.x)<br>(N) | xx.x (x.x)<br>(N) | xx.x (x.x)<br>(N) |  | / |
| No PES |  | xx.x (x.x)<br>(N)<br>/ | xx.x (x.x)<br>(N)<br>aMLR | xx.x (x.x)<br>(N)<br>aMLR | xx.x (x.x)<br>(N)<br>aMLR | xx.x (x.x)<br>(N)<br>aMLR |  | / |
| Category/semantic<br>fluency (animal<br>naming) <sup>76</sup> | xxx |  |  |  |  |  | x.xx (xx.x-xx.x) | acRMR |
| PES |  | xx.x (x.x)<br>(N) | xx.x (x.x)<br>(N) | xx.x (x.x)<br>(N) | xx.x (x.x)<br>(N) | xx.x (x.x)<br>(N) |  |  |
| No PES |  | xx.x (x.x)<br>(N)<br>/ | xx.x (x.x)<br>(N)<br>aMLR | xx.x (x.x)<br>(N)<br>aMLR | xx.x (x.x)<br>(N)<br>aMLR | xx.x (x.x)<br>(N)<br>aMLR |  |  |
| TICS-m (/39) <sup>77</sup><br>PES | xxx | xx.x (xx.x)<br>(N) | xx.x (xx.x)<br>(N) | xx.x (xx.x)<br>(N) | xx.x (xx.x)<br>(N) | xx.x (xx.x)<br>(N) | x.xx (xx.x-xx.x) | acRMR<br>/ |
| No PES |  | xx.x (xx.x)<br>(N)<br>/ | xx.x (xx.x)<br>(N)<br>aMLR | xx.x (xx.x)<br>(N)<br>aMLR | xx.x (xx.x)<br>(N)<br>aMLR | xx.x (xx.x)<br>(N)<br>aMLR |  | / |
| DSM5-4 level [/3] <sup>60</sup><br>PES | xxx | x.x [x.x,<br>x.x] (N) | x.x [x.x,<br>x.x] (N) | x.x [x.x,<br>x.x] (N) | x.x [x.x,<br>x.x] (N) | x.x [x.x,<br>x.x] (N) | x.xx (xx.x-xx.x) | aoRMR<br>/ |
| No PES |  | x.x [x.x,<br>x.x] (N)<br>/ | x.x [x.x,<br>x.x] (N)<br>aOLR | x.x [x.x,<br>x.x] (N)<br>aOLR | x.x [x.x,<br>x.x] (N)<br>aOLR | x.x [x.x,<br>x.x] (N)<br>aOLR |  | / |
| DSM5-7 level [/6] <sup>60</sup><br>PES | xxx | x.x [x.x,<br>x.x] (N) | x.x [x.x,<br>x.x] (N) | x.x [x.x,<br>x.x] (N) | x.x [x.x,<br>x.x] (N) | x.x [x.x,<br>x.x] (N) | x.xx (xx.x-xx.x) | aoRMR<br>/ |
| No PES |  | x.x [x.x,<br>x.x] (N)<br>/ | x.x [x.x,<br>x.x] (N)<br>aOLR | x.x [x.x,<br>x.x] (N)<br>aOLR | x.x [x.x,<br>x.x] (N)<br>aOLR | x.x [x.x,<br>x.x] (N)<br>aOLR |  | / |
| Free-Cog (/30) <sup>78</sup><br>PES | xxx | xx.x (x.x)<br>(N) | xx.x (x.x)<br>(N) | xx.x (x.x)<br>(N) | xx.x (x.x)<br>(N) | xx.x (x.x)<br>(N) | x.xx (xx.x-xx.x) | acRMR |
| No PES |  | xx.x (x.x)<br>(N)<br>/ | xx.x (x.x)<br>(N)<br>aMLR | xx.x (x.x)<br>(N)<br>aMLR | xx.x (x.x)<br>(N)<br>aMLR | xx.x (x.x)<br>(N)<br>aMLR |  |  |
| Tele Free-Cog (/24) <sup>78</sup><br>PES | xxx | xx.x (x.x)<br>(N) | xx.x (x.x)<br>(N) | xx.x (x.x)<br>(N) | xx.x (x.x)<br>(N) | xx.x (x.x)<br>(N) | x.xx (xx.x-xx.x) | acRMR |
| No PES |  | xx.x (x.x) | xx.x (x.x) | xx.x (x.x) | xx.x (x.x) | xx.x (x.x) |  |  |

|  |  | (N)<br>/ | (N)<br>aMLR | (N)<br>aMLR | (N)<br>aMLR | (N)<br>aMLR |  |  |
| --- | --- | --- | --- | --- | --- | --- | --- | --- |
| 6-CIT [/28] <sup>79</sup><br>PES<br>No PES | xxx | x.x [x.x,<br>x.x] (N)<br>x.x [x.x,<br>x.x] (N)<br>/ | x.x [x.x,<br>x.x] (N)<br>x.x [x.x,<br>x.x] (N)<br>aMLR | x.x [x.x,<br>x.x] (N)<br>x.x [x.x,<br>x.x] (N)<br>aMLR | x.x [x.x,<br>x.x] (N)<br>x.x [x.x,<br>x.x] (N)<br>aMLR | x.x [x.x,<br>x.x] (N)<br>x.x [x.x,<br>x.x] (N)<br>aMLR | xx.x (xx.x, xx.x) | acRMR |
| Zung depression<br>scale (/100) <sup>80</sup><br>PES<br>No PES | xxx | x.x [x.x,<br>x.x] (N)<br>x.x [x.x,<br>x.x] (N)<br>/ | x.x [x.x,<br>x.x] (N)<br>x.x [x.x,<br>x.x] (N)<br>aMLR | x.x [x.x,<br>x.x] (N)<br>x.x [x.x,<br>x.x] (N)<br>aMLR | x.x [x.x,<br>x.x] (N)<br>x.x [x.x,<br>x.x] (N)<br>aMLR | x.x [x.x,<br>x.x] (N)<br>x.x [x.x,<br>x.x] (N)<br>aMLR | xx.x (xx.x, xx.x) | acRMR |
| In institution or died<br>(%) | xxx |  |  |  |  |  | x.xx (xx.x-xx.x) | abRMR |
| PES |  | xxx (xx.x)<br>(N) | xxx (xx.x)<br>(N) | xxx (xx.x)<br>(N) | xxx (xx.x)<br>(N) | xxx (xx.x)<br>(N) |  |  |
| No PES |  | xxx (xx.x)<br>(N) | xxx (xx.x)<br>(N) | xxx (xx.x)<br>(N) | xxx (xx.x)<br>(N) | xxx (xx.x)<br>(N) |  |  |
|  |  | / | aBLR | aBLR | aBLR | aBLR |  |  |

Indicted scores for death: NIHSS Cog-4 10, MoCA -1, t-MoCA -1, letter F -1, TICS-m -1, DSM-5 4-level 3, DSM-5 7-level 6, animal naming -1, free-cog -1, t-free-cog -1, 6-CIT 29, clinical frailty 10, Zung depression 102.5.

**Table S7-3.** Cognition variables at baseline and days 14, 90, 180 and 365. *Philip. Results exclude participants who died by the relevant timepoint.

| Measure |  |  |  |  |  |
| --- | --- | --- | --- | --- | --- |
| Day | 0 | 14 | 90 | 180 | 365 |
| Participants | N | N | N | N | N |
| NIHSS COG-4 [/9] <sup>74</sup> § | x.x [x.x, x.x]<br>(N) | x.x [x.x, x.x]<br>(N) | / | / | / |
| MOCA (/30) <sup>75</sup> | xx.x (x.x) (N) | xx.x (x.x) (N) | / | / | / |
| t-MOCA (/22) * | xx.x (x.x) (N) | xx.x (x.x) (N) | xx.x (x.x) (N) | xx.x (x.x) (N) | xx.x (x.x) (N) |
| Phonemic fluency (Letter F) <sup>76</sup> | x.x (x.x) (N) | x.x (x.x) (N) | x.x (x.x) (N) | x.x (x.x) (N) | x.x (x.x) (N) |
| TICS-m (/39) <sup>77</sup> | xx.x (xx.x) (N) | xx.x (xx.x) (N) | xx.x (xx.x) (N) | xx.x (xx.x) (N) | xx.x (xx.x) (N) |
| Cog-R4VaD-4 level [/6] <sup>60</sup> | x.x [x.x, x.x]<br>(N) | x.x [x.x, x.x] (N) | x.x [x.x, x.x]<br>(N) | x.x [x.x, x.x]<br>(N) | x.x [x.x, x.x]<br>(N) |
| Cog-R4VaD-7 level [/6] <sup>60</sup> | x.x [x.x, x.x]<br>(N) | x.x [x.x, x.x] (N) | x.x [x.x, x.x]<br>(N) | x.x [x.x, x.x]<br>(N) | x.x [x.x, x.x]<br>(N) |
| Category/semantic fluency (animal naming) <sup>76</sup> | xx.x (x.x) (N) | xx.x (x.x) (N) | xx.x (x.x) (N) | xx.x (x.x) (N) | xx.x (x.x) (N) |
| Free-Cog (/30) <sup>78</sup> | xx.x (x.x) (N) | xx.x (x.x) (N) | / | / | / |
| Tele Free-Cog (/24) <sup>78</sup> | xx.x (x.x) (N) | xx.x (x.x) (N) | xx.x (x.x) (N) | xx.x (x.x) (N) | xx.x (x.x) (N) |
| 6-CIT [/28] <sup>79</sup> | x.x [x.x, x.x]<br>(N) | x.x [x.x, x.x] (N) | x.x [x.x, x.x]<br>(N) | x.x [x.x, x.x]<br>(N) | x.x [x.x, x.x]<br>(N) |
| Clinical frailty index [/9] <sup>73</sup> | x.x [x.x, x.x]<br>(N) | x.x [x.x, x.x] (N) | x.x [x.x, x.x]<br>(N) | x.x [x.x, x.x]<br>(N) | x.x [x.x, x.x]<br>(N) |
| Zung depression scale (/100) <sup>80</sup> | xx.x (xx.x) (N) | xx.x (xx.x) (N) | xx.x (xx.x) (N) | xx.x (xx.x) (N) | xx.x (xx.x) (N) |
| IQCODE | xx.x (x.x) | xx.x (x.x) | xx.x (x.x) | xx.x (x.x) | xx.x (x.x) |

Note: Data are median [interquartile range] (N) or mean (standard deviation) (N).

### SUPPLEMENT 8

**RESPONDER ANALYSIS**

**Title**

Comparison of patients with severe dysphagia who responded to versus those who did not respond to pharyngeal electrical stimulation. Data from the PhEAST trial.

**Authors**

TBD

**Aim**

Compare baseline characteristics and outcomes in people who responded versus those who did not respond to PES.

**Participants**

All those randomised to PES.

**Definition of response**

At day 90 follow-up: DSRS <4 (vs >=4).

**Outcomes**

Groups: DSRS at day 90 0 or 1 versus 2 to 12 versus died

Baseline: demographic and other characteristics

Follow-up: Swallowing and functional outcomes

**Analyses**

Descriptive statistics: number (%), median [interquartile range] or mean (standard deviation).

Data will be tabulated in three groups.

**Table S8-1.**
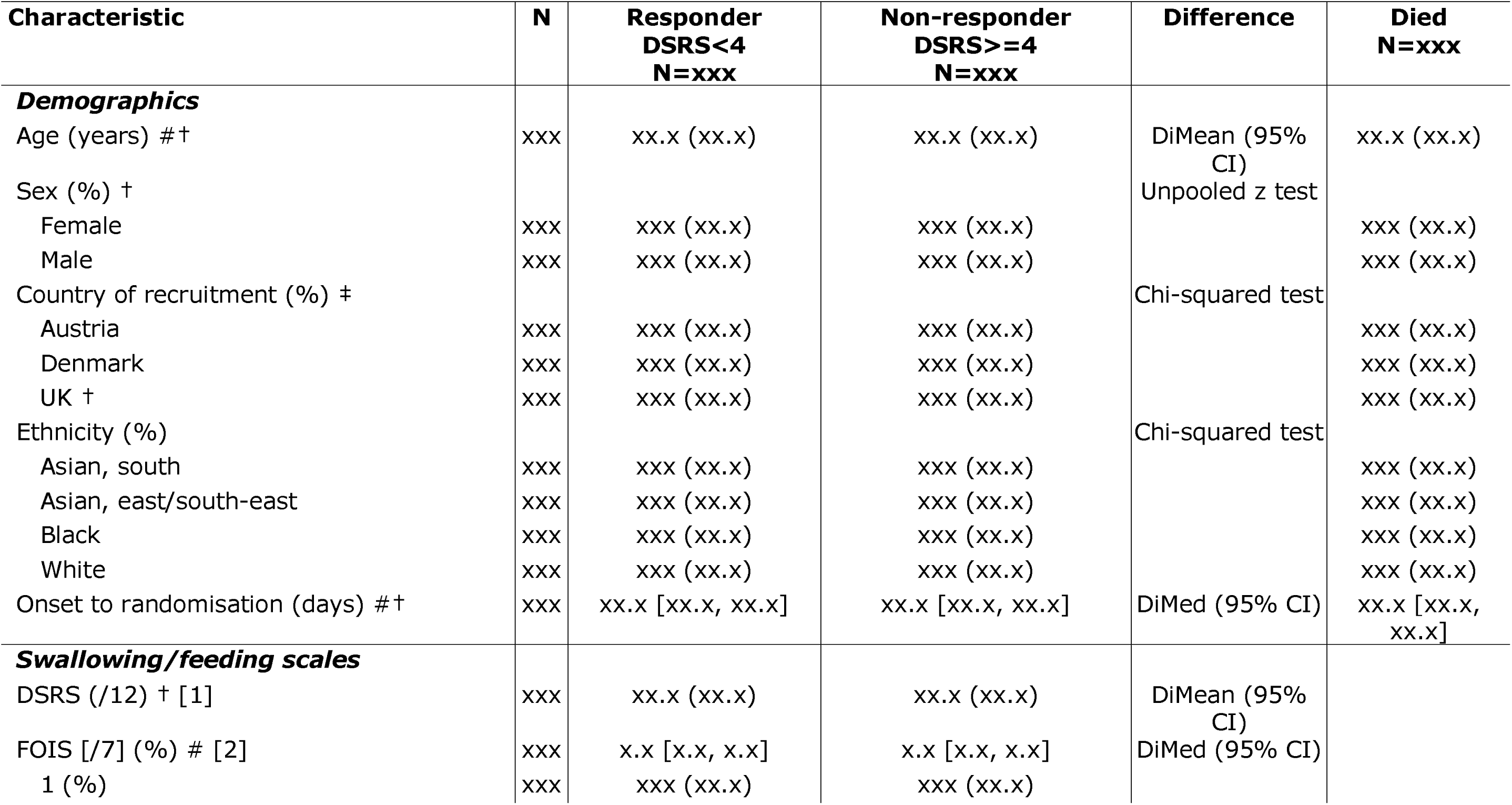
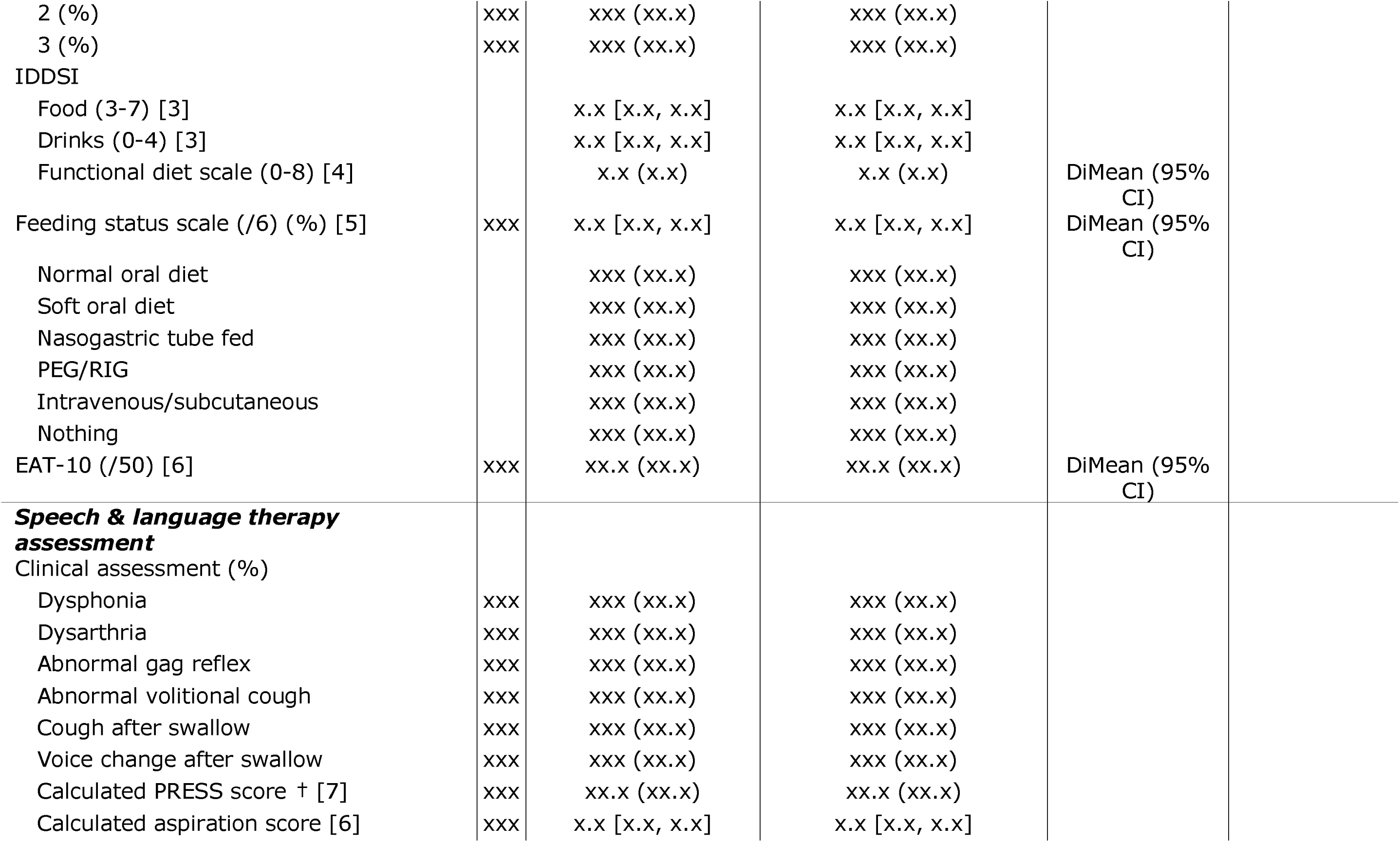
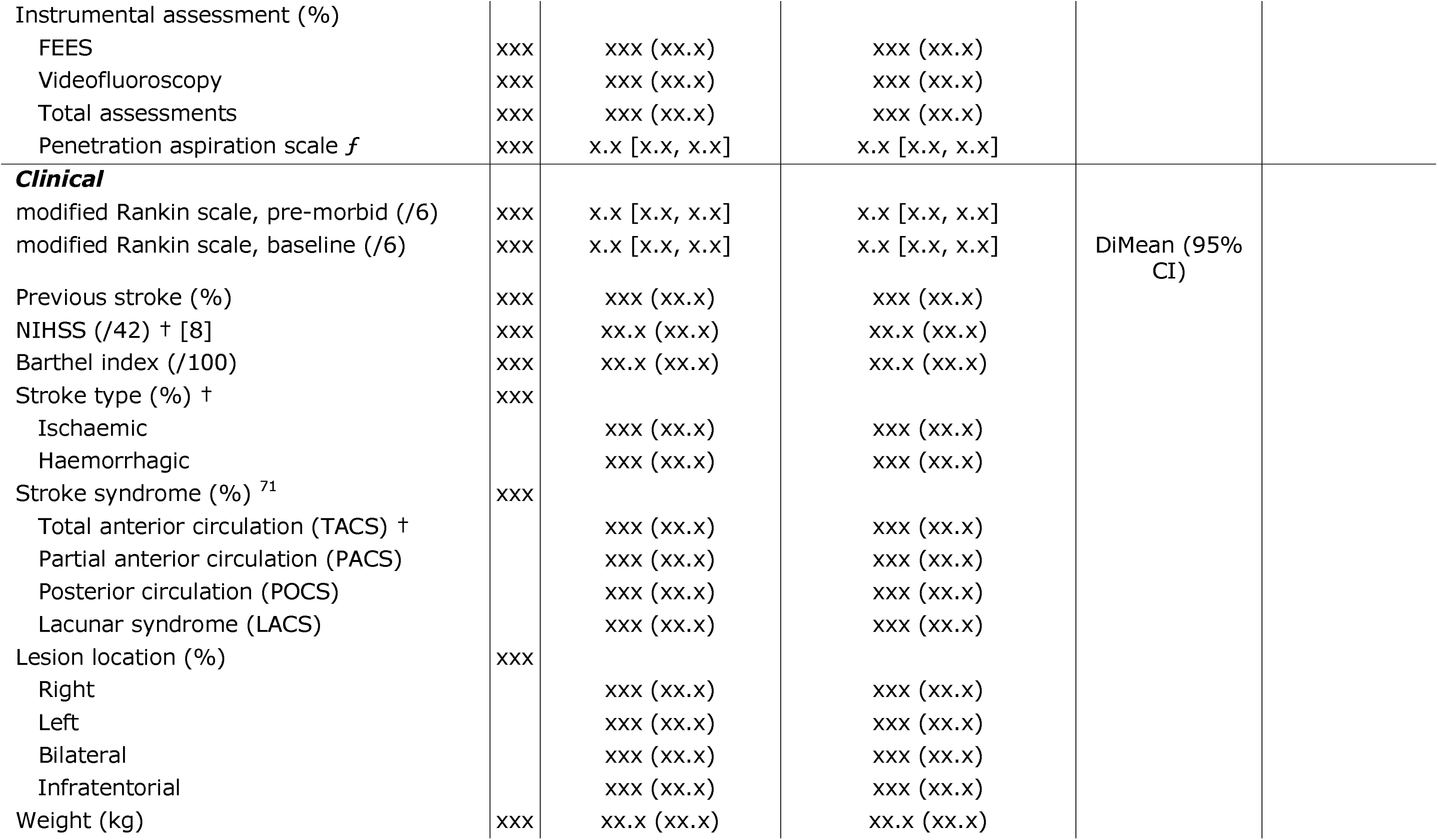
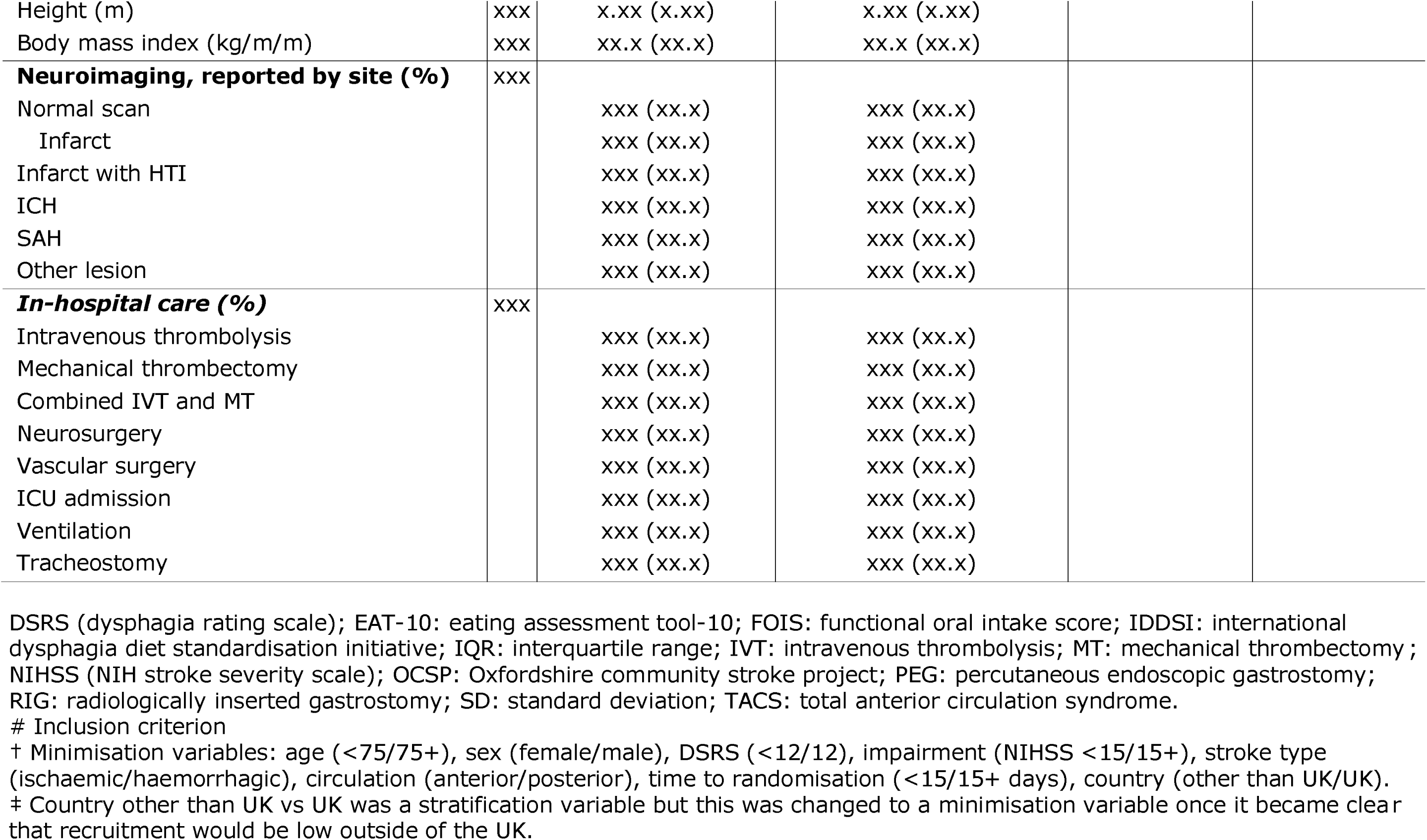
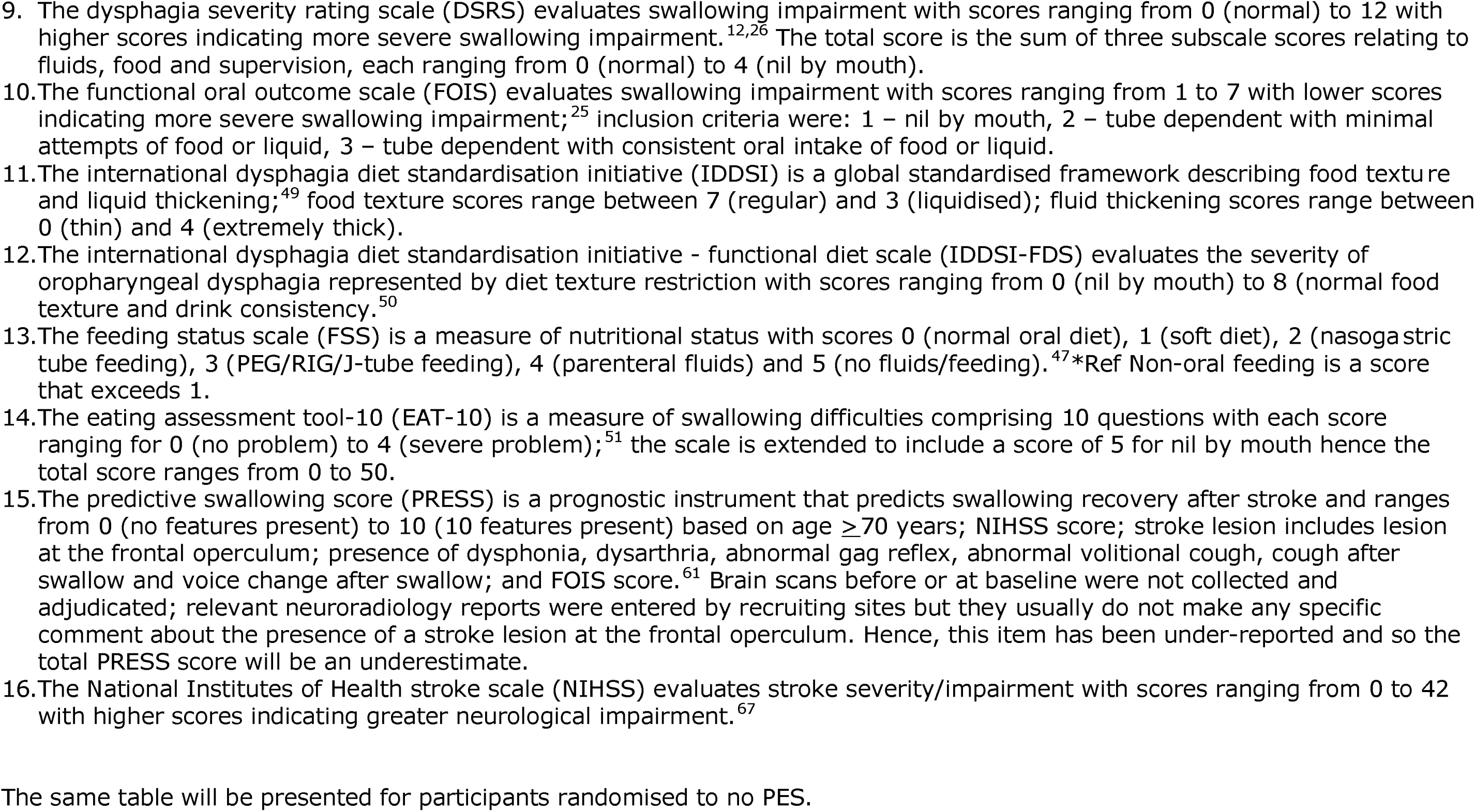
Baseline characteristics by responder group at 90 days. Data are number (%), median [interquartile range] or mean (standard deviation); comparison by unadjusted risk difference, difference in medians (Mann-Whitney u test) or difference in means (Welsh test) with 95% confidence intervals. Information on those who had dies by day are given for completeness.

**Table S8-2.** Comparison of DSRS at day 90 between PES and no PES in pre-specified subgroups.

| **Outcome** | **N** | **All**<br>**(N=xxx)** | **PES**<br>**(N=xxx)** | **No PES**<br>**(N=xxx)** | **aDIM/acOR/aOR**<br>**(95% CI)†** | **Analysis** |
| --- | --- | --- | --- | --- | --- | --- |
| Day 90 | xxx | xx.x (xx.x) | xx.x (xx.x) | xx.x (xx.x) | xx.x (xx.x, xx.x) | aMLR |
| Day 90 change from baseline | xxx | x.x (x.x) | x.x (x.x) | x.x (x.x) | x.x (x.x, x.x) | aMLR |
| Subgroup, change $\geq -1$ vs $< -1$ | | | | | | |
| $\geq -1$ | xxx | xx.x (xx.x) | xx.x (xx.x) | xx.x (xx.x) | xx.x (xx.x, xx.x) | aMLR |
| $< -1$ | xxx | xx.x (xx.x) | xx.x (xx.x) | xx.x (xx.x) | xx.x (xx.x, xx.x) | aMLR |
| Subgroup, change $\geq -2$ vs $< -2$ | | | | | | |
| $\geq -2$ | xxx | xx.x (xx.x) | xx.x (xx.x) | xx.x (xx.x) | xx.x (xx.x, xx.x) | aMLR |
| $< -2$ | xxx | xx.x (xx.x) | xx.x (xx.x) | xx.x (xx.x) | xx.x (xx.x, xx.x) | aMLR |
| Subgroup, change $\geq -3.5$ vs $< -3.5$ | | | | | | |
| $\geq -3.5$ | xxx | xx.x (xx.x) | xx.x (xx.x) | xx.x (xx.x) | xx.x (xx.x, xx.x) | aMLR |
| $< -3.5$ | xxx | xx.x (xx.x) | xx.x (xx.x) | xx.x (xx.x) | xx.x (xx.x, xx.x) | aMLR |
| Subgroup, absolute $\geq 4$ vs $< 4$ | | | | | | |
| $\geq 4$ | xxx | xx.x (xx.x) | xx.x (xx.x) | xx.x (xx.x) | xx.x (xx.x, xx.x) | aMLR |
| $< 4$ | xxx | xx.x (xx.x) | xx.x (xx.x) | xx.x (xx.x) | xx.x (xx.x, xx.x) | aMLR |

**Figure 1. Subgroup analysis of the primary outcome.**

Shown are the difference in means for swallowing impairment assessed using the dysphagia severity rating scale at day 90 in pre-specified subgroups. The widths of the confidence intervals have not been adjusted for multiplicity and should not be used in place of hypothesis testing. FOIS scores range from 1 to 7, with 0 indicating death and lower scores indicating worse swallowing impairment. DSRS scores range from 0 to 12, with 13 indicating death and higher scores indicating worse swallowing impairment.

- Change day 90-0: >=-l, <-l
- Change day 90-0: >=-2, <-2
- Change day 90-0: >=-3.5, <-3.5,
- Absolute day 90: <4, >=4
- Change day 90-0: mean difference

FOREST PLOT WITH INTERACTION TERMS

## REFERENCES

1. Cohen DL, Roffe C, Beavan J, et al. Post-stroke dysphagia: A review and design considerations for future trials. Int J Stroke 2016; 11(4): 399–411.

2. Labeit B, Michou E, Hamdy S, et al. The assessment of dysphagia after stroke: state of the art and future directions. Lancet Neurol 2023; 22(9): 858–70.

3. Labeit B, Michou E, Trapl-Grundschober M, et al. Dysphagia after stroke: research advances in treatment interventions. Lancet Neurol 2024; 23(4): 418–28.

4. Bath PM, Lee HS, Everton LF. Swallowing therapy for dysphagia in acute and subacute stroke. Cochrane Database Syst Rev 2018; 10(10): CD000323

5. NICE. Transcutaneous neuromuscular electrical stimulation for oropharyngeal dysphagia in adults: NICE, 2018.

6. Hamdy S, Aziz Q, Rothwell JC, et al. The cortical topography of human swallowing musculature in health and disease. Nat Med 1996; 2(11): 1217­24.

7. Hamdy S, Rothwell JC, Aziz Q, Singh KD, Thompson DG. Long-term reorganization of human motor cortex driven by short-term sensory stimulation. Nat Neurose! 1998; 1(1): 64–8.

8. Hamdy S, Aziz Q, Rothwell J, et al. Explaining oropharyngeal dysphagia after unilateral hemispheric stroke. The Lancet 1997; 350(9097): 686–92.

9. Suntrup S, Teismann I, Wollbrink A, et al. Pharyngeal electrical stimulation can modulate swallowing in cortical processing and behavior - magnetoencephalographic evidence. Neuroimage 2015; 104: 117–24.

10. Fraser C, Power M, Hamdy S, et al. Driving plasticity in human adult motor cortex is associated with improved motor function after brain injury. Neuron 2002; 34(5): 831–40.

11. Jefferson S, Mistry S, Michou E, Singh S, Rothwell J, Hamdy S. Reversal of a virtual lesion in human pharyngeal motor cortex by high frequency contralesional brain stimulation. Gastroenterology 2009; 137(3): 841–9.

12. Jayasekeran V, Singh S, Tyrrell P, et al. Adjunctive functional pharyngeal electrical stimulation reverses swallowing disability after brain lesions. Gastroenterology 2010; 138(5): 1737–46.

13. Vasant D, Michou E, Tyrrell P, et al. OC-063 Pharyngeal Electrical Stimulation (pes) In Dysphagia Post-acute Stroke: A Double blind, Randomised Trial. Gut 2014; 63(Supplement 1): A31.

14. Scutt P, Lee H, Hamdy S, Bath P. Pharyngeal Electrical Stimulation for Treatment of Poststroke Dysphagia: Individual Patient Data Meta-Analysis of Randomised Controlled Trials. Stroke Research and Treatment 2015; 2015(429053): 8.

15. Bath PM, Scutt P, Love J, et al. Pharyngeal Electrical Stimulation for Treatment of Dysphagia in Subacute Stroke: A Randomized Controlled Trial. Stroke 2016; 47(6): 1562–70.

16. Bath PM, Woodhouse LJ, Suntrup-Krueger S, et al. Pharyngeal electrical stimulation for neurogenic dysphagia following stroke, traumatic brain injury or other causes: Main results from the PHADER cohort study. EClinicalMedicine 2020; 28: 100608.

17. Harvey RL, Smith R, Bathula R, et al. Pharyngeal electrical stimulation to treat dysphagia in acute stroke: learnings from cases in the PhEED clinical trial. J Rehabil Med 2025; 57: jrm43538.

18. NICE. Pharyngeal electrical stimulation for neurogenic dysphagia. London: National Institute for Health and Care Excellence, 2024.

19. Suntrup S, Marian T, Burchard Schroder J, et al. Electrical pharyngeal stimulation for dysphagia treatment in tracheotomized stroke patients: a randomized controlled trial. Intensive Care Medicine 2015; 41(9): 1629–37.

20. Dziewas R, Stellato R, van der Tweel I, et al. Pharyngeal electrical stimulation for early decannulation in tracheotomised patients with neurogenic dysphagia after stroke (PHAST-TRAC): a prospective, single-blinded, randomised trial. Lancet Neurol 2018; 17(10): 849–59.

21. Dziewas R, Michou E, Trapl-Grundschober M, et al. European Stroke Organisation and European Society for Swallowing Disorders guideline for the diagnosis and treatment of post-stroke dysphagia. Eur Stroke J 2021; 6(3): LXXXIX–CXV.

22. Intercollegiate Stroke Working Party. National Clinical Guideline for Stroke for the UK and Ireland. London: Royal College Physicians, 2023.

23. Prabhakaran S, Gonzalez NR, Zachrison KS, et al. 2026 Guideline for the Early Management of Patients With Acute Ischemic Stroke: A Guideline From the American Heart Association/American Stroke Association. Stroke 2026.

24. Benfield JK, Everton LF, Woodhouse LJ, et al. Pharyngeal electrical stimulation versus none for post stroke dysphagia: rationale, design and protocol for the parallel group superiority pharyngeal electrical stimulation for acute stroke dysphagia trial (PhEAST) (2SRCTN98886991). Cerebrovasc Dis Extra 2026: 1–13.

25. Crary MA, Mann GD, Groher ME. Initial psychometric assessment of a functional oral intake scale for dysphagia in stroke patients. Arch Phys Med Rehabil 2005; 86(8): 1516–20.

26. Everton LF, Benfield JK, Hedstrom A, et al. Psychometric assessment and validation of the dysphagia severity rating scale in stroke patients. Sci Rep 2020; 10(1): 7268.

27. Collins R, MacMahon S. Reliable assessment of the effects of treatment on mortality and major morbidity, 2: clinical trials. Lancet 2001; 357(9253): 373–80.

28. Bath PM, Houlton A, Woodhouse L, et al. Statistical analysis plan for the ‘Efficacy of Nitric Oxide in Stroke* (ENOS) trial. Int J Stroke 2014; 9(3): 372–4.

29. Bath PM, Robson K, Woodhouse LJ, et al. Statistical analysis plan for the ‘Triple Antiplatelets for Reducing Dependency after Ischaemic Stroke* (TARDIS) trial. Int J Stroke 2015; 10(3): 449–51.

30. Scutt P, Appleton JP, Dixon M, et al. Statistical analysis plan for the ‘Rapid Intervention with Glyceryl trinitrate in Hypertensive stroke Trial-2 (R2GHT-2)’. Eur Stroke J 2018; 3(2): 193–6.

31. Gamble C, Krishan A, Stocken D, et al. Guidelines for the Content of Statistical Analysis Plans in Clinical Trials. JAMA 2017; 318(23): 2337–43.

32. Hemming K, Kearney A, Gamble C, et al. Prospective reporting of statistical analysis plans for randomised controlled trials. Trials 2020; 21(1): 898.

33. Ali M, Bath PM, Curram J, et al. The Virtual International Stroke Trials Archive. Stroke 2007; 38(6): 1905–10.

34. Sandercock PA, Niewada M, Czlonkowska A, International Stroke Trial Collaborative G. The International Stroke Trial database. Trials 2011; 12(1): 101.

35. Enos Trial Investigators. Efficacy of nitric oxide, with or without continuing antihypertensive treatment, for management of high blood pressure in acute stroke (ENOS): a partial-factorial randomised controlled trial. Lancet 2015; 385(9968): 617–28.

36. Bath PM, Woodhouse LJ, Appleton JP, et al. Antiplatelet therapy with aspirin, clopidogrel, and dipyridamole versus clopidogrel alone or aspirin and dipyridamole in patients with acute cerebral ischaemia (TARD1S): a randomised, open-label, phase 3 superiority trial. Lancet 2018; 391(10123): 850–9.

37. Bath PM, Scutt P, Blackburn DJ, et al. Intensive versus Guideline Blood Pressure and Lipid Lowering in Patients with Previous Stroke: Main Results from the Pilot ‘Prevention of Decline in Cognition after Stroke Trial* (PODCAST) Randomised Controlled Trial. PLoS One 2017; 12(1): e0164608.

38. Sprigg N, Flaherty K, Appleton JP, et al. Tranexamic acid for hyperacute primary ZntraCerebral Haemorrhage (T2CH-2): an international randomised, placebo-controlled, phase 3 superiority trial. Lancet 2018; 391(10135): 2107–15.

39. Right-Investigators. Prehospital transdermal glyceryl trinitrate in patients with ultra-acute presumed stroke (RIGHT-2): an ambulance-based, randomised, sham-controlled, blinded, phase 3 trial. Lancet 2019; 393(10175): 1009–20.

40. Weir CJ, Lees KR. Comparison of stratification and adaptive methods for treatment allocation in an acute stroke clinical trial. Stat Med 2003; 22: 705–26.

41. Hernandez AV, Steyerberg EW, Butcher I, et al. Adjustment for strong predictors of outcome in traumatic brain injury trials: 25% reduction in sample size requirements in the IMPACT study. J Neurotrauma 2006; 23(9): 1295–303.

42. Optimising the Analysis of Stroke Trials C, Gray LJ, Bath PM, Collier T. Should stroke trials adjust functional outcome for baseline prognostic factors? Stroke 2009; 40(3): 888-94.

43. Gray LJ, Collier T, Bath PMW, on behalf of the OAST Collaboration. Calculation of sample size for stroke trials assessing functional outcome: comparison of binary and ordinal approaches. European Stroke Conference; 2008; Nice, France; 2008.

44. Lees KR, Bath PMW, Schellinger PD, et al. Contemporary outcome measures in acute stroke research: choice of primary outcome measure Stroke 2012; 43(4): 1163–70.

45. Schellinger PD, Bath PMW, Lees K, et al. Assessment of addtional endpoints relevant to the benefit of patients after stroke - what, when, where, in whom. Int J Stroke 2012; 7(3): 227–30.

46. Bath PMW, Woodhouse L, Scutt P, et al. Management of high blood pressure in acute stroke: Efficacy of Nitric Oxide in Stroke (ENOS), a partial-factorial randomised controlled trial. Lancet 2015; 385(9968): 617–28.

47. Woodhouse LJ, Scutt P, Hamdy S, et al. Route of Feeding as a Proxy for Dysphagia After Stroke and the Effect of Transdermal Glyceryl Trinitrate: Data from the Efficacy of Nitric Oxide in Stroke Randomised Controlled Trial. Transl Stroke Res 2018; 9(2): 120–9.

48. Wilkinson G, Everton LF, Krishnan K, Benfield J, Hamdy S, Bath PM. Mode of nutrition as a reflection of swallowing ability in acute and sub-acute stroke: Validation of a bedside tool. J Stroke Cerebrovasc Dis 2025; 34(12): 108484.

49. Cichero JA, Lam P, Steele CM, et al. Development of International Terminology and Definitions for Texture-Modified Foods and Thickened Fluids Used in Dysphagia Management: The IDDSI Framework. Dysphagia 2017; 32(2): 293–314.

50. Steele CM, Namasivayam-MacDonald AM, Guida BT, et al. Creation and Initial Validation of the International Dysphagia Diet Standardisation Initiative Functional Diet Scale. Arch Phys Med Rehabil 2018; 99(5): 934–44.

51. Belafsky PC, Mouadeb DA, Rees CJ, et al. Validity and reliability of the Eating Assessment Tool (EAT-10). Ann Otol Rhinol Laryngol 2008; 117(12): 919–24.

52. Jenkinson C, Fitzpatrick R, Crocker H, Peters M. The Stroke Impact Scale: validation in a UK setting and development of a SIS short form and SIS index. Stroke 2013; 44(9): 2532–5.

53. Zung WW, Richards CB, Short MJ. Self-rating depression scale in an outpatient clinic. Further validation of the SDS. Arch Gen Psychiatry 1965; 13(6): 508–15.

54. Quinn TJ, Dawson J, Lees JS, Chang TP, Walters MR, Lees KR. Time spent at home poststroke: “Home-time” a meaningful and robust outcome measure for stroke trials. Stroke 2008; 39: 231–3.

55. EMEA. Points to Consider on Multiplicity Issues in Clinical Trials. EMEA 2002.

56. Schulz KF, Grimes DA. Multiplicity in randomised trials 11: subgroup and interim analyses. Lancet 2005; 365(9471): 1657–61.

57. Wilkinson G, McLaughlin C, Rehman H, Hamdy S, Bath PM. Treatment of post-stroke dysphagia with pharyngeal electrical stimulation: the multicentre assessment of pharyngeal electrical stimulation (MAPS) Clinical Registry. Submitted 2026.

58. Kenedi H, Campbell-Vance J, Reynolds J, et al. Implementation and Analysis of a Free Water Protocol in Acute Trauma and Stroke Patients. Crit Care Nurse 2019; 39(3): e9-el7.

59. Rosenbek J, Robbins J, Roecker E, Coyle J, Wood J. A penetration-aspiration scale. Dysphagia 1996; 11(2): 93–8.

60. Wardlaw JM, Doubal F, Brown R, et al. Rates, risks and routes to reduce vascular dementia (R4vad), a UK-wide multicentre prospective observational cohort study of cognition after stroke: Protocol. Eur Stroke J 2021; 6(1): 89–101.

61. Galovic M, Stauber AJ, Leisi N, et al. Development and Validation of a Prognostic Model of Swallowing Recovery and Enteral Tube Feeding After Ischemic Stroke. JAMA Neurol 2019.

62. Scutt P, Lee HS, Hamdy S, Bath PM. Pharyngeal Electrical Stimulation for Treatment of Poststroke Dysphagia: Individual Patient Data Meta­Analysis of Randomised Controlled Trials. Stroke Res Treat 2015; 2015: 429053.

63. Ali M, Bath PMB, Davis SM, et al. The virtual international stroke trials archive (VISTA). Stroke 2007; 38: 1905–10.

64. Curtis L, Burns A. Unit Costs of Health and Social Care 2023. Canterbury: Personal Social Services Research Unit, 2023.

65. White IR, Horton NJ, Carpenter J, Pocock SJ. Strategy for intention to treat analysis in randomised trials with missing outcome data. BMJ (Clinical research ed) 2011; 342: d40.

66. White IR, Royston P, Wood AM. Multiple imputation using chained equations: Issues and guidance for practice. Stat Med 2011; 30(4): 377–99.

67. Lyden P, Brott T, Tilley B, et al. Improved reliability of the NIH Stroke Scale using video training. NINDS TPA Stroke Study Group. Stroke 1994; 25(11): 2220–6.

68. Wei L, Lachin J. Two-Sample Asymptotically Distribution-Free Tests for Incomplete Multivariate Observations. Journal of the American Statistical Association 1984; 79(387): 653–61.

69. Lachin JM. Applications of the Wei-Lachin multivariate one-sided test for multiple outcomes on possibly different scales. PLoS One 2014; 9(10): el08784.

70. Wardlaw JM, Woodhouse LJ, Mhlanga, II, et al. Isosorbide Mononitrate and Cilostazol Treatment in Patients With Symptomatic Cerebral Small Vessel Disease: The Lacunar Intervention Trial-2 (LAC1-2) Randomized Clinical Trial. JAMA Neurol 2023; 80(7): 682–92.

71. Bamford J, Sandercock P, Dennis M, Burn J, Warlow C. Classification and natural history of clinically identifiable subtypes of cerebral infarction. Lancet 1991; 337(8756): 1521–6.

72. Dhillon PS, Butt W, Podlasek A, et al. Effect of proximal blood flow arrest during endovascular thrombectomy (ProFATE): Study protocol for a multicentre randomised controlled trial. Eur Stroke J 2023; 8(2): 581–90.

73. Rockwood K, Song X, MacKnight C, et al. A global clinical measure of fitness and frailty in elderly people. Canadian Medical Association Journal 2005; 173(5): 489–95.

74. Cumming TB, Blomstrand C, Bernhardt J, Linden T. The N1H stroke scale can establish cognitive function after stroke. Cerebrovascular diseases (Basel, Switzerland) 2010; 30(1): 7–14.

75. Nasreddine ZS, Phillips NA, Bedirian V, et al. The Montreal Cognitive Assessment, MoCA: a brief screening tool for mild cognitive impairment. Journal of the American Geriatrics Society 2005; 53(4): 695–9.

76. Tombaugh TN, Kozak J, Rees L. Normative data stratified by age and education for two measures of verbal fluency: FAS and animal naming. Arch Clin Neuropsychol 1999; 14(2): 167–77.

77. Desmond DW, Tatemichi TK, Hanzawa L. The telephone interview for cognitive status (TICS): Reliability and validity in a stroke sample. Int J Geriatr Psychiatry 1994; 9: 803–7.

78. Burns A, Harrison JR, Symonds C, Morris J. A novel hybrid scale for the assessment of cognitive and executive function: The Free-Cog. Int J Geriatr Psychiatry 2021; 36(4): 566–72.

79. Brooke P, Bullock R. Validation of a 6 item cognitive impairment test with a view to primary care usage. Int J Geriatr Psychiatry 1999; 14(11): 936–40.

80. Zung WWK. A self-rating depression scale. Arch Gen Psychiatry 1965; 12: 63–70.

